# The clinical, humanistic, and economic outcomes, including experiencing of patient safety events, associated with admitting patients to single rooms compared with shared accommodation for acute hospital admissions. A narrative synthesis systematic literature review

**DOI:** 10.1101/2022.09.27.22280411

**Authors:** Andrea Bertuzzi, Alison Martin, Nicola Clarke, Cassandra Springate, Rachel Ashton, Wayne Smith, Andi Orlowski, Duncan McPherson

## Abstract

**Objectives:** Assess the impact of single rooms versus multioccupancy accommodation on inpatient health-care outcomes and processes.

**Design:** Systematic review.

**Setting:** Hospitals and secondary care units.

**Participants:** Inpatients receiving routine, emergency, high-dependency, or intensive care with a named type of hospital accommodation.

**Main outcome measures:** Qualitative synthesis of findings.

**Results:** Of 4,861 citations initially identified, 215 were deemed suitable for full-text review, of which 145 were judged to be relevant to this review. Five main method types were reported: 60 before - and-after comparisons, 75 contemporaneous comparisons, 18 qualitative studies of accommodation preferences, 10 evidence syntheses. All studies had methodological issues that potentially biased the results by not adjusting for confounding factors that are likely to have contributed to the outcomes. Ninety-two papers compared clinical outcomes for patients in single rooms versus shared accommodation, but no clearly consistent conclusions could be drawn about overall benefits of single rooms versus shared accommodation (multioccupancy rooms, bays, or wards). Single rooms were most likely to be associated with a small overall clinical benefit for the most severely ill patients, especially neonates in intensive care. Patients who preferred single rooms tended to do so for privacy, and for reduced disturbances. By contrast, men, older adults, children, and adolescents were more likely to prefer shared accommodation to avoid loneliness. While shared accommodation seemed to be the most cost-effective approach for construction, greater costs associated with building single rooms were small and likely to be recouped over time by other efficiencies.

**Conclusions:** The lack of difference between inpatient accommodation types in a large number of studies suggests that there would be little effect on clinical outcomes, particularly in routine care. Patients in intensive care areas are most likely to benefit from single rooms. Most patients preferred single rooms for privacy and some preferred shared accommodation for avoiding loneliness.

**Summary:** *What is already known on this topic:*

- The effects of single rooms versus shared accommodation on hospital inpatients’ outcomes are not well understood
- Many studies are qualitative or narrative because randomised controlled trials are not practical and most comparative studies have only become possible after relocation to new facilities
- This systematic review investigated the potential range of impacts that inpatient single rooms and shared accommodation have on the health-care processes, outcomes, and costs

*What this study adds:*

- The evidence, though extensive, revealed no clear advantage for one type of inpatient hospital accommodation for many of the areas assessed.
- There was weak evidence indicating advantages for single bedrooms in some areas, such as lower risk of hospital acquired infection in adult intensive care and a range of outcomes in neonatal intensive care.
- Most patients preferred single rooms for privacy and some preferred shared accommodation for avoiding loneliness.

## INTRODUCTION

The UK government announced that 40 new hospitals will be delivered in England by 2030.(1) The majority will be acute secondary care hospitals. One decision is whether beds should be in a single or multioccupancy room. Once each hospital is built it is difficult to change the proportion of single rooms to shared accommodation. It is important to get this right at the start. Many views have been expressed on the correct proportion of single rooms in a hospital in England. NHS England’s National Medical Director, Stephen Powis, believes in “…single rooms being the default…”.(2) In response David Oliver wrote that, “… our goal [should] perhaps be a greater proportion of single rooms, rather than these exclusively”.(3) The topic has been debated in the British Medical Journal.(4) There is a need for a systematic analysis of the evidence on what is most likely to yield the best outcome for patients..

For some situations isolation of the patient in a single room is part of the clinical intervention. For example, a patient with severe immune compromise may be isolated to protect them from acquiring infection. Similarly, patients with highly transmissible infections may be isolated to prevent spread of infection. A single room is also used where privacy is extremely important, for example delivery units on maternity wards or for dying patients and their families. For most patients, accommodation in either a single or multioccupancy room is possible and may have a range of balanced risks and benefits. Patients may have a range of reasons for their preference including what would count as a good experience of hospital admission. However, there is no settled, obvious evidence base to illustrate what type of accommodation is best for overall patient outcomes or patient experience.

This study set out to find published evidence to investigate whether inpatient stays in single rooms or in shared accommodation (i.e., multioccupancy rooms, bays, or wards), have been associated with any impact on the processes undertaken by the hospital and on patients’ outcomes. A wide range of clinical, social, and economic outcomes were included from the primary perspective of patients across a range of acute hospital types. Staff perspectives, while not formally assessed, were included if reported as part of a study on patient and caregiver views. The objective was to compare staying in a single room versus shared accommodation for care in which the type of accommodation was not part of the intervention itself. This systematic review protocol has been registered with PROSPERO, registration number CRD42022311689. Ethics approval was not required for this study.

## METHODS

### Identification of papers

We performed a systematic literature review of content in Medline (via PubMed) and Embase for comparative clinical trials, observational studies, and systematic literature reviews published in any language up to 17 February 2022. Additional searches were performed via Google Scholar and the National Institute for Health and Care Excellence. We used combinations of “hospital”, design”, “management”, “health care facility”, “single”, “multi”, “room”, “bay”, “bed”, and “accommodation”, optimised for the search platform (see supplementary information – Appendix: Search Strategy). Eligible papers addressed care of adult and/or paediatric inpatients staying in hospital for routine, emergency, or intensive care and who were assigned to a particular accommodation type (single room or shared accommodation). We excluded papers that assessed long-stay patients, day patients, and those attending accident and emergency departments who were not later admitted to an acute hospital; patients who were relocated to a single room during admission (e.g., for isolation after contracting and infectious disease or for terminal care); no direct comparison condition for staying in a single room; non-clinical outcomes; and impact of care on health-care professionals and/or support staff. We also excluded narrative reviews, perspective papers, letters, editorials, and conference abstracts with no relevant data.

Retrieved abstracts were screened by two researchers (AB and NC) using the inclusion criteria in the appendix. Disagreements were resolved by discussion with the project leader. Shortlisted papers were retrieved as full texts. The reference lists of all papers included in this analysis were reviewed to identify any additional publications of primary research that met the inclusion criteria. Full papers were screened for relevance by two researchers (AB and AM) independently.

The quality of each paper was assessed by the same researcher using the Downs and Black checklist for observational studies(5) and the Joanna Briggs Institute checklists (https://jbi.global/critical-appraisal-tools) for qualitative studies and for systematic reviews. These checklists enable assessment of reporting quality, generalisability of findings, biases in measurements of intervention and outcome, confounding in the selection of participants, and power (whether negative findings could be the result of chance). Each quality assessment checklist score was converted to a percentage of the maximum possible score and were categorised for the purposes of this report into high (75–100%), moderate (50–74%), or low quality (<50%; see supplementary table 1).

### Data extraction and synthesis

Data were extracted into an Excel spreadsheet and checked, with final adjudication by the project leader (AM), and were synthesised narratively, according to the methods of Campbell et al.(6) The fields for extraction were study methodology, baseline characteristics of participants (when provided), clinical outcomes, non-clinical outcomes, resource use, and costs. The clinical outcomes of interest were in-hospital mortality, overall mortality (≥30 days), morbidity (e.g., falls, deterioration, new pressure ulcers, and complications), patient safety incidents, and hospital-acquired infections. Non-clinical outcomes of interest were patient and family member experiences, length of stay, cost of stay, experience of accommodation change and number of changes (for the same type of care) during admission, and impact on the caregivers and family members of dependent patients. Outcomes were assessed based on the measures used in the original articles. Extracted data were sorted by outcome and then by population and setting. Relevant data for each outcome were summarised narratively by comparing heterogeneity across studies in terms of whether differences were statistically significant and in favour of single room or shared accommodation.

### Statistical analysis

As substantial heterogeneity across studies (e.g., how data were reported, study methods, etc) was expected, formal meta-analysis was not deemed feasible. Thus, no formal measures of heterogeneity or overall effect size were performed, and all data reported are descriptive. To aid comparison and assess consistency of the conclusions, data are presented in summary tables.

Certainty of findings was assessed based on whether the direction of benefit was consistently statistically significant for single rooms or shared accommodation (across all studies or those with the lowest risk of bias) or was inconsistent or not statistically significant.

There are special areas of hospitals where responses to the intervention might differ, such as intensive-care or paediatric units and areas for women in labour. Therefore, we aimed to present data separately by different subgroups.

### Patient and Public Involvement

Patients and the public were not involved in designing this study. The study aim was to establish what is already published in peer reviewed literature on the topic, including the views of patients, their parents or caregivers, and the public.

## RESULTS

### Study characteristics

The initial searches returned 4,861 potentially relevant abstracts. After screening and removal of duplicates, 145 publications were included in this review (Figure 1). There were six main types of studies: 60 before-and-after comparisons (shared accommodation followed by relocation to single-rooms); 75 comparisons of patients allocated to single rooms compared with others simultaneously in shared accommodation; 18 qualitative studies recording the views of patients, caregivers, or healthcare professionals on accommodation preferences; 10 evidence syntheses, including systematic literature reviews, guidelines and other reports; and three economic evaluations of accommodation type (Figure 2). Some studies incorporated more than one design.

**Figure 1.**
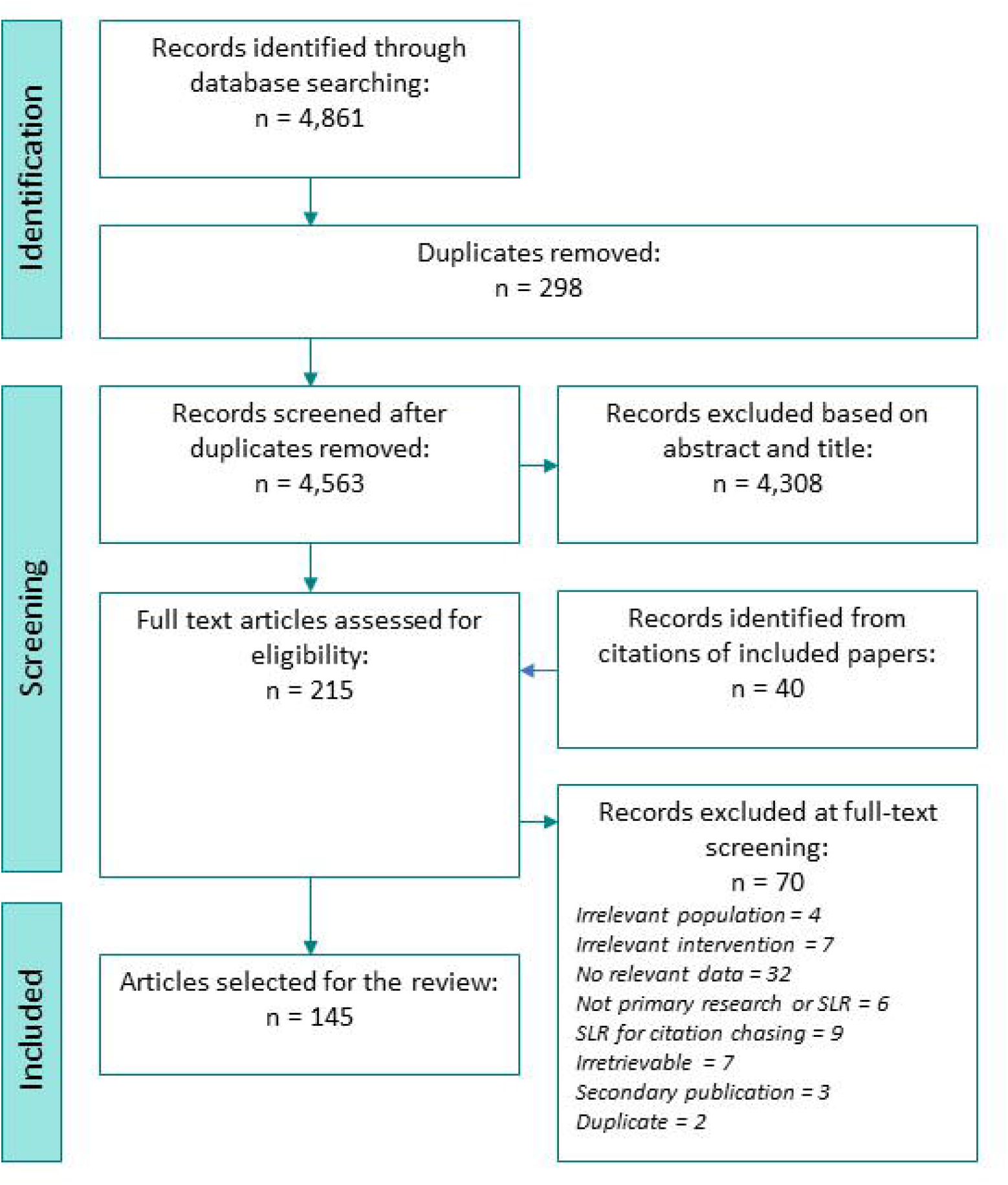
Selection of papers for review.

**Figure 2.**
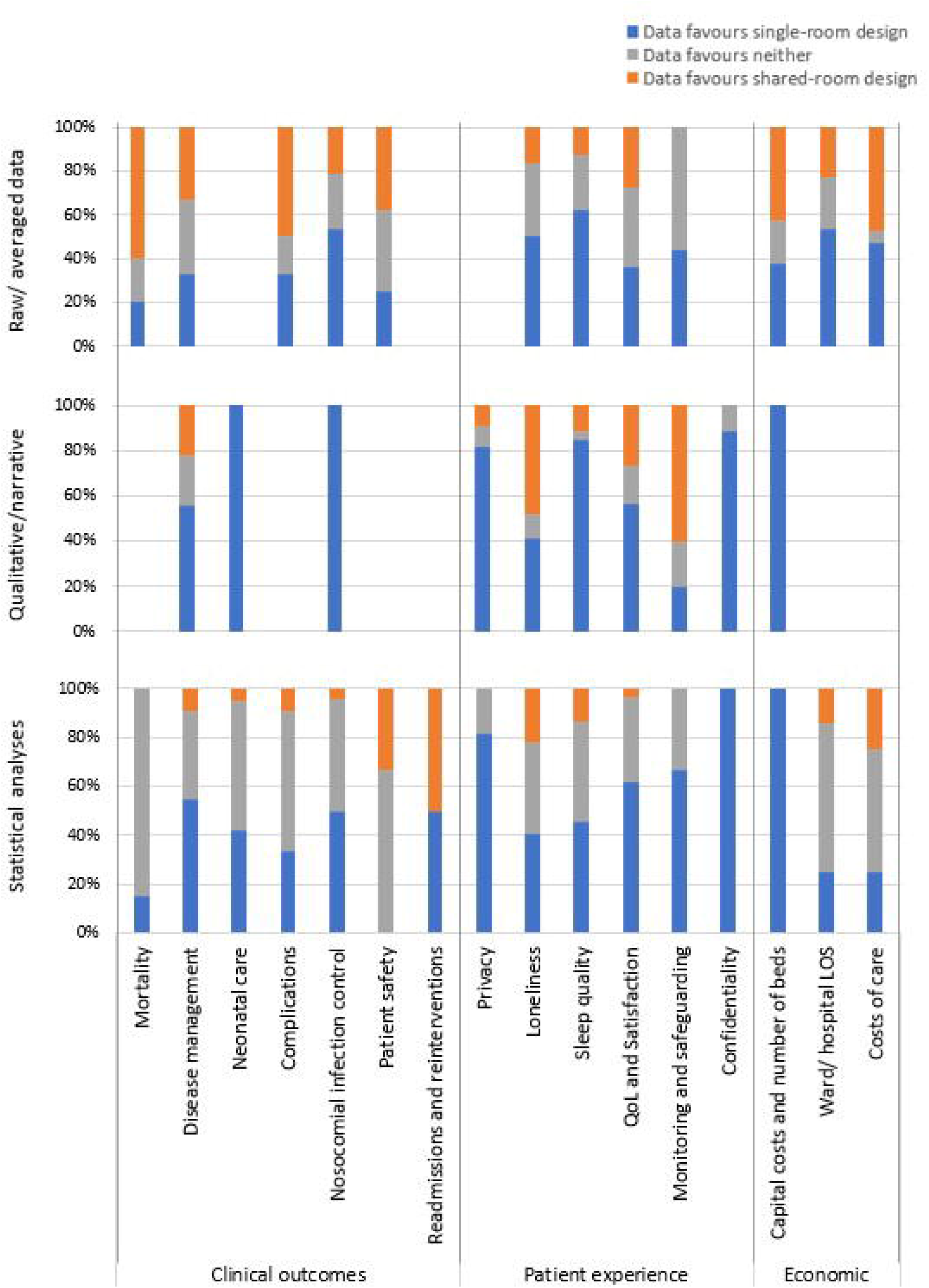
Percentage of studies reporting data in favour of either single-room or shared-room design, according to the type of data available and outcome reported.

All studies had methodological issues that potentially biased the results by not adjusting for confounding factors that are likely to have contributed to the outcomes. In the 60 before-and-after trials, many factors other than accommodation changed due to moving into new facilities, such as unfamiliarity with new layouts and logistics. In the 75 contemporaneous comparisons, reasons for bed space allocations were not generally reported (e.g., availability, severity of illness), making their effects on the differences in outcomes unclear. Nine studies did not report baseline characteristics, and of those that did, only three reported no significant difference between age, sex, and comorbidity or health status of patients at baseline.

The quality of studies varied widely. Thirty-four studies were assigned high quality scores (75⍰100%) with a range of 78⍰100% (see supplementary table 1). Twenty-three studies were classified as being of low quality (<50%) with a range of 10⍰48%.

### Mortality

Eighteen studies reported mortality (see Figure 3 and supplementary Table 2).(7–24) Ten were before-and-after studies and the others were contemporary studies. Only one article scored less than 50% for quality and two had high quality scores. Six studies involved neonates/infants, one assessed children, and the remainder were concerned with adult/elderly care. The numbers of deaths were low, meaning that the studies might not have had enough patient-years of follow-up to detect small but statistically significant differences in mortality. Likewise, whether reported increases in mortality reflected true increases in risk or were due to confounding factors (e.g., unreported reasons for patients being allocated single rooms) is unclear.

**Figure 3.**
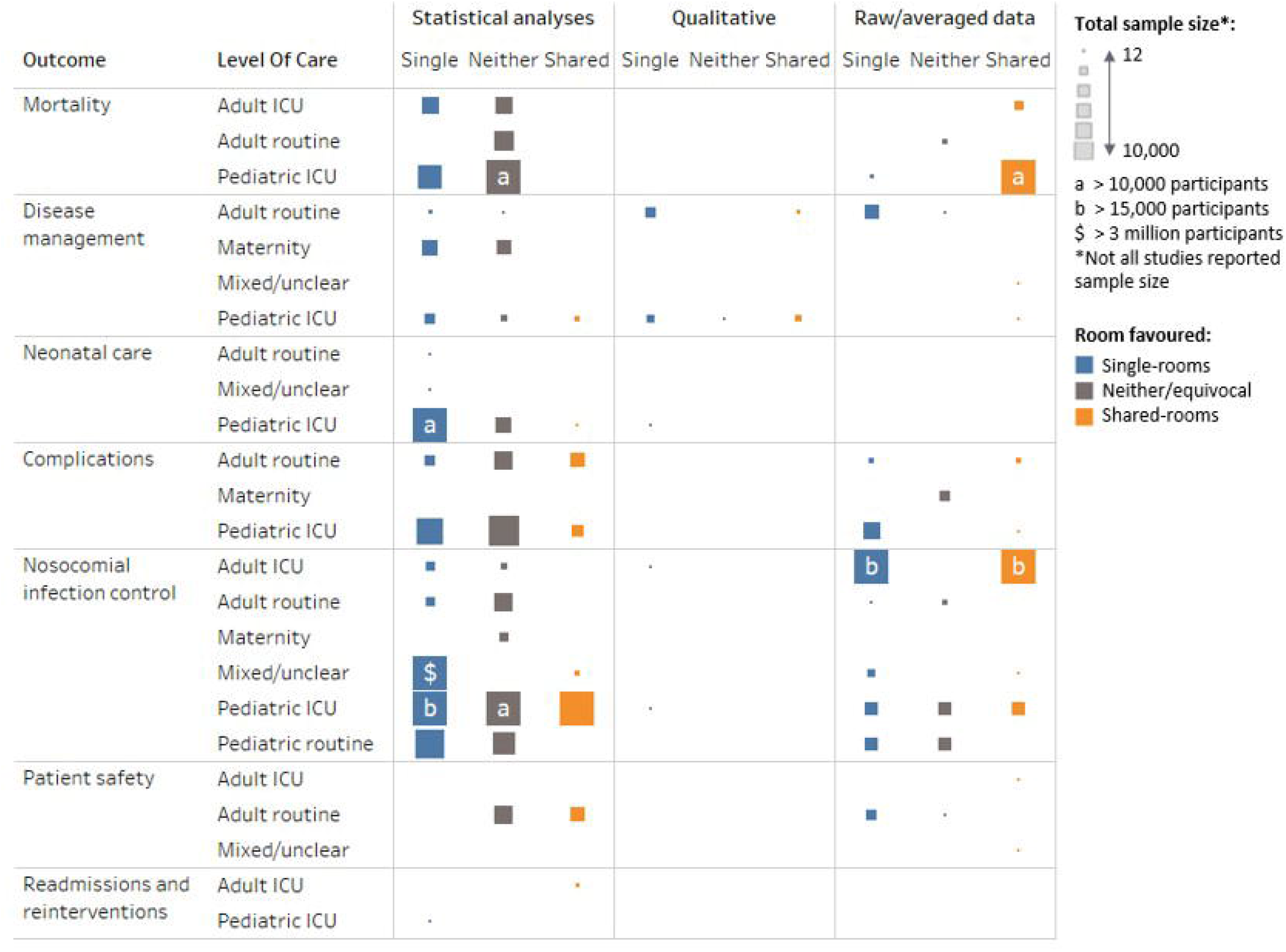
Clinical outcomes represented by the total sample size with data for that outcome, by level of care and the type of data reported and room design favoured.

#### Routine care

Four studies involved patients receiving routine care,(7,8,16,22), all in adults, none of which found a significant difference in mortality between those in single rooms versus shared accommodation, including up to 1 year after discharge.

#### Intensive care

Six studies assessed mortality among adults in ICUs.(12,13,17,18,20,24) One study by Bracco and colleagues(17) favoured single rooms. That study included 2,522 adults in ICUs in Canada and reported mortality of 2.9% among those in single rooms or cubicles compared with 8.3% among those in shared accommodation (p<0.001). A study of 666 adults in ICUs in Korea with COVID-19 favoured shared accommodation, reporting 2.4% mortality versus 4.6% among those treated in single rooms but no statistical analysis of the difference was reported.(20) The other studies showed no differences between accommodation types.

Seven studies assessed mortality specifically among neonates in ICU.(9–11,15,19,21,23) Three favoured single rooms. Lehtonen et al.(23) assessed 4,662 neonates in 331 neonatal ICUs (NICUs) across 10 countries (Canada, Australia, New Zealand, Finland, Israel, Japan, Spain, Sweden, Switzerland, Italy) and found that those cared for in units with single rooms had lower odds of death or any major morbidity than those in units with no such facilities (adjusted odds ratio 0.76; 95% CI 0.64–0.89). Two papers reported reduced mortality in single rooms among a small population of neonates in intensive care, but statistical significance was not reported.(9,10) By contrast, two studies favoured shared accommodation. Puumala and colleagues(15) reported a lower percentage of deaths among 9,995 neonates. Harris et al.(19,25) assessed NICUs in 11 hospitals in the USA and found fewer deaths among neonates nursed in units with shared accommodation compared with units with single rooms. However, statistical significance was not reported in either.

Lazar et al(14) was the only study to assess children in a paediatric ICU and found no difference between accommodations types.

### Patient care and disease management

Twelve publications reported on outcomes related to patient care and disease management (see Figure 3 and supplementary Table 3).(26,8,27–36) All were in adults or non-specified age groups. Three were before-and-after studies, four were contemporaneous studies, and five were evidence syntheses. Four studies had quality scores below 50% but four had scores greater than 75%. All papers assessed routine care.

Most findings favoured single rooms. Significance was shown for improvements in cleanliness,(30) pain management(30), and interactions between patients and medical staff,(31) and other findings were descriptive. A study in Australia of 1,569 orthopaedic patients had fewer emergency calls due to deterioration in condition after a move to single rooms compared with patients in shared accommodation.(8) As room allocation was based partly on severity of illness, nurses tended to position themselves nearer higher-risk patients to aid visualisation. Lawson and Phiri(27) found better patient satisfaction with care and lower analgesic use in orthopaedic patients in single rooms.

Three systematic reviews found that patients in single rooms may have faster recovery due to better sleep and a more pleasant environment,(33) but there was no consistent effect on use of medication.(35) An OECD WHO report concluded that single-room occupancy was associated with reduced pain scores, but due to a lack of detail had a very low quality score (14%).(37)

Findings in favour of shared accommodation were feelings of safety(28) and less use of restraints.(34) In a comparison of only single rooms after a move to a new hospital with only shared accommodation in the previous hospital, falls and medication errors in the medical assessment unit increased notably immediately but by 9 months had fallen to levels lower than previously.(26) However, in the single-room ward for older adults, falls and pressure ulcers significantly increased after the move and remained higher than before moving. No similar trends were seen after the move in a control hospital with 50% single rooms and 50% shared accommodation, and this was the preferred choice of nurses before and after the move (38% and 40%, respectively).

### Maternity and neonatal care

Twenty-three studies were found that assessed maternity and neonatal care (see Figure 3 and supplementary Table 4).(10,11,15,38–57) Most (n=14) were before-and after studies and the remainder were contemporaneous studies. Three studies were low quality and only one had a high-quality score. Many of these studies included statistically assessed findings, with most favouring single rooms or showing no difference between accommodation types.

#### Maternity care

Nine studies considered maternity care and perceptions of mothers and family members.(39,40,42,44,45,47,48,50,51)

Harris and colleagues(41) assessed 976 low-risk patients, 583 of whom received all care in single rooms and 393 in separate labour, delivery, and recovery areas. While overall use of intrapartum interventions was similar, maternal outcomes were better in single rooms. After discharge, Erdeve et al(48) reported that mothers of babies in NICUs who received care in shared accommodation had significantly more acute care visits (p=0.046), telephone consultations (p=0.01), and rehospitalizations (p<0.05) than those cared for in single rooms, and the reasons were more likely to be for issues related to prematurity like feeding difficulties compared with anatomical disorders. This perception is supported by the findings of Janssen et al,(42) which showed that mother in single rooms rated information and instructions at discharge as being clearer than those in shared accommodation.

Multiple studies indicated that satisfaction with care teams was greater in single rooms, including duration and quality of interactions and needs met.(39,40,42–45,47) In one study participants felt that parental presence was greater in single rooms than in shared accommodation.(50) In a US study, women reported less pain in single rooms than in shared accommodation.(43)

#### Neonatal care

Fourteen papers reported on outcomes in neonatal care.(10,15,38,41–43,46,49,52–57) Many of the results for neonates in ICUs showed no differences in outcomes between accommodation types. Significant improvements were seen in breastfeeding outcomes(10,15,55,58) and weight gain(46,58) in favour of single rooms. However, Tandberg et al(49) reported that longer-term weight gain (4 months) was better after neonatal care in shared accommodation. In two studies, reduced apnoea events were associated with single rooms,(10,59) and in another study less need for mechanical ventilation was reported in single rooms.(55) Significantly reduced neonatal pain scores were also reported.(43)

### Complications of disease

Twenty-three articles assessed disease complications (see Figure 3 and supplementary Table 5).(7,8,16–18,22–24,26,34,37,41,43,48,54,56,60–64) Nine were before-and-after studies, 10 were contemporaneous studies, one used a mix of study designs, and three were evidence syntheses. Two articles had quality scores below 50% and two had scores greater than 75%. Findings generally favoured single rooms or showed no differences between accommodation types.

#### Routine care

Eight papers assessed complications specifically in routine care and all assessed care of adults.(7,10,16,22,41,63–65) Only one study reported results with significance assessed, which showed reduced incidence of delirium among older adults with dementia nursed in single rooms (hazard ratio 0.66, 95% CI 0.48–0.93, p<0.02).(65) The Scottish guidelines on delirium recommend reducing light and noise and having familiar items around patients with or at risk of developing delirium,(64) which might be supported by this finding.

In a small relocation study (n=64)(26) pressure injuries seemed to be increased around tenfold in single rooms and falls in 50% or 100% shared accommodation, but a substantial change in case mix made this finding difficult to interpret. By contrast, in a larger non-controlled UK relocation study (n=1,569), no significant difference was noted between different types of accommodation.

The findings for other complications, such as hip fracture rates following falls, thromboembolic events, infections, and other medical complications were not significantly different among orthopaedic patients in single rooms compared with those in shared accommodation in an Australian study.(42) Patients in single rooms were more likely to be female and much more likely to have private health insurance, which may have biased the outcomes.

#### Intensive care

Twelve papers specified assessments in ICU settings, of which three assessed adults(17,18,24) and nine concerned neonatal care.(23,43,48,54,56,60,60–62) In one study of 1,253 adults in Brazil,(18) delirium was significantly less likely among those in ICU single rooms than in shared accommodation, but no significant difference was seen between groups of elderly patients in different types of ICU accommodation in the Netherlands.(24) Organ failure was reported to be significantly lower in patients managed in single rooms in one study.(17) However, few data are available in adults and most studies reported no differences between accommodation types.

An international study of 4,662 preterm neonates found a significantly lower risk of death or any major morbidity, including sepsis and retinopathy of prematurity, among those nursed in NICUs with single family rooms (odds ratio 0.76, 95% CI 0.64-0.89).(23) In contrast, another study showed lower rates of necrotising enterocolitis and intraventricular haemorrhage in shared accommodation.(46) However, in other studies, rates of these and other serious complications were similar in all ward types.(46,56,59,60) Lester et al(60) found that neonatal stress levels were reduced among babies in NICU single maternity care rooms compared with those in shared accommodation.

### Prevention of infection

Fifty-one studies discussed prevention of infection (see Figure 3 and supplementary Table 6).(8,10,11,13–15,17–19,21,26,34–36,46,49,53,56,61,66–98) Twenty were before-and-after studies, 28 were contemporaneous studies, one used a mix of study designs, and three were evidence syntheses. Seven had low quality scores and nine had high quality scores. More than half (n=33) studies reported statistically analysed data.

#### Routine care

Routine care was assessed in adults in 10 studies(8,69,75,77,79,80,84,87,89,93) and eight involved mixed age populations and care levels that stated or were assumed to include adults and routine care.(26,35,36,67,71,72,88,95) Hospital-acquired infection rates were shown to be reduced in single rooms in six studies.(26,36,67,69,71,87,88) However, in Maben et al,(26) this finding depended on the ward mix: *Clostridium difficile* infections were reduced in single rooms where the split with shared accommodation was half and half, whereas all shared accommodation performed better than all single rooms. In Darley et al(67) this finding was only for *C difficile*, whereas hospital-acquired MRSA rates did not differ by accommodation type. Bocquet et al(78) and Munier-Marion et al(87) found reduced nosocomial influenza infections in single rooms and one study showed a reduced risk of norovirus infection.(82) By contrast, McDonald and colleagues(71) noted reduced infection rates for *Enterococcus* spp, *C difficile*, and MRSA. In a systematic review, Voigt et al(35) concluded that the quality of evidence did not support the use of single rooms over shared accommodation. Indeed, only two studies showed increased infections in shared accommodation.(26,83) Nevertheless, in one study patients(77) preferred single rooms for infection prevention. In a study of more than 1 million patients of all ages across 2018 hospitals in the USA, O’Neill and colleagues(95) found that single rooms were significantly associated with reductions in central-line-associated bloodstream infections.

Seven studies assessed routine care in children.(78,81,82,86,90,96,97) Two found a decrease in nosocomial infections in single rooms ⍰ one overall(78) and one for diarrhoea in gastrointestinal and neurosurgical units,(81) but the latter found no difference between accommodation types in a cardiological unit. In two large studies in Finland (n=1,927 and n=5,119), Kinnula and colleagues(96,97) saw increases in hospital-acquired infections among children admitted to shared accommodation in an infectious disease ward because there was no grouping by aetiology. All hospital-acquired infections with symptoms during the hospital stay and 49% of those manifesting after discharge led to diarrhoea. The risk of infection was doubled among children sharing accommodation with patients who had respiratory infections (OR 2.3, 95% CI: 1.1–4.8; p=0.03). Risk decreased per year of age. Among 83,334 children assessed in two hospitals, Quach et al(90) found significantly increased rates of respiratory infections when accommodation was more than 50% single rooms (rate per 1,000 patient-days 1.33, 95% CI 1.29–1.37).

#### Intensive care

Outcomes in ICUs were reported specifically in 26 studies, 11 in adults and mixed-age populations,(13,17,18,66,68,70,73,85,91,92,98) one in children,(14) and 14 in neonates.(10,11,15,19,21,46,49,53,56,61,72,74,76,94)

Among adult populations, only one study showed outcomes in favour of shared accommodation, with reductions per 10,000 patient-days in cultures positive for *Enterobacter* spp, *Haemophilus, Streptococcus viridans, Acinetobacter* spp, *Streptococcus pneumoniae*, Group B *Streptococcus* spp, *Neisseria* spp, and MRSA.(73) However, in the same study, single rooms showed lower rates of infections with many common organisms, such as *Staphylococcus* spp, *C difficile*, and *Pseudomonas* spp. Four studies showed significant data on reduced bacterial infection and transmission in single rooms based on isolates and antibiotic use,(17,66,70,91) although in the study by Halaby et al,(70) transmission of *Morganella* spp, *Proteus* spp, *Serratia* spp, and *Pseudomonas* spp did not differ between accommodation types. Two studies indicated reduced risks of bloodstream infections in single rooms.(17,92) As for routine care, patients perceived infection prevention to be better in single rooms than in share spaces.(68)

Among neonates and among children in ICUs, the findings were mixed. In favour of shared accommodation, four studies reported reduced cases of nosocomial sepsis,(10,15) one reduced colonisation with multidrug-resistant organisms,(46) and one nosocomial infections with pneumonia.(19) Four studies indicated no difference between accommodation types for sepsis or septicaemia and/or found that the use of single rooms was associated with fewer sepsis cases.(15,49,56,74) Only one study showed an increase in sepsis in shared accommodation, and that was specifically in neonates born at or after term.(15)

### Patient safety

11 studies considered patient safety (see Figure 3 and supplementary Table 7).(8,16,22,26,34–36,65,84,99,100). Three were before-and-after studies, four were contemporaneous studies, one used mixed design, and three were evidence syntheses. Two had low quality scores and three had high quality scores. Most of the studies assessed routine care or mixed care populations, and generally the populations were adults and elderly people.

Overall, the data showed no differences between accommodation types or favoured shared accommodation. Only the OECD study indicated reduced risk of falls in single rooms,(36) but the quality of this study was deemed to be very low due to reporting very few details of the research. Significantly lower rates of falls were seen in multi-bed accommodation in two studies.(16,22) The study of Poncette et al,(100) which analysed alarm data in an ICU, found that the number of alarms per bed per day was higher in single rooms than in shared accommodation.

### Readmissions and reinterventions

Only two studies reported on readmissions and reinterventions (see Figure 3 and supplementary Table 8). They were both contemporaneous studies and one had a quality score of 74% and one of 78%. One showed that single rooms were associated with lower rates of rehospitalisation.(48) The other favoured shared accommodation, with fewer patients returning to theatre within 6 weeks of treatment.(63)

### Privacy

Forty-eight publications, including six evidence syntheses, reported on privacy (see supplementary table 9).(8,9,19,26,28,30–34,36,38–42,44,47,51,58,59,68,77,86,98,99,101–122) Eighteen were before-and-after studies, 23 were contemporaneous studies, one used mixed designs, and six were evidence syntheses. Nine had low quality scores but 19 had high quality scores. They were mainly descriptive studies but overwhelmingly favoured single rooms.

#### Routine care

Twenty-eight studies assessed privacy among adults receiving routine care,(8,26,28,30,31,33,34,36,39,41,42,58,77,99,101,102,105,108,109,112–119) with seven of these reporting statistical analyses.(26,28,30,36,41,42,105,114) Key aspects of privacy in single rooms were improved confidentiality when discussing personal information, use of private bathrooms, and privacy during early post-partum care (e.g., assistance with feeding). However, in the study by Florey and colleagues,(105) 83% of patients in shared rooms also reported feeling that they had adequate privacy. Likewise, the systematic reviews by Taylor and colleagues(34) and Dowdeswell and colleagues(32) found advantages and disadvantages with regards to privacy in all studies they assessed. Patients reported feeling as though they could ask more questions or make more remarks in single rooms than in shared accommodation, and more scored physicians’ responses as being empathetic.(31) Qualitative or descriptive studies also strongly supported greater privacy in single rooms.(8,77,99,101,102,108,109,112,115–118,121)

Four studies assessed routine care among children.(86,104,111,120) Boztepe et al(111) found that children did not ranked privacy highly and were more concerned about procedures being painful. In this study many of the children had extensive history of hospitalisation. The other three studies reported greater privacy in single rooms, but children also seemed to enjoy the social aspect of shared accommodation. The main reasons for preferring single rooms were private bathrooms and the capacity for family members to stay. Sleep was an important aspect of care in single rooms for children and parents. (58,86,120)

#### Intensive care

Nine studies reported on privacy for adults in ICUs, and generally the findings favoured single rooms.(32,34,36,40,44,47,59,68,110) Three studies reported statistical evidence of improved privacy in single rooms among adult patients.(36,40,44) The literature reviews by Dowdeswell and colleagues(32) and Taylor and colleagues(34) showed mixed findings among studies.

Eleven studies addressed neonatal care in ICUs.(9,19,38,40,44,47,51,98,103,106,107) All but two(38,51) favoured single rooms for privacy.

### Loneliness/isolation and family contact

Fifty-five publications, five were evidence syntheses, reported patients’ views about loneliness or family contact associated with single-room accommodation (see Figure 4 and supplementary Table 10).(9,16,19,25,26,28,30,32,33,36,38,39,42,44,45,47–49,49,50,54,57–59,68,74,77,86,98,99,102,104–110,114,115,117–120,123–132). Twenty were before-and-after studies, 29 were contemporaneous studies, one used mixed study designs, and five were evidence syntheses. Only nine had quality scores less than 50%, while high quality scores were assigned to 17.

**Figure 4.**
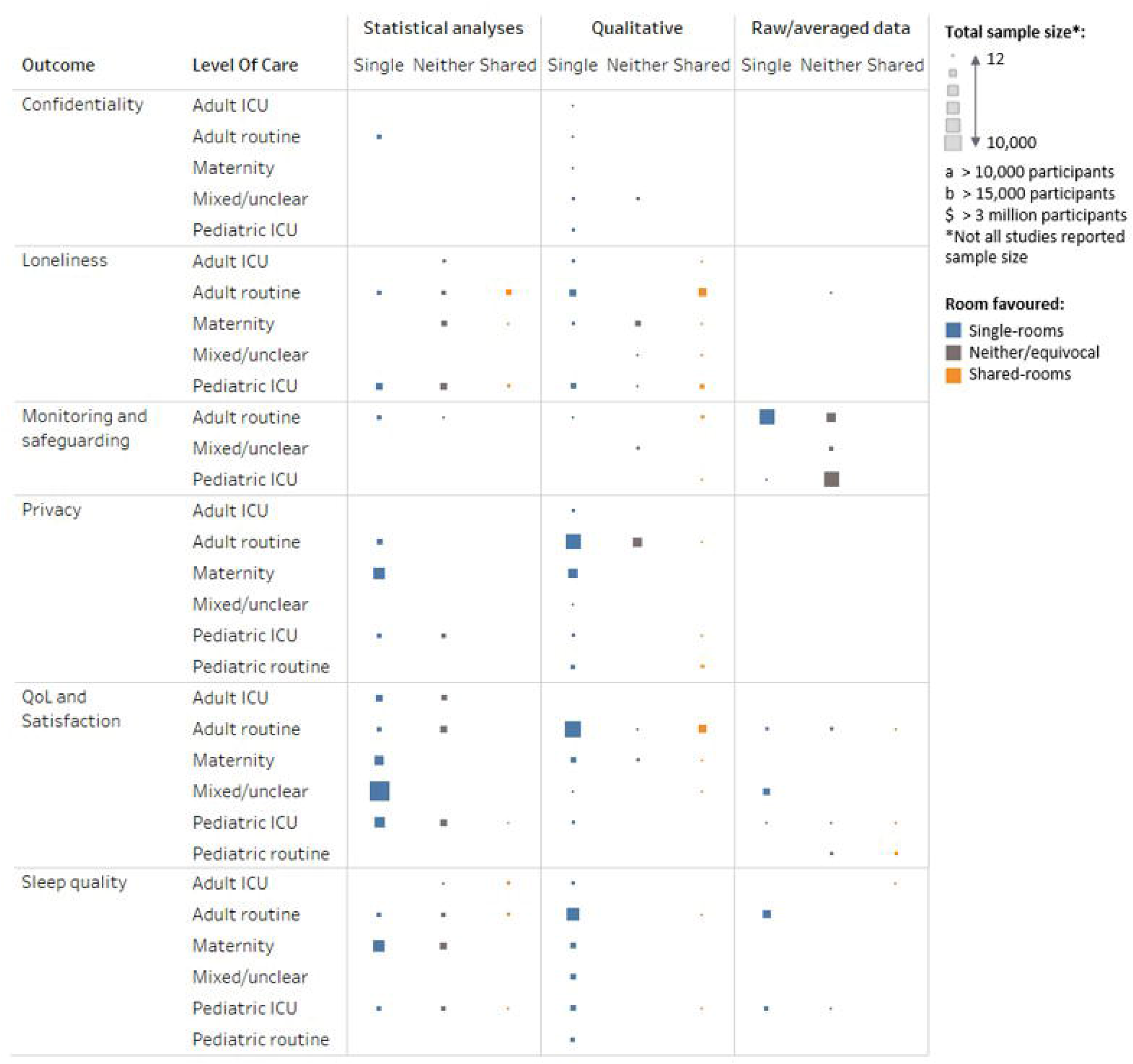
Patient-experience outcomes represented by the total sample size with data for that outcome, by level of care and the type of data reported and room design favoured.

Two main themes seemed to be revealed in patients’ perspectives: shared accommodation were strongly preferred for social interaction to avoid loneliness/isolation,(9,16,26,28,33,38,47,58,68,77,86,102,104,105,109,114–120,123,124,126,129) whereas single rooms were preferred for privacy (e.g., for bathroom use, during consultations and visits, and to spend with children, particularly neonates).(9,28,32,38,45,47,49,49,50,54,57,59,68,74,77,86,98,104–106,108–110,114,127,130,132)

### Noise, disturbance, and sleep

Forty-five publications, four of which were evidence syntheses, reported patients’ views about noise, disturbance, and sleep associated with single-room accommodation (see Figure 4 and supplementary Table 11).(8,9,24–26,28,30,32,33,36,38–42,47,54,58,59,68,76,77,86,103,105,107,108,110,112–114,117–119,131,133–141) Sixteen were before-and-after studies, 24 were contemporaneous studies, one used mixed study designs, and four were evidence syntheses. Nine had quality scores less than 50% and 13 had high quality scores.

In general, patients felt that single rooms were quieter and led to less sleep disruption. However, measurement of noise levels showed that there were no substantial objective differences between single rooms and shared accommodation.(30,54,135) One study reported lower noise levels in single rooms, but the difference was not statistically assessed.(76) Stevens et al(54) and Meyer et al(139) found that respiratory support and other medical devices could raise noise levels in single rooms enough to disturb sleep even when ambient noise had been reduced. HCPs also reported that single rooms improved patients’ sleep.(9) Poncette and colleagues(100) found that fewer alarms raised in shared accommodation reduced overall noise levels. One study also noted that patients preferred single rooms because they felt they were less likely to disturb other patients.(119) Most studies addressing sleep found that it was improved in single rooms. (28,30,32,33,36,58,59,86,113,114,117,118,136,140) This was generally due to fewer disturbances and/or a perceived quieter environment that in shared accommodation. Hosseini and colleagues(114) and Sakr and colleagues(140) noted that the risk of new-onset insomnia was significantly higher among patients in shared accommodation (95.7% vs 75%, p=0.011).

Humidity and temperature were discussed in one article. Van Enk and colleagues(135) reported that in a NICU with centrally controlled humidity, shared accommodation had non-significantly lower percentage of relative humidity than single rooms but showed much greater variance (26.8% (±17.0) in single rooms vs 26.0% (±89.0)). Both mean values were lower than the recommended range for NICU (30160%). Temperature could be controlled within individual single rooms and mean values were significantly lower than those in the shared accommodation, which had central temperature control per nursery (mean 73.8°F [range 65.3–77.5°F] in single rooms vs 76.0°F [range 71.1184.5°F], p=0.0001). More than 85% of readings in both, though, were within the recommended range of 72— 78°F, although readings outside the range were too hot in shared accommodation and too cold in single rooms. The authors suggested that thermostats should be allowed to vary only within the recommended range.

Lighting was assessed in three studies, two of which favoured single rooms due to less illumination for neonates (40,54) and one study of patients with delirium that favoured lower light in a shared accommodation.(24)

### Satisfaction with care

Fifty-one publications, six of which were evidence syntheses, reported patients’ satisfaction with care (see Figure 4 and supplementary Table 12).(8,12,19,25,27,28,30,32,33,35,36,38–40,42,44,49,50,57,58,60,62,74,77,86,99,101,103,105,107–109,111–114,116–119,121,125,128,129,131,133,136,142–146) Fifteen were before-and-after studies, 29 were contemporaneous studies, one used mixed study designs, one was an economic analysis and six were evidence syntheses. The quality scores of ten studies were less than 50% and of 14 were 75⍰100%.

Overall, results show either little difference between accommodation types or results in favour of single rooms. Single rooms seemed to be favoured most by mothers in maternity units, whereas preference towards shared accommodation seemed to increase with rising age. The economic analysis found that patients were willing to pay for private care to have single rooms.(146)

#### Routine care

Routine care was assessed in 30 studies (see supplementary table 12).(8,27,28,30,32,33,35,36,39,42,58,77,86,99,101,105,111–114,116–118,121,129,131,133,136,143,144) Patients preferred shared accommodation in seven studies.(33,86,116–118,129,133) Generally they preferred interaction with other patients, and in two reports they stated that they found the shared accommodation more secure and safe.(117,118) Specific reasons given for preferring single rooms were privacy,(30,58,77,99,112,131,133) comfort/environment,(27,42,101,108,143), level of care and information, effect on recovery,(32,36,42,101,113,114,136) and safety.(30,35)

#### Intensive care

We found 24 reports of intensive care(12,19,25,32,36,38,40,44,50,57–60,62,74,103,107,108,125,128,130,131,143,145). Only two reported findings that favoured shared accommodation. Campbell-Yeo et al(50) found that in an open-bay NICU, mothers reported better self-efficacy and less uncertainty about their babies’ health. Also in a NICU, Pineda and colleagues(57) found that the risk of stress among mothers was significantly lower in shared accommodation than in single rooms, although life stress did not differ between accommodation types. By contrast, other assessments of stress found that risk was reduced in single rooms(19,25,60,74,130) or did not differ.(50,62,130) Satisfaction with design/environment, where assessed, favoured single rooms.(59,108)

Findings on satisfaction with maternity care was greater for parents in single rooms in three studies(12,59,60) but did not differ between accommodation types in two.(74,145) Single rooms seemed to have little effect on postpartum depression or irritability, most measures not differing between accommodation types(57,62,74,130,145) and only four findings favouring single rooms.(19,25,50,130)

Only eight of the 51 studies were related to satisfaction with care in other patient populations, involving cardiovascular, cancer, adolescent, or mixed adult care.(12,32,36,58,108,128,131,143) All these studies’ findings supported single rooms.

### Patient monitoring and safeguarding

Although the impact of single rooms on healthcare staff was not the focus of this review, 14 of the included publications reported the views of HCPs as well as patient-reported outcomes that we used to explore monitoring of patients (see Figure 4 and supplementary Table 13).(11,13,17,21,28,30,47,68,77,106,108,112,114,133) Four were before-and-after studies, eight were contemporaneous studies, one used mixed study designs, and one was an evidence synthesis. No study had a low-quality score. Five of the studies were classified as being of high quality.

Most of the studies presented descriptive/qualitative findings. Three studies reported statistically assessed data, all in relation to routine care and among adult patients. Two(28,114) favoured single rooms, reporting that availability to patients, meeting patients’ needs, and access to patients were improved. The fourth study showed no difference between single rooms and shared accommodation for responding to patients’ call alerts. One study reported that nurses felt they might spend longer with patients in single rooms, depriving other patients of as much care.(106) In another, safety of patients in units with single rooms was raised as an issue due to increased distances between nurses and patients and impeded observation of patients.(68)

### Patient confidentiality

Confidentiality was assessed in 11 studies (see Figure 4 and supplementary Table 14).(26,36,47,68,77,105,106,109,114,115) Five were before-and-after studies, four were contemporaneous studies, one used mixed study designs, and one was an evidence synthesis. Two studies had low quality scores while four had high quality scores.

All studies concluded that patient confidentiality was better maintained when patients were in single rooms, with one study finding no difference for adults with cardiovascular disease in ICU(108) (see supplementary table 14). Malcom et al(115) found that the lack of privacy and confidentiality in shared accommodation affected patients’ relationships with other patients.

### Availability of beds, space requirements, and capital costs

Sixteen studies reported on beds, space, and costs associated with different accommodation types (see Figure 5 and supplementary Table 15).(10,12,13,21,26,27,38,57,67,90,96,97,106,108,126,147) Nine were before-and-after studies, six were contemporaneous studies, and one used mixed study designs. Two studies had quality scores below 50% and three were classified as being of high quality.

**Figure 5.**
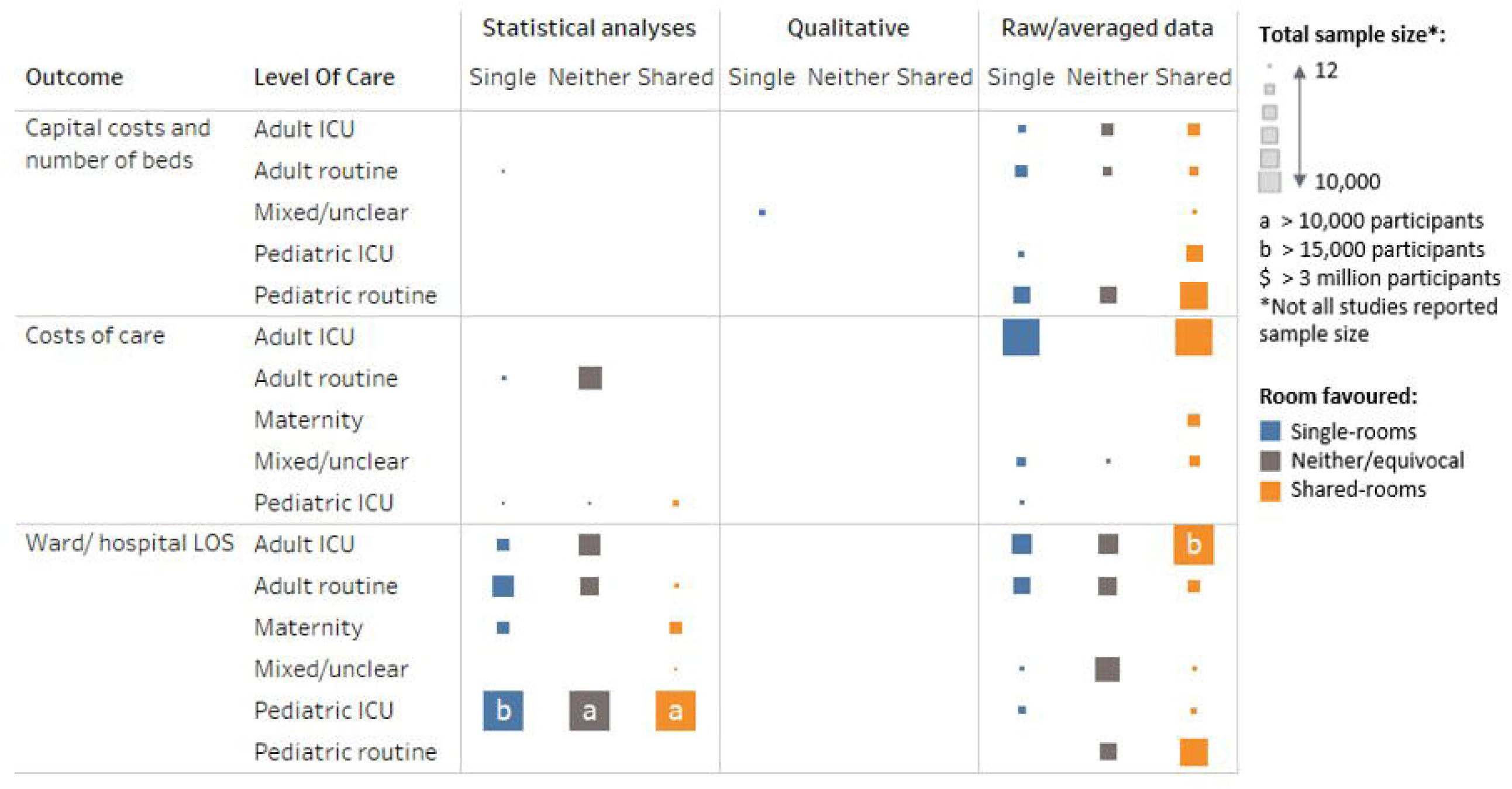
Economic outcomes represented by the total sample size with data for that outcome, by level of care and the type of data reported and room design favoured.

There did not seem to be strong evidence in favour of either accommodation type. The inclusion of single rooms substantially increases the amount of floor space required to achieve the same number of beds as in shared accommodation, with estimates suggesting between 30% and 50% more floor space being required per bed, which increases capital costs.(10,12,26,27,106,126,147) Shared accommodation provides greater flexibility to add beds in times of need.(13,21,57,90,97) Darley and colleagues(67) found that numbers of bed-days lost due to ward closures caused by norovirus outbreaks was greatly reduced after moving to a hospital with 75% single rooms from the previous 10% single rooms.

### Length of stay

Fifty-three publications, including two evidence syntheses, reported on length of stay associated with single-room accommodation (see Figure 5 and supplementary Table 16).(7–13,15–20,22–27,35,36,40,41,43,46–49,52,53,55,56,60–63,65,69,73–75,85,91,96,96,107,112,113,125,134,143,147–149) Twenty-eight were before-and-after studies, 23 were contemporaneous studies, one used mixed study designs, and two were evidence syntheses. Five of the studies were classified as being of low quality but only seven fell into the high-quality category. The evidence was highly mixed with no clear benefit from either accommodation type.

#### Routine care

Of 20 studies assessing routine care, 18 concerned adults and the elderly.(7,8,22,26,27,35,36,41,63,65,69,77,112,113,134,143,147,149) Among these, eight found that length of stay was shorter in single rooms,(7,16,26,27,41,65,112,147,149) but in the study by Maben et al(26) this was true only for an older people’s ward and not for a medical assessment unit, and in that by Lawson and Phiri,(27) while it was true for non-surgical orthopaedic patients and psychiatric patients, no difference was seen for surgical orthopaedic patients. One study found that length of stay was shorter in shared accommodation among older patients with dementia, overall and among those who had experienced inpatient falls.(22) No difference in overall length of stay was reported in seven studies, including two evidence syntheses.(8,35,36,63,69,134,143)

The two studies of routine care in children by Kinnula and colleagues showed no difference between accommodation types in one(96) but longer duration of admissions among children in shared accommodation in another.(97)

#### Intensive care

Of 32 studies assessing length of stay in intensive care, 23 considered neonates(9–11,15,19,23,40,43,46,47,47,48,52,53,55,56,60–62,74,91,107,125,130,148) and nine adults.(12,13,17,18,20,24,73,75,85) As for routine care, the results were highly mixed.

Among the nine studies assessing care of adults in ICUs, five showed no significant differences between single rooms and shared accommodation.(12,13,24,24,75) Teltsch et al(73) assessed care after a change from multi-bed to single rooms and compared the findings to a hospital with no change. The length of stay in the ICU in the comparator hospital increased year on year from 3.8 days to 4.2 days from 2000 to 2005. While higher after the change to single rooms, the length of stay did not change substantially over the same period, and after adjustment was an estimated 10% lower overall (relative ratio 0.90, 95% CI 0119%). Bracco and colleagues(17) reported that patients were able to stay longer in the same bed in single rooms during infection outbreaks in an ICU, although overall LOS was not significantly different.

In NICUs, statistically significant shorter durations of stay in hospital were reported in three studies. Puumala and colleagues(15) found that stays were shorter for very and extremely preterm babies, but there was no difference between accommodation types for moderately preterm babies, and stays for term and post-term babies were shorter in shared accommodation. Lehtonen et al(23) found that stays were on average 3.4 days shorter (95% CI 3.114.7) and van Veenendaal et al(74) found a median difference of 2 days in favour of single rooms. Qualitative/descriptive studies also favoured single rooms in five studies.(9,40,47,107) Three studies identified shorter stays in shared accommodation,(19,56,148) but 13 found no difference between accommodation types(10,11,43,46,48,52,53,55,60–62,125,130).

### Costs of care

Nineteen publications, including two evidence syntheses, reported on costs or resource use associated with different types of accommodation (see Figure 5 and supplementary Table 17).(8,16,19,22,25,26,35,41,54,63,91,107,110,132,133,146,148,150) Eight were before-and-after studies, seven were contemporaneous studies, two used mixed study designs, and two were evidence syntheses. Four studies had low quality scores and six had high quality scores.

Several studies reported multiple measures of costs and found evidence supporting both types of accommodation. Therefore, the evidence split was 10 studies finding in favour of single rooms, 10 in favour of shared accommodation and six showing no difference in measures.

Boardman and Forbes recommended taking into account the following construction and running costs, given that single-room facilities required more space to construct: land costs, construction costs, maintenance (refinishing and updating), housekeeping and operating costs, and health care provision (potentially longer distances to cover). Maben et al(26,133) estimated in 2015 that the cost of building a hospital solely comprising single rooms could be around 5% more than building one with predominantly shared accommodation but suggested that the difference becomes marginal over time. Harris and colleagues(25) found that the most cost-effective configuration in terms of construction costs per square foot was a combination of open bays and single rooms.

Findings in favour of single rooms were due to reduced overall staffing costs,(41,150) reduced length of stay,(91,107) reduced waiting and transfer times,(146) higher proportions of patients being discharged to rehabilitation,(63) reduced infections,(91) and operational efficiencies.(35,148) Reasons favouring shared accommodation were lower cleaning and housekeeping costs,(26,133) perceived increased nursing staff,(26,148) and lower labour costs in NICUs.(148)

## DISCUSSION

This systematic review identified a substantial body of evidence associated with hospital accommodation, yet no clearly consistent conclusions could be drawn about overall benefits of single rooms versus multi-bed ward spaces. The narrative and heterogeneous nature of much of the evidence also meant that a formal statistical synthesis, such as a meta-analysis, was not feasible. Nevertheless, some themes did emerge and might be worth considering further. Single rooms were most likely to be associated with overall clinical benefit for the most severely ill patients, especially neonates in intensive care, although the evidence is mixed even in these high-risk populations. Patients who preferred single rooms tended to do so for privacy, particularly having a private bathroom, and for reduced disturbances. By contrast, there were distinct patterns of men, older adults, children, and adolescents being more likely to prefer shared accommodation, particularly for the social aspects. While mixed accommodation types seemed to be the most cost-effective approach to construction because the capital cost of single room building is higher than that for shared accommodation, the running costs seem likely to be recouped over time by other efficiencies.

While patients and HCPs expressed preferences, health-care outcomes seem unlikely to be substantially affected by hospital accommodation. This is reassuring because most patients also have little influence over this aspect of their care. Likewise, HCPs might also have little influence over which accommodation type their patients are assigned. Patient or family preferences for single rooms are particularly strong in NICUs and maternity wards, but other groups, in particular men, older adults and adolescents, are more likely to prefer shared accommodation. The split of accommodation and whether the predictable adverse effects of accommodation design can be mitigated in these areas would be worth considering at the planning stages of new buildings.

The average cost per patient of units comprising only single rooms was lower than those consisting of only shared accommodation. Mean direct cost per patient in a single room has been estimated to be 15.5% lower for neonates in ICU and 24% lower for care in maternity units but may be similar or reduced in adults in routine care wards. Shorter length of stay was an important contributor to this difference and could increase the number of patients who can be treated in the beds available. However, the effects found were small and local variations may change the economic picture for a particular hospital. It is also unclear how far the reduced length of stay was due to the single room, and how much was caused by confounding factors associated with being in a new hospital. Nearly all the studies we considered were from high-income regions and mostly based in European or anglophone countries. Thus, policy makers should incorporate local building and labour costs in decisions.

Determining the impact of moving to single-room wards will therefore always need to overcome the impact of confounding factors such as concomitant changes to processes and improvements in other facilities and services that may also have led to the changes, or that may have acted in opposition to the direct effects of the different accommodation.

This study has some limitations. Of 215 articles originally retrieved, we selected 145 for review, which is still a large number. None of the 145 studies used randomised study designs. Randomised controlled studies are not practical to assess hospital accommodation and, therefore, most studies were prone to bias. In particular, in hospitals with mixed single rooms and shared accommodation, patients must be allocated to the rooms, which will be partly due to their medical condition or other personal factors and partly due to bed availability at the time of admission. This introduces selection bias, as the reasons why patients were in single rooms was often not reported. We minimised this bias by excluding studies where the allocation to single rooms or shared accommodation was known to be for an apparent clinical purpose. Additionally, 60 of the publications compared outcomes before and after moving from shared accommodation to single rooms. This introduced confounding, as many factors other than the studied intervention would have changed at the same time, so attributing the outcome to the intervention alone is misleading. The opportunity of building a new hospital is rare. While it provides an ideal platform for before-and-after studies, nearly all of these studies were of a single hospital, so the total number of hospitals studied is low, meaning that uncertainty about risks and benefits remains after this systematic review.

In practical terms, although there are still uncertainties about the true impact of changing to single room wards, the 145 publications synthesised in this report show that the clinical and economic consequences of such a change are likely to be modest. Focussed research on the impact of accommodation type on hospital acquired airborne transmitted infections may be warranted, as may studies of sufficient duration to examine long term productivity changes after opening a new hospital. Because the global effect sizes found were modest, the effects within national systems or for specialist hospitals might be different in size and direction from the global. In particular, none of the included studies were in low or middle income countries, so generalisability to those settings is difficult.

## Supporting information

Search Strategy

## Data Availability

All data produced in the present work are contained in the manuscript or supplementary material.

## ACKNOWLEDGMENTS

All authors wrote sections of the final manuscript. DMcP conceived the study and defined the research question. AM defined the detailed method and arbitrated disagreements. AB and NC conducted the literature screening and data extraction. CS developed the figures and summary tables. The corresponding author attests that all listed authors meet authorship criteria and that no others meeting the criteria have been omitted.

The Corresponding Author has the right to grant on behalf of all authors and does grant on behalf of all authors, a worldwide licence to the Publishers and its licensees in perpetuity, in all forms, formats and media (whether known now or created in the future), to i) publish, reproduce, distribute, display and store the Contribution, ii) translate the Contribution into other languages, create adaptations, reprints, include within collections and create summaries, extracts and/or, abstracts of the Contribution, iii) create any other derivative work(s) based on the Contribution, iv) to exploit all subsidiary rights in the Contribution, v) the inclusion of electronic links from the Contribution to third party material where-ever it may be located; and, vi) licence any third party to do any or all of the above.

The lead author affirms that the manuscript is an honest, accurate, and transparent account of the study being reported; that no important aspects of the study have been omitted; and that any discrepancies from the study as originally planned (and, if relevant, registered) have been explained.

## REGISTRATION

PROSPERO registration number CRD42022311689, registered on 2^nd^ March 2022, after preliminary searches were undertaken and before formal screening began.

