## Supplementary material for "The clinical, humanistic, and economic outcomes, including experiencing of patient safety events, associated with admitting patients to single rooms compared with shared accommodation for acute hospital admissions. A narrative synthesis systematic literature review": Search Strategy

### Appendix: Search Strategy

### 1. Searches

We ran systematic searches of Medline via PubMed and Embase via embase.com and supplemented these databases with additional searches of Google Scholar and the National Institute for Health and Care Excellence website using the search strategy below.

Embase search strategy

|  | Search terms | Number of hits (17 Feb 2022) |
| --- | --- | --- |
| 1 | 'hospital design'/exp OR 'hospital design' OR 'hospital management'/exp OR 'hospital management' OR 'health care facility'/exp OR 'health care facility' | 2,538,404 |
| 2 | (single OR multi*) NEAR/2 (room* OR bay* OR bed* OR accommodation) | 6,553 |
| 3 | 1 AND 2 | 2,005 |

PubMed search strategy

|  | Search terms | Number of hits (17 Feb 2022) |
| --- | --- | --- |
| 1 | **"Hospital Design and Construction"[Mesh] OR "Hospital Administration"[Mesh] OR "Health Facility Environment"[Mesh]** | 289,082 |
| 2 | **((single OR multiple) AND (room OR room* OR bay OR bays OR bed OR beds OR accommodation))** | 70,205 |
| 3 | 1 AND 2 | 2,599 |

Google scholar search strategy

|  | Search terms | Number of hits (14 Feb 2022) |
| --- | --- | --- |
| 1 | **Hospital AND ("single room" OR "multiple room" OR "single bed" OR "multiple bay")** | 42 |

NICE website

|  | Search terms | Number of hits (14 Feb 2022) |
| --- | --- | --- |
| 1 | **"single room"** | 215 |

We have used NICE’s online Evidence Search tool ([https://www.evidence.nhs.uk](https://www.evidence.nhs.uk/)).

This search strategy yields 4,563 abstracts for screening after deduplication, once the additional sources have been included.

### 2. Inclusion criteria

The inclusion criteria for the review are set out below.

| Criterion | Inclusion criterion | Exclusion criterion |
| --- | --- | --- |
| Population | Adults, adolescents or children who are inpatients in an acute hospital for more than 24 hours  Includes:  Those requiring level 0 or 1 care  Neonates, children and young adults (everyone under age 18)  Pregnant/labouring women  Vulnerable adults (those requiring assistance while living in the community)  Those with dementia or delirium  Those requiring level 2 or 3 care  Those primarily receiving rehabilitation  Those admitted for elective care  Those admitted for emergency care | Patients admitted to long-stay, community or other non-acute hospitals  Patients admitted as day cases to an acute hospital  Patients attending Accident and Emergency or medical assessment units but not admitted to an acute hospital |
| Interventions | Staying in a single room for the whole admission | Patients who are relocated to a single room during their admission (e.g. for isolation due to contracting infectious disease or for terminal care) |
| Comparison | Staying in a multi-bed bay or room | No direct comparison |
| Outcomes | In-hospital mortality  30-day, or longer, mortality  Morbidity such as falls, deterioration, new pressure ulcers and additional diagnoses  Experience of patient safety incidents  Hospital-acquired infection  Quantitative, qualitative and patient-reported measures of experience  Length of stay  Cost of stay  Experience of accommodation change during admission, or number of bed moves during admission  Impact on caregivers and family of dependent patient | Non-clinically relevant outcomes  Impact on healthcare professionals and support staff |
| Study methodology | Comparative clinical trials and observational studies  Systematic reviews of relevant studies  Study protocols: index separately | Non-comparative observational studies  Narrative reviews, opinion pieces, letters, editorials  Conference abstracts with no relevant data |
| Study size | Any |  |
| Language and location | Any |  |

### 3. Studies excluded at full text screening

| Citation | Reason for exclusion |
| --- | --- |
| Adabi, G. *et al.* (2019). Barriers to sleep in patients hospitalised with an acute exacerbation of chronic obstructive pulmonary disease. *J Sleep Res.* 28(SUPPL 1). | Outcome: No relevant data reported |
| Alzheimer's Disease International (2020) World Alzheimer Report 2020 Design Dignity Dementia: dementia-related design and the built environment, Volume 1 https://www.alzint.org/u/WorldAlzheimerReport2020Vol1.pdf | Outcome: No relevant data reported |
| Baillie, J. (2015). Hospitals single-room design evaluated. *Health Estate* 69(1): 27-30. | Irretrievable |
| Bernhardt, J. and Cumming, T. (2013). The elephant in the single room debate: keeping patients active. *BMJ Clin Res Ed.* 347. | Outcome: No relevant data reported |
| Dean, E. (2010). Patient opinion divided on the introduction of single rooms. *Nurs Stand.* 24(40): 12-13. | Outcome: No relevant data reported |
| Dean, E. (2015). Study finds ideal ward has a mix of single rooms and multi-bed bays. *Nurs Stand.* 29(28): 11. | Outcome: No relevant data reported |
| Dulny, G. *et al.* (2013). An analysis of risk factors of *Clostridium difficile* infection in patients hospitalized in the teaching hospital in 2008. *Przeglad epidemiologiczny* 67(3): 445-50, 547-51. | Intervention: No data on single room |
| Kelly, R. *et al.* (2019). The experience of person-centred practice in a 100% single-room environment in acute care settings-A narrative literature review. *J Clin Nurs.* 28(13-14): 2369-2385. | Outcome: No relevant data reported |
| Lerner, A. O. *et al.* (2019). Environmental contamination by carbapenem-resistant Acinetobacter baumannii: The effects of room type and cleaning methods. *Infect Control Hosp Epidemiol.* 41(2): 166-171. | Outcome: No relevant data reported |
| LI, M. *et al.* (2020). Construction and application of three-dimensional evaluation model of single bed efficiency in hospital. *Chin J Hosp Admin*: 127-130. | Irretrievable |
| Linqvist Leonardsen, AC *et al.* (2016). A qualitative study of patient experiences of decentralized acute healthcare services. *Scand J Prim Health Care* 34(3): 317-324. | Population: not acute hospital |
| Mental Welfare Commission for Scotland (2015) Making progress: older functional assessment wards. https://www.mwcscot.org.uk/sites/default/files/2019-06/making_progress_older_adult_functional_assessment_wards.pdf | Outcome: No relevant data reported |
| NIHR Evidence (2015) NIHR Alert: One size does not fit all – evaluating the move to a hospital with 100% single rooms. https://evidence.nihr.ac.uk/alert/one-size-does-not-fit-all-evaluating-the-move-to-a-hospital-with-100-single-rooms/ | Secondary to Maben 2016 #1802 |
| NIHR Journals Library Health Services and Delivery Research (2015) Evaluating a major innovation in hospital design: workforce implications and impact on patient and staff experiences of all single room hospital accommodation. https://www.journalslibrary.nihr.ac.uk/hsdr/hsdr03030/#/abstract | Duplicate of Maben 2015 #1804 |
| Oliver, D. (2021). David Oliver: Should single rooms be the default for NHS inpatients? *BMJ* 375:n2612. | Outcome: No relevant data reported |
| Pennington, H. and Isles, C. (2013). Should hospitals provide all patients with single rooms? *BMJ* 347: f5695. | Outcome: No relevant data reported |
| Ali, E. (2020) Single-room maternity care: Systematic review and narrative synthesis. *Nurs Open.* 7(6):1661-1670. https://pubmed.ncbi.nlm.nih.gov/33072349 | SLR: citation chase |
| Royal College of Psychiatrists (2020) Next steps for funding mental healthcare in England: Infrastructure. Version 2 https://www.rcpsych.ac.uk/docs/default-source/improving-care/better-mh-policy/policy/next-steps-for-funding-mental-healthcare---infrastructure-royal-college-of-psychiatrists-august-2020.pdf | Outcome: No relevant data reported |
| Russo, PL *et al.* (2018). Establishing the prevalence of healthcare-associated infections in Australian hospitals: Protocol for the Comprehensive Healthcare Associated Infection National Surveillance (CHAINS) study. *BMJ Open* 8(11). | Outcome: No relevant data reported |
| Semret, M. *et al.* (2016). Cleaning the grey zones of hospitals: A prospective, crossover, interventional study. *Am J Infect Control* 44(12): 1582-1588. | Intervention: No data on single room |
| Sengupta, S. *et al.* (2021). Not All Multi.drug Resistant Organism (MDRO)S are Alike-Lessons from Candida Auris in Singapore. *Antimicrobial Resistance and Infection Control* 10(SUPPL 2). | Outcome: No relevant data reported |
| Shannon, MM. *et al.* (2019). Can the physical environment itself influence neurological patient activity? *Disabil Rehabil*. 41(10): 1177-1189. | Intervention: No data on single room |
| Simon, M. *et al.* (2016). Is single room hospital accommodation associated with differences in healthcare-associated infection, falls, pressure ulcers or medication errors? A natural experiment with non-equivalent controls. *J Health Serv Res Policy* 21(3): 147-155. | Secondary publication to Maben 2016, no additional data |
| Teo, R. *et al.* (2015). Patients' preference: Single rooms or shared wards? *Scottish Med J* 59(4): e40. | Outcome: No relevant data reported |
| Welsh Government (2020) National care review of NHS learning disability hospitals provision https://gov.wales/sites/default/files/publications/2020-03/national-care-review-of-learning-disabilities-hospital-inpatient-provision.pdf | Outcome: No relevant data reported |
| Fairhall, K. *et al.* (2009). Single-bed versus multi-bed hospital rooms: The case for patient safety. *World Health Design* 7: 57-61. | Irretrievable |
| BaHammam, A. (2006). Sleep quality of patients with acute myocardial infarction outside the ccu environment: A preliminary study. *Med Sci Monit* 12(4): CR168-CR172. | Outcome: No relevant data reported |
| Harris, S., Farren, M., Janssen, P., Klein, M., & Lee, S. (2004). Single room maternity care offers an efficient and physician friendly environment, without compromising perinatal outcomes. *J Obstetr Gynecol Canada* 26, 633–640. | Irretrievable |
| Ishii, H. *et al.* (2007). Advantages of silent and air-conditioned environment on polysomnography. *Respir Circul* 55(2): 233-236. | Irretrievable |
| Isles, LF., Flynn, R., & Isles, C. (2009). Patient preferences for single rooms or shared accommodation in a district general hospital. *Scottish Med J*. 54, 5–8. | DUPLICATE |
| Jones, R. *et al.* (2016). The effects of single-family rooms on parenting behavior and maternal psychological factors. *J Obstet Gynecol Neonatal Nurs* 45(3): 359-370. | Irretrievable |
| Feeley, N. *et al.* (2020). A comparative study of mothers of infants hospitalized in an open ward neonatal intensive care unit and a combined pod and single-family room design. *BMC Pediatrics* 20(1). | Outcome: No relevant data reported |
| Herr, CE. *et al.* (2003). Additional costs for preventing the spread of methicillin-resistant *Staphylococcus aureus* and a strategy for reducing these costs on a surgical ward. *Infect Control Hosp Epidemiol* 24(9): 673-678. | Intervention: No data on single room |
| Hurst, K. (2009). Do single rooms require more staff than other wards? *Nurs Stand* 24(4): 16. | Population: only data is on staff workload |
| Jobe, AH. (2017). The single-family room neonatal intensive care unit – critical for improving outcomes? *J Pediatr* 185: 10-12. | Outcome: No relevant data reported |
| Joshi, R. *et al.* (2018). Does the architectural layout of a NICU affect alarm pressure? A comparative clinical audit of a single-family room and an open bay area NICU using a retrospective study design. *BMJ Open* 8(6): e022813. | Outcome: No relevant data reported |
| Langley, JM. and Hanakowski, M. (2000). Variation in risk for nosocomial chickenpox after inadvertent exposure. *J Hosp Infect* 44(3): 224-226. | Outcome: No relevant data reported |
| McKinley, LT. *et al.* (2022). Implementation of a nutrition care bundle and improved weight gain of extremely preterm infants to 36 weeks postmenstrual age. *J Pediatr* 241: 42-47.e42. | Outcome: No relevant data reported |
| Mental Welfare Commission for Scotland (2018) Young person monitoring report 2017-2018 https://www.mwcscot.org.uk/sites/default/files/2019-06/young_person_monitoring_report_2017-18.pdf | Outcome: No relevant data reported |
| National Nursing Research Unit (2009) Policy + Issue 17: Splendid Isolation? The pros and cons of single occupancy rooms for the NHS https://www.kcl.ac.uk/nmpc/research/nnru/policy/policy-plus-issues-by-theme/hownursingcareisdelivered/policyissue17.pdf | METHOD |
| Pellikka, HK. *et al.* (2020). Finnish parents' responsibilities for their infant's care when they stayed in a single family room in a neonatal intensive care unit. *J Pediatr Nurs* 53: e28-e34. | Outcome: No relevant data reported |
| Rose, P. and Blythe, S. (2008). Use of single rooms on the children's ward: Part 1. *Paediatr Nurs* 20(10): 13-17. | SLR |
| Rose, P. and Blythe, S. (2009). Use of single rooms on the children's ward, Part 2: Guideline for practice. *Paediatr Nurs* 21(1): 31-35. | Outcome: No relevant data reported |
| Saha, S. *et al.* (2022). Mapping the impact of ICU design on patients, families and the ICU team: A scoping review. *J Crit Care* 67: 3-13. | SLR |
| Servel, AC. and Rideau Batista Novais, A. (2016). [Single-family rooms for neonatal intensive care units impacts on preterm newborns, families, and health-care staff. A systematic literature review]. *Arch Pediatr* 23(9): 921-926. | SLR |
| Shahheidari, M. and Homer, C. (2012). Impact of the design of neonatal intensive care units on neonates, staff, and families: a systematic literature review. *J Perinat Neonat Nurs* 26(3): 260-266. | SLR |
| Shin, JH. *et al*. (2000). Nosocomial cluster of Candida lipolytica fungemia in pediatric patients. Eur J Clin Microbiol Infect Dis 19(5): 344-349. | Intervention: No data on single room |
| Soleimani, F. *et al.* (2020). Impacts of the design of a neonatal intensive care unit (single-family room care and open-ward care) on clinical and environmental outcomes. *Crescent J Med Biol Sci* 7(1): 1-6. | SLR |
| Stolker, JJ. *et al.* (2006). Are patients' views on seclusion associated with lack of privacy in the ward? Arch Psychiatr Nurs 20(6): 282-7. | Outcome: No relevant data reported |
| Tse, Y. (2013). Children thrive on companionship, not single rooms. *BMJ Clin. Res. Ed.* 347: f6335. | METHOD |
| van de Glind, I. *et al.* (2007). Do patients in hospitals benefit from single rooms? A literature review. *Health Policy* 84(2-3): 153-161. | SLR |
| Van Eijk, M. *et al.* (2010). Quality and quantity of sleep in multiple versus single patient room intensive care units. *Intensive Care Med* 36: S189. | Outcome: No relevant data reported |
| van Veenendaal, NR. *et al.* (2019). Hospitalising preterm infants in single family rooms versus open bay units: a systematic review and meta-analysis. *Lancet Child Adolesc Health* 3(3): 147-157. | SLR |
| van Veenendaal, NR. *et al.* (2020). Hospitalising preterm infants in single family rooms versus open bay units: A systematic review and meta-analysis of impact on parents. *EClinicalMedicine* 23. | SLR |
| Vohr, BR. (2019). The importance of parent presence and involvement in the single-family room and open-bay NICU. *Acta Paediatr* 108(6): 986-988. | METHOD |
| WHO Regional Office for Europe – WHO Europe (2009) Capital investment for health. Case studies from Europe (2009) https://www.euro.who.int/__data/assets/pdf_file/0014/43322/E92798.pdf | METHOD |
| Yamaguchi, K. *et al.* (2019). A Study on Operation Architectural Design and Planning of Single-Room PICU in JAPAN Children’s Hospital. Sustainable Urban Environments: Research, Design and Planning for the Next 50 Years, EDRA. | Outcome: No relevant data reported |
| Bigazzi, E., Turrisi, L.,Zagli, G.,Pecile, P.,Bonizzoli, M.,Peris, A. (2010). Bay rooms vs single-bed rooms in intensive care unit nosocomial infections: a case-control study. *CritCare* 14(Suppl.1):P458-P.http://dx.doi.org/10.1186/cc8690[PubMedPMID: PMC2934264]. | Outcome: No relevant data reported |
| Stevens, D., Thompson, P., Helseth, C., Pottala, J., Khan, M., Munson, D. (2011). A comparison of outcomes of care in an open bay and single family room neonatal intensive care facility. *J Neonat Perinat Med* 4: 189–200. | Irretrievable |
| Gotlieb, JB. (2000). Understanding the effects of nurses, patients’ hospital rooms and patients’ perception of control on the perceived quality of a hospital. *Health Mark Q* 18:1–14. | Outcome: No relevant data reported |
| Hamel, M., Zoutman, D., O’Callaghan, C. (2010). Exposure to hospital roommates as a risk factor for health care–associated infection. *Am J Infect Control* 38:173–81. | Intervention: No data on single room |
| Heddema, ER. & van Benthem, BHB. (2011). Decline in incidence of *Clostridium difficile* infection after relocation to a new hospital building with single rooms. *J Hosp Infect* 79, 93–98. | METHOD |
| Okeke, J., Daniel, J., Naseem, A. *et al.* (2013). Impact of all single rooms with ensuite facility in an acute care hospital in Wales (UK). *Age Ageing* 42(Suppl 3):iii1–11. | Outcome: No relevant data reported |
| Okeke, J., Aithal, S., Edwards, C., Ramakrishna, S., & Singh, I. (2014). Outcome of inpatient falls in single bedded and multi-bedded bays. *Age Ageing* 43, ii1–ii11. https://doi.org/10.1093/ageing/afu124. | Outcome: No relevant data reported |
| Shaughnessy, MK., Micielli, RL., DePestel, DD. *et al.* (2011) Evaluation of hospital room assignment and acquisition of Clostridium difficile infection. *Infect Control Hosp Epidemiol* 32(3):201–6. [PubMed: 21460503] | Intervention: No data on single room |
| Shepley, MM., Harris, DD., White, R. (2008). Open-bay and single family room neonatal intensive care units – caregiver satisfaction and stress. *Environ Behav* 40(2):249–268. | Population: Impact on staff only |
| Singh, I., Edwards, C., Okeke, J. (2015). Impact of cognitive impairment on inpatient falls in single room setting and its adverse outcomes. *J Gerontol Geriatr Res* S4. S4eS001. | Secondary publication of Singh 2015, no additional relevant data about single rooms |
| Ulrich, RS. (2006). Effects of single versus multi-bed accommodation and outcomes. Presented at the Symposium on Single-Bed Ward Accommodation, Cardiff, Wales. | METHOD |
| van Oel, CJ., Mlihi, M., & Freeke, A. (2021). Design models for single patient rooms tested for patient preferences. *HERD*, 14(1), 31-46. https://doi.org/10. 1177/1937586720937995 | Outcome: No relevant data reported |
| Williams, C., & Gardiner, C. (2015). Preference for a single or shared room in a UK inpatient hospice: Patient, family and staff perspectives. *BMJ Support Palliat Care* 5, 169–174. doi: 10.1136/bmjspcare-2013-000514 | Population: hospice – not acute hospital |
