## Supplemental Tables for "The clinical, humanistic, and economic outcomes, including experiencing of patient safety events, associated with admitting patients to single rooms compared with shared accommodation for acute hospital admissions. A narrative synthesis systematic literature review"

Health
Economics
Unit

### Appendix

The data presented in summary tables 2–17 have been condensed substantially from what was reported in the papers. For each table there is one row per paper, detailing the setting and population samples in the study, and the outcomes reported according to whether the data were in favour of a single-room design, a shared-room design, or neither in favour nor against either design. Where statistical analyses were conducted the statistical significance is reported in the tables however no other numerical data is presented. Where no formal analysis was reported only the label pertaining to the outcome data are presented. For example, if the proportion of deaths was lower in the single-room design then "% deaths" is reported in the table under the heading "Data favours single-room design", or if qualitative analysis of interviews reports that patients would prefer a shared-room design because is it more sociable then "Qualitative (patient preference, social)" is reported in the table under the heading "Data favours shared-room design".

!

Key and abbreviations

$^a$=adjusted
$^u$=univariate analysis
$^m$=multivariate analysis
NS=not statistically significant ($p<0.05$ is considered statistically significant)
BSI, bloodstream infection; LOS=length of stay; PEMR=physician estimate of mortality risk; SFR=Single family room; SRMC=Single room maternity care

Text is in italics if it is unclear where the true benefit lies, for example where the data is significantly greater for one room type compared to another but the interpretation of benefit may be subjective.

Cells coloured in blue are where a formal comparative statistical analysis was reported.

#### Table 1. Summary of study quality scores

| Citation | Study methodology | QA score |
|---|---|---|
| Adamson 2003[1] | SLR | 82% |
| Anåker et al 2017[2] | Prospective observational, before and after hospital relocation | 59% |
| Anåker et al 2019[3] | Qualitative, before and after hospital relocation | 90% |
| Apple 2014[4] | Prospective observational, qualitative | 52% |
| Bevan et al 2016[5] | Prospective observational | 59% |
| Blandfort et al 2019[6] | Prospective observational, before and after hospital relocation | 67% |
| Blandfort et al 2019[7] | Prospective observational, before and after hospital relocation | 67% |
| Boardman & Forbes 2011[8] | Economic analysis | 91% |
| Bocquet et al 2021[9] | Retrospective observational, case-control | 74% |
| Bodack et al 2016[10] | Prospective observational | 56% |
| Bonizzoli et al 2011[11] | Retrospective observational, before and after hospital relocation | 30% |
| Boztepe et al 2017[12] | Prospective observational | 63% |
| Bracco et al 2007[13] | Prospective observational | 74% |
| Bradbury-Jones et al 2013[14] | SLR | 86% |
| Campbell-Yeo et al 2021[15] | Prospective case–control, before and after hospital relocation | 74% |
| Cantoni et al 2009[16] | Retrospective observational, before and after hospital relocation | 67% |
| Carlson et al 2006[17] | Prospective observational, before and after hospital relocation | 33% |
| Carter et al 2008[18] | Prospective observational, before and after hospital relocation | 33% |
| Caruso et al 2014[19] | Retrospective observational | 74% |
| Cobo et al 2001[20] | Retrospective case–control | 74% |

| | | |
|---|---|---|
| Curtis & Northcott 2017[21] | Qualitative, before and after hospital relocation | 80% |
| Cusack et al 2019[22] | Observational before hospital relocation | 56% |
| Darcy Mahoney et al 2020[23] | Prospective observational | 59% |
| Darley et al 2018[24] | Retrospective observational, before and after hospital relocation | 56% |
| Davis et al 2019[25] | Retrospective observational, before and after hospital relocation | 67% |
| Deitrick et al 2010[26] | Qualitative | 90% |
| de Matos et al 2020[27] | Prospective observational | 63% |
| Domanico et al 2010[28] | Prospective observational, before and after hospital relocation | 63% |
| Domanico et al 2011[29] | Prospective observational, before and after hospital relocation | 63% |
| Douglas & Douglas 2005[30] | Qualitative | 90% |
| Dowdeswell et al 2004[31] | SLR | 36% |
| Dowling et al 2012[32] | Prospective case–control, before and after hospital relocation | 63% |
| Eberhard-Gran et al 2000[33] | Prospective case–control | 59% |
| Edéll-Gustafsson et al 2015[34] | Qualitative | 90% |
| Ehrlander et al 2009[35] | Retrospective observational | 78% |
| Erdeve et al 2008[36] | Prospective case–control | 74% |
| Erdeve et al 2009[37] | Prospective case–control | 78% |
| Erickson et al 2011[38] | Prospective observational before and after hospital relocation | 67% |
| Everts et al 1996[39] | Prospective observational | 52% |
| Felice Tong et al 2018[40] | Retrospective case–control | 78% |
| Ferri et al 2015[41] | Qualitative, before and after hospital relocation | 100% |
| Florey et al 2009[42] | Retrospective case–control, before and after hospital relocation | 44% |
| Foo 2022 et al[43] | Prospective observational | 74% |
| Ford-Jones et al 1990[44] | Prospective observational | 52% |
| Fraenkel et al 2018[45] | Retrospective case-control | 67% |
| Gregersen et al 2021[46] | Retrospective observational, before and after hospital relocation | 70% |
| Grundt et al 2021[47] | Prospective case-control | 67% |
| Halaby et al 2017[48] | Retrospective observational, before and after hospital relocation | 48% |
| Harris et al 2004[49] | Prospective case–control, before and after hospital relocation | 74% |
| Harris et al 2006[50] | Retrospective observational | 63% |
| Harris et al 2006[51] | Retrospective observational | 52% |
| Hosseini & Bagheri 2017[52] | Prospective observational | 63% |
| Hourigan et al 2018[53] | Prospective observational, before and after hospital relocation | 63% |
| Hyun et al 2021[54] | Retrospective case–control | 78% |
| Jansen et al 2021[55] | Retrospective observational, before and after hospital relocation | 63% |

| | | |
|---|---|---|
| Janssen et al 2000[56] | Prospective case–control, before and after hospital relocation | 56% |
| Janssen et al 2006[57] | Prospective observational | 59% |
| Jones et al 2016[58] | Qualitative, before and after hospital relocation | 100% |
| Jongerden et al 2013[59] | Prospective observational, before and after hospital relocation | 67% |
| Jou et al 2015[60] | Retrospective case–control | 74% |
| Julian et al 2015[61] | Retrospective observational | 78% |
| Jung et al 2022[62] | Retrospective observational, before and after hospital relocation | 67% |
| Kainiemi et al 2021[63] | Prospective observational, before and after hospital relocation | 59% |
| Kinnula et al 2008[64] | Prospective observational | 63% |
| Kinnula et al 2012[65] | Prospective observational | 67% |
| Knight & Singh 2016[66] | Prospective observational | 59% |
| Kosuge et al 2013[67] | Prospective observational, before and after hospital relocation | 41% |
| Labarère et al 2004[68] | Prospective observational | 70% |
| Lawson & Phiri 2000[69] | Prospective observational, before and after hospital relocation | 41% |
| Lazar et al 2015[70] | Prospective observational, before and after hospital relocation | 48% |
| Lehtonen et al 2020[71] | Prospective observational | 74% |
| Lester et al 2014[72] | Prospective observational, before and after hospital relocation | 63% |
| Lester et al 2016[73] | Prospective observational, before and after hospital relocation | 59% |
| Liu et al 2019[74] | Qualitative | 100% |
| Lorenz & Dreher 2011[75] | Retrospective case–control | 78% |
| Maben et al 2015[76] | Report, before and after hospital relocation with control hospitals | 78% |
| Maben et al 2016[77] | Prospective observational, before and after hospital relocation with control hospitals | 67% |
| Malcolm 2005[78] | Qualitative | 80% |
| Mattner et al 2007[79] | Prospective observational | 74% |
| McDonald et al 2019[80] | Prospective observational, before and after hospital relocation | 48% |
| McKeown et al 2015[81] | Retrospective observational | 48% |
| Mental Welfare Commission Scotland 1991[82] | Report | 30% |
| Meyer et al 1994[83] | Prospective observational | 59% |
| Milford et al 2008[84] | Prospective observational, before and after hospital relocation | 30% |
| Miller et al 1998[85] | Prospective observational | 59% |
| Monson et al 2018[86] | Prospective case–control | 78% |
| Morgan 2010[87] | Prospective observational | 44% |
| Munier-Marion et al 2016[88] | Prospective observational | 74% |
| Nahas et al 2016[89] | Retrospective observational | 56% |

| | | |
|---|---|---|
| Nash et al 2021[90] | Prospective observational/ qualitative | 63% |
| Nassery & Landgen 2019[91] | Qualitative | 90% |
| OECD & World Health Organization 2019[92] | Report | 14% |
| Olson & Smith1992[93] | Prospective observational | 52% |
| O'Neill et al 2018[94] | Retrospective observational | 74% |
| Park et al 2020[95] | Retrospective observational | 63% |
| Pease & Finlay 2002[96] | Prospective observational | 48% |
| Persson & Määttä 2012[97] | Qualitative | 90% |
| Persson et al 2015[98] | Qualitative | 90% |
| Pilmis et al 2020[99] | Prospective observational | 63% |
| Pineda et al 2012[100] | Prospective case–control | 70% |
| Poncette et al 2021[101] | Retrospective observational | 56% |
| Puumala et al 2020[102] | Retrospective observational, before and after hospital relocation | 67% |
| Pyrke et al 2017[103] | Prospective observational, before and after hospital relocation | 59% |
| Quach et al 2018[104] | Retrospective case–control | 59% |
| Real et al 2018[105] | Prospective observational, before and after hospital relocation | 56% |
| Reed & Shmid 1986[106] | Narrative report, before and after hospital relocation | 10% |
| Reid et al 2015[107] | Prospective observational, before and after hospital relocation | 48% |
| Roos et al 2020[108] | Qualitative, before and after hospital relocation | 90% |
| Rosbergen et al 2020[109] | Prospective observational, before and after hospital relocation | 74% |
| Rowlands & Noble 2008[110] | Qualitative | 90% |
| Sadatsafavi et al 2016[111] | Retrospective economic analysis | 100% |
| Sadatsafavi et al 2019[112] | Retrospective economic analysis, before and after hospital relocation | 100% |
| Sakr et al 2021[113] | Prospective observational | 74% |
| Schalkers et al 2015[114] | Qualitative | 100% |
| Scottish Intercollegiate Guidelines Network 2014[115] | Guideline | 73% |
| Singh et al 2015[116] | Retrospective observational, before and after hospital relocation | 70% |
| Singh et al 2016[117] | Prospective observational | 70% |
| Søndergaard et al 2022[118] | SLR | 91% |
| Song et al 2018[119] | Retrospective observational, before and after hospital relocation | 63% |
| Stelwagen et al 2021[120] | Qualitative | 100% |
| Stevens et al 2011[121] | Prospective observational, before and after hospital relocation | 52% |
| Stevens et al 2012[122] | Prospective observational, before and after hospital relocation | 44% |

| | | |
|---|---|---|
| Stevens et al 2014[123] | Prospective observational, before and after hospital relocation | 56% |
| Stiller et al 2017[124] | Retrospective observational | 59% |
| Swanson et al 2013[125] | Prospective observational, before and after hospital relocation | 37% |
| Tandberg et al 2018[126] | Prospective observational | 70% |
| Tandberg et al 2019[127] | Prospective case–control | 67% |
| Tandberg et al 2019[128] | Prospective observational | 67% |
| Taylor et al 2018[129] | SLR | 91% |
| Tegnestedt et al 2013[130] | Prospective observational | 70% |
| Teltsch et al 2011[131] | Retrospective case–control, before and after hospital relocation | 67% |
| Toivonen et al 2017[132] | Prospective case–control, before and after hospital relocation | 63% |
| Vaisman et al 2018[133] | Retrospective case–control | 67% |
| van de Glind et al 2008[134] | Prospective observational | 74% |
| van der Hoeven et al 2022[135] | Retrospective observational, before and after hospital relocation | 63% |
| Van Enk & Steinberg 2011[136] | Prospective observational, before and after hospital relocation | 44% |
| van Veenendaal et al 2020[137] | Retrospective observational, before and after hospital relocation | 70% |
| Van Veenendaal et al 2022[138] | Prospective observational | 70% |
| Vietri et al 2004[139] | Prospective case–control, before and after hospital relocation | 59% |
| Vohr et al 2017[140] | Prospective observational, before and after hospital relocation | 67% |
| Voigt et al 2018[141] | SLR | 86% |
| Walsh et al 2006[142] | Prospective observational, before and after hospital relocation | 33% |
| Washam et al 2018[143] | Retrospective case–control | 78% |
| Watson et al 2014[144] | Prospective observational, before and after hospital relocation | 44% |
| Zaal et al 2013[145] | Prospective observational | 67% |

Quality is graded by colour: green, good; orange, medium; red, poor. Abbreviation: SLR, systematic literature review.

Health Economics Unit

#### Table 2. Summary of studies reporting mortality data

| Citation | QA | Location | Population | Number of patients/ hospitals | Patient type | Type of admission | Level of care | Data that favour single room | Data showing no difference | Data that favour shared room |
|---|---|---|---|---|---|---|---|---|---|---|
| **Before and after a hospital relocation** | | | | | | | | | | |
| Cantoni 2009[16] | 67% | Switzerland | Adults | 227 patients, 1 hospital | Stem cell transplant | Elective | Routine | | % deaths | |
| Davis 2019[25] | 67% | Australia | Adults | 1569 patients, 1 hospital | Orthopaedic | Elective | Routine | | *p*=0.664 | |
| Domanico 2010,[28] Domanico 2011[29] | 63% | United States | Neonates | 161 carers, 1 hospital, 2 units | Paediatric | NR | NICU | % deaths | | |
| Jansen 2021[55] | 63% | Netherlands | Neonates | 712 patients, 1 hospital, 2 units | Premature neonates | Maternity care | NICU | | p=0.38 all-cause mortality p=0.96 infection-related mortality | |
| Jongerden 2013[59] | 67% | Netherlands | Adults | 323 patients, 1 hospital | Mixed, Adults | Mixed | ICU | | p=0.98 | |
| Jung 2022[62] | 67% | South Korea | Adults | 901 patients, 1 hospital | Mixed | Unclear | ICU | | p=0.168 | |
| Lazar 2015[70] | 48% | Israel | Children | 4162 patients, 1 hospital | Children | Mixed | PICU | | p=0.22 | |
| Puumala 2020[102] | 67% | United States | Neonates | 9995 patients, 1 hospital | Premature neonates | Emergency | NICU | | | % deaths |
| Singh 2015[116] | 70% | United Kingdom | Adults, Elderly | 1749 patients, 1 hospital | Internal medicine, Geriatric | Mixed | Routine | | p=0.12 one-year mortality *p*=0.35 inpatient mortality *p*=0.29 30-day discharge mortality | |
| **Contemporaneous comparison** | | | | | | | | | | |
| Bracco 2007[13] | 74% | Canada | Adults | 2522 patients (of whom 207 known MRS carriers at | Mixed, Post surgery, | Mixed | ICU | *p*<0.001 | | |

| Citation | QA | Location | Population | Number of patients/ hospitals | Patient type | Type of admission | Level of care | Data that favour single room | Data showing no difference | Data that favour shared room |
|---|---|---|---|---|---|---|---|---|---|---|
| | | | | admission), 1 hospital, 1 ward | Medical admission | | | | | |
| Caruso 2014[19] | 74% | Brazil | Adults | 1253 patients, 1 hospital | Adults | Mixed | ICU | | p=0.18 | |
| Harris 2006[50] | 63% | United States | Neonates | 21 parents, 75 HCPs, 11 hospitals | Neonates | Emergency | NICU | | | % deaths |
| Hyun 2021[54] | 78% | South Korea | Adults | 666 patients, 1 hospital | Respiratory, COVID-19 | Emergency | ICU | | | % deaths |
| Julian 2015[61] | 78% | United States | Neonates | 1823 patients 1 hospital, 1 unit | Neonates | Mixed | NICU | | p=0.56 CLOS or mortality | |
| Knight 2016[66] | 59% | United Kingdom | Elderly | 100 patients, 2 hospitals | Geriatric, Dementia | Mixed | Routine | | p>0.95 inpatient mortality p=0.33 30-day mortality | |
| Lehtonen 2020[71] | 74% | 10 countries | Neonates | 4662 patients, 331 units | Neonates | Emergency | ICU | OR 0.76, 0.64-0.89, major morbidity or mortality | OR 0.85, 0.70-1.02, mortality only | |
| Zaal 2013[145] | 67% | Netherlands | Older Adults | 156 patients 1 hospital | Older Adults with dementia | Mixed | ICU | | p=0.72, % deaths | |

#### Table 3. Summary of studies reporting data on patient care and disease management

| Citation | QA | Location | Population | Number of patients/ hospitals | Patient type | Type of admission | Level of care | Data that favour single room | Data showing no difference | Data that favour shared room |
|---|---|---|---|---|---|---|---|---|---|---|
| **Before and after a hospital relocation plus Contemporaneous comparison** | | | | | | | | | | |
| Maben 2016[77] | 67% | United Kingdom | Unclear | 32 patients,<br>21 HCP,<br>1 hospital relocation,<br>2 control hospitals | Mixed | Unclear | Mixed | | Medication errors 9 months after the move | Fewer medication errors immediately after the move |
| **Before and after a hospital relocation** | | | | | | | | | | |
| Davis 2019[25] | 67% | Australia | Adults | 1569 patients,<br>1 hospital relocation | Orthopaedic | Elective | Routine | Lower % medical deterioration requiring rapid response or clinical review | | |
| Lawson 2000[69] | 41% | United Kingdom | Adults | 424 patients, 2 hospitals, 4 wards relocation | Orthopaedic patients | Unclear | Routine | Lower use of painkillers<br>% responders<br>% verbal outbursts<br>% threatening behaviour | | |
| **Contemporaneous comparison** | | | | | | | | | | |
| Ehrlander 2009[35] | 78% | United States | Adults | 117 patients,<br>1 hospital | Mixed | Unclear | Routine | | | Qualitative (feelings of safety) |
| McKeown 2015[81] | 48% | Ireland | Unclear | 880 patients.<br>24 hospitals | End of life | Emergency, Elective | Routine | Perceived acceptability of patient's death<br>Symptom management<br>Symptom experience<br>Patient care | | |
| Nahas 2016[89] | 56% | United Kingdom | Adults, Elderly | 60 patients,<br>2 hospitals | Orthopaedic (elective hip/ knee arthroplasty) | Elective | Routine | p=0.020, cleanliness<br>p=0.015, staff pain management | p=0.190, toileting help given | |

Health Economics Unit

| Citation | QA | Location | Population | Number of patients/ hospitals | Patient type | Type of admission | Level of care | Data that favour single room | Data showing no difference | Data that favour shared room |
|---|---|---|---|---|---|---|---|---|---|---|
| | | | | | | | | p<0.001, pain control | | |
| Van de Glind 2008[134] | 74% | Netherlands | Adults | 52 encounters, 1 hospital | Urology | Unclear | Routine | p=0.003, greater duration of physician-patient encounter<br>% encounter time patient speaks is greater<br>Patients disclose more emotional cues, and information cues<br>p=0.031, more physician responses to the patient cues | % encounter time physician speaks was no different<br>Patients disclose more emotional cues | |
| **Evidence synthesis** | | | | | | | | | | |
| Dowdeswell 2004[31] | SLR 36% | International | Unclear | Unclear | Mixed | Mixed | Mixed | Hospital acquired infection treatment;<br>Hand-hygiene;<br>Cleaning and decontamination;<br>Recovery;<br>In situ medical treatment<br>Family involvement<br>Environment match the patient's progress | | |
| OECD WHO 2019[92] | Report 14% | Europe | NR | NR | Mixed | Mixed | Mixed | | Pain scores | |
| Søndergaard 2022[118] | SLR 91% | International | NR | NR | Acute, Surgical, | Unclear | Routine | Sleep quality<br>Personal control<br>Environment | | |

| Citation | QA | Location | Population | Number of patients/ hospitals | Patient type | Type of admission | Level of care | Data that favour single room | Data showing no difference | Data that favour shared room |
|---|---|---|---|---|---|---|---|---|---|---|
| | | | | | Internal medicine | | | Recovery time | | |
| Taylor 2018[129] | SLR 91% | International | NR | NR | Mixed | Mixed | Mixed | | | Restraint use e.g., rails |
| Voigt 2018[141] | SLR 86% | International | NR | NR | NR | Unclear | Routine | | Medication errors and usage | |

Health Economics Unit

#### Table 4. Summary of studies reporting data on maternity and neonatal care

| Citation | QA | Location | Population | Number of patients/ hospitals | Patient type | Type of admission | Level of care | Data that favour single room | Data showing no difference | Data that favour shared room |
|---|---|---|---|---|---|---|---|---|---|---|
| **Before and after a hospital relocation** | | | | | | | | | | |
| Campbell-Yeo 2021[15] | 74% | Canada | Neonates | 71 mothers, 2 wards | Neonates | Emergency | ICU | Parental presence and involvement (mother and partner feeding) | | |
| Carter 2008[18] | 33% | United States | Adults | 1 hospital 53 parents | Neonates | Emergency | ICU | All p's<0.05 parent perceptions of access to staff | | |
| Domanico 2011[29] | 63% | United States | Neonates | 162 patients (PEMRs 2/3=150, PEMRs 4=12), 1 hospital, 2 units | Paediatric | NR | NICU | PEMR 2-3:<br>patient progress:<br>*p*<0.001, total apnoea events<br>*p*<0.001, apnoea events/day<br>*p*=0.031, days on mother's breastmilk;<br>*p*=0.001, days on mother's breastmilk per LOS;<br>*p*=0.003, interval to enteral feeding;<br>*p*<0.001, interval to breastmilk feeding;<br>*p*=0.048, days on parenteral nutrition;<br>*p*=0.004, days on parenteral nutrition per LOS | PEMR 2-3<br>p=0.94, gestational age<br>p=0.92, admission weight<br>p=NS, acuity<br>p=0.45, weight gain<br>p=0.17, length gain<br>p=0.17, head circumference gain<br>p=0.84, total CPAP days<br>p=0.7, CPAP days/LOS<br>p=0.17, total caffeine days<br>p=0.11, total caffeine days/LOS<br>*p*=0.765, interval to formula feeding<br><br>PEMR 4: | |

| Citation | QA | Location | Population | Number of patients/ hospitals | Patient type | Type of admission | Level of care | Data that favour single room | Data showing no difference | Data that favour shared room |
|---|---|---|---|---|---|---|---|---|---|---|
| | | | | | | | | | p=0.47, gestational age<br>p=0.49, admission weight<br>p=NS, acuity<br>p=0.76, weight gain<br>p=0.47, length gain<br>p=0.70, head circumference gain<br>p=0.59, total CPAP days<br>p=0.94, CPAP days/LOS<br>p=0.82, total caffeine days<br>p=0.94, total caffeine days/LOS<br>p=0.70, total apnoea events<br>p=0.18, apnoea events/day<br>*p*=0.937, interval to enteral feeding<br>*p*=0.571, interval to formula feeding<br>*p*=0.818, days on parenteral nutrition<br>*p*=0.937, days on parenteral nutrition per LOS | |

| Citation | QA | Location | Population | Number of patients/ hospitals | Patient type | Type of admission | Level of care | Data that favour single room | Data showing no difference | Data that favour shared room |
|---|---|---|---|---|---|---|---|---|---|---|
| Dowling 2012[32] | 63% | United States | Neonates | 40 mothers, 1 hospital | Neonates | Emergency | ICU | | p=NS., all breastfeeding measures | |
| Erickson 2011[38] | 67% | United States | Neonates | 73 patients, 1 hospital | Preterm neonates | Emergency | NICU | p=0.04, time to enteral nutrition | p=0.05, weight gain/day<br>p=0.30, weight gain/day normalized to kg birth weight<br>p=0.47, time to parenteral nutrition | |
| Harris 2004[49] | 74% | Canada | Adults | 976 patients, 1 new hospital unit established | Pregnant women | Maternity | Routine | p=0.04, continuous or intermittent electronic foetal monitoring<br>p=0.03, IV therapy<br>p=0.01, 1-minute Apgar <7 | p=NS for augmentation of labour, 20-minutes initial electronic foetal monitoring at admission, epidural, narcotics, mode of delivery, and episiotomy | |
| Hourigan 2018[53] | 63% | United States | Neonates | 32 patients, 1 hospital | Neonates | Emergency | ICU | | *p*=0.30, receiving some maternal or donor breastmilk | *p*=0.04, primarily receiving maternal or donor breastmilk |
| Janssen 2000[56] | 56% | Canada | Adults | 426 patients, 1 hospital relocation | Pregnant women | Maternity | Routine | p<0.001, patient satisfaction with amount of nurse interaction for physical, emotional, and | p=0.10, baby received supplementation with water<br>p=0.25, p=0.05 clear discharge instructions of | |

| Citation | QA | Location | Population | Number of patients/ hospitals | Patient type | Type of admission | Level of care | Data that favour single room | Data showing no difference | Data that favour shared room |
|---|---|---|---|---|---|---|---|---|---|---|
| | | | | | | | | spiritual needs, in labour, and postpartum p<0.001, patient satisfaction with nurse response time, teaching time, information received, feeding related teaching p<0.001, number of babies who received supplementation with formula p<0.001, number breastfeeding p=0.044, number breastfed within 1-2 hours post-delivery p=0.01, clear discharge instructions of when expect a call from the community health nurse p<0.001, clear instructions of how to use car seat, and nurse reviewed handouts | when to call the doctor, and when to make an appointment respectively | |

| Citation | QA | Location | Population | Number of patients/ hospitals | Patient type | Type of admission | Level of care | Data that favour single room | Data showing no difference | Data that favour shared room |
|---|---|---|---|---|---|---|---|---|---|---|
| Lester 2014[72] | 63% | United States | Neonates | 403 patients, 1 hospital relocation | Neonates | Emergency | ICU | Narrative - reduced stress<br><br>p<0.0001, reduced pain | | |
| Puumala 2020[102] | 67% | United States | Neonates | 9995 patients, 1 hospital | Neonates | Emergency | ICU | p<0.001, interval to oral feeding | | |
| Olson 1992[93] | 52% | United States | Adults | 351 patients, 28 HCP, 1 hospital | Pregnant women | Maternity | Routine | *p*<0.05, nurse preferred single rooms<br><br>*p*<0.01, nurse think single room is better for premature neonates | *p*>0.05, nurses think open rooms are better for ventilated/ critically ill infant | |
| Stevens 2012[122] | 44% | United States | Neonates | 73 patients, 1 hospital relocation | Neonates | Emergency | ICU | *p*=0.04, interval to enteric nutrition | p=NS., other nutrition parameters | |
| Swanson 2013[125] | 37% | United States | Neonates, Carers, HCPs | 55 parent surveys,<br>42 AP surveys,<br>151 NN surveys<br>1 hospital relocation | Neonates | Emergency | NICU | p<0.05, Advanced neonatal practitioner perceptions of development, facility and privacy<br>p<0.05, Neonatal nurses perceptions of development, facility and privacy. | Advanced neonatal practitioners: p=NS., teamwork, communication, safety<br><br>Neonatal nurses: p=NS., communication, safety<br><br>Parents: p=NS, development and safety | Neonatal nurses: p<0.05, teamwork |

| Citation | QA | Location | Population | Number of patients/ hospitals | Patient type | Type of admission | Level of care | Data that favour single room | Data showing no difference | Data that favour shared room |
|---|---|---|---|---|---|---|---|---|---|---|
| Toivonen 2017[132] | 63% | Finland | Neonates | 20 nurses, 1 hospital relocation | Neonates | Emergency | ICU | p=0.001, duration of nurse-parent interactions<br>p<0.0001, duration of nurse-family interactions | p=0.349, number of nurse-parent interactions<br>p=0.471, number of nurse-infant interactions<br>p=0.073, duration of nurse-infant interactions<br>p=0.488, number of nurse-family interactions | |
| Van der Hoeven 2022[135] | 63% | Netherlands | Infants | 1293 infants, 1 hospital | Infants | Unclear | ICU | p<0.001, weight at discharge<br>p=0.003, rate of weight gain | p=0.13, gestational age at full enteral feeding | |
| Contemporaneous comparison | | | | | | | | | | |
| Bodack 2016[10] | 55% | Germany | Neonates | 35 sets of parents | Premature neonates | Maternity care | NICU | Qualitative (quality of care) | Qualitative (communication) | |
| Erdeve 2008[36] | 74% | Turkey | Adults, Neonates | 60 infants, 49 mothers, 1 hospital | Preterm neonates | Emergency | NICU | | p=0.084, Routine visits<br>p=0.046, acute care visits<br>p=0.154, number of breastfed infants | p=0.005, more total applications to health services<br><br>p=0.001, more consultations by phone |
| Grundt 2021[47] | 67% | Norway | Neonates | 77 patients, 66 mothers, 2 hospitals, 2 units | Premature neonates | Maternity | NICU | *p*=0.08, *p*=0.06, volume breastmilk produced 7, 14, days post-delivery, respectively<br>p<0.001, *p*=0.02, | *p*=0.71, number of sessions at the breast<br>*p*=0.46, mother breastfeeding self-efficacy<br>*p*=0.51, p=0.33, | |

| Citation | QA | Location | Population | Number of patients/ hospitals | Patient type | Type of admission | Level of care | Data that favour single room | Data showing no difference | Data that favour shared room |
|---|---|---|---|---|---|---|---|---|---|---|
| | | | | | | | | infants breastfed directly and exclusively at discharge, at term, respectively p<0.001, *p*=0.003, p=0.00, infants partly directly breastfed at discharge, at term, and 4 months corrected age, respectively p=0.00[a] use of nipple shields | infants exclusively directly breastfed, or on solids, at 4 months corrected age, respectively *p*=0.33, *p* =0.61, use of nipple shields adjusted for post-menstrual age 33 weeks, 34 weeks, respectively | |
| Lester 2014[72] | 63% | United States | Neonates | 403 patients, 1 hospital | Neonates | Emergency | ICU | p=0.005, weight at discharge p=0.017, rate of weight gain p=0.015, interval to full enteral feeding | | |
| Pineda 2012[100] | 70% | United States | Neonates | 81 patients, 1 hospital | Premature neonates | Emergency | NICU | | p=0.75, breastmilk feeding at discharge | |
| Stelwagen 2021[120] | 100% | Netherlands | Adults | 1 hospital<br>36 parents | Neonates | Emergency | ICU | Narrative - apnoea and periodic breathing | | |
| Tandberg 2019[128] | 67% | Norway | Neonates | 77 patients, 2 hospitals | Neonates | Emergency | ICU | Greater birth weight, length, and head circumference | p=0.45, p=0.42, breastmilk feeding exclusively at discharge, and term +4 months | Greater weight at term +4 months |

| Citation | QA | Location | Population | Number of patients/ hospitals | Patient type | Type of admission | Level of care | Data that favour single room | Data showing no difference | Data that favour shared room |
|---|---|---|---|---|---|---|---|---|---|---|
| | | | | | | | | | | greater length at term +4 months |
| Vohr 2017[140] | 67% | United States | Neonates | 651 patients, 1 hospital relocation | Neonates | Emergency | NICU | *p*<0.001, weight gain per day *p*<0.001, weight gain at discharge *p*=0.002, human milk at 1 week *p*=0.001, human milk at 4 weeks *p*<0.001, volume of milk | | |

#### Table 5. Summary of studies reporting data on complications of disease

| Citation | QA | Location | Population | Number of patients/ hospitals | Patient type | Type of admission | Level of care | Data that favour single room | Data showing no difference | Data that favour shared room |
|---|---|---|---|---|---|---|---|---|---|---|
| **Before and after a hospital relocation plus Contemporaneous comparison** | | | | | | | | | | |
| Maben 2016[77] | 67% | United Kingdom | Unclear | 32 patients, 21 HCP, 1 hospital relocation, 2 control hospitals | Mixed | Unclear | Mixed | | | Pressure ulcers per 1,000 patient-days |
| **Before and after a hospital relocation** | | | | | | | | | | |
| Blandfort 2019[7] | 67% | Denmark | Adults, Elderly | 1014 patients, 2 hospitals | Geriatric, Dementia | Elective | Routine | p=0.02, incidence of delirium | p=0.57, duration of first episode of delirium | |
| Cantoni 2009[16] | 67% | Switzerland | Adults | 227 patients, 1 hospital | Stem cell transplant | Elective | Routine | Number of patients with infections (total, pneumonia, CMV-reactivation, CMV-primary, invasive mould, other)<br>Infection rates (pneumonia: clinical diagnosis) | | Number of patients with infections (microbiologically documented, primary sepsis)<br><br>Infection rates (sepsis, pneumonia, pneumonia: microbiological diagnosis) |
| Davis 2019[25] | 67% | Australia | Adults | 1569 patients, 1 hospital relocation | Orthopaedic | Elective | Routine | | p=0.243, hospital-acquired pressure injuries | |
| Harris 2004[49] | 74% | Canada | Adults | 976 patients, 1 new hospital unit established | Pregnant women | Maternity | Routine | | p=NS for rates of postpartum haemorrhage, pyrexia, rates of thick meconium, and cases of meconium aspiration | |

| Citation | QA | Location | Population | Number of patients/ hospitals | Patient type | Type of admission | Level of care | Data that favour single room | Data showing no difference | Data that favour shared room |
|---|---|---|---|---|---|---|---|---|---|---|
| Lester 2014[72] | 63% | United States | Neonates | 403 patients, 1 hospital relocation | Neonates | Emergency | ICU | Less stress (some related to increased maternal involvement) $p<0.0001$, maternal involvement related to lower pain scores $p<0.0001$, increased maternal involvement in care of the neonate $p<0.0001$, reduction in pain due to the SFR NICU alone | | |
| Singh 2015[116] | 70% | United Kingdom | Adults, Elderly | 1749 patients, 1 hospital relocation | Internal medicine, Geriatric | Mixed | Routine | | | $p<0.01$, hip fractures due to falls |
| Stevens 2012[122] | 44% | United States | Neonates | 73 patients, 1 hospital relocation | Neonates | Emergency | ICU | | OR 1.267, 0.929-1.730, serious adverse outcomes | |
| Lester 2016[73] | 59% | United States | Neonates | 216 patients, 1 hospital relocation | Premature neonates | Maternity | ICU | | $p=0.90$, periventricular leukomalacia $p=0.80$, retinopathy of prematurity (stage 3, 4, 5) $p=0.16$, sepsis $p=0.13$, bronchopulmonary dysplasia | $p=0.09$, necrotising enterocolitis $p=0.08$, intraventricular haemorrhage (grade3/4) |
| Monson 2018[86] | 78% | United States | Neonates | 90 preterm infants, 15 term- | Preterm neonates | Emergency | NICU | | $p=0.35$, bronchopulmonary dysplasia | |

| Citation | QA | Location | Population | Number of patients/ hospitals | Patient type | Type of admission | Level of care | Data that favour single room | Data showing no difference | Data that favour shared room |
|---|---|---|---|---|---|---|---|---|---|---|
| | | | | born control infants, 1 hospital | | | | | p=0.38, infection | |
| **Contemporaneous comparison** | | | | | | | | | | |
| Bracco 2007[13] | 74% | Canada | Adults | 2522 patients (of whom 207 known MRSA carriers at admission), 1 hospital, 1 ward | Mixed, Post surgery, Medical admission | Mixed | ICU | Organ failure | | |
| Caruso 2014[19] | 74% | Brazil | Adults | 1253 patients, 1 hospital | Adults | Mixed | ICU | p<0.01 delirium prevalence p<0.01 medical admissions p<0.01 postoperative admissions | p=0.33 number of days with delirium | |
| Erdeve 2008,[36] Erdeve 2009[37] | 74% | Turkey | Adults, Neonates | 60 infants, 49 mothers, 1 hospital | Preterm neonates | Emergency | NICU | | p=0.720 clinical risk index for babies p=0.673 neonatal therapeutic intensity scoring system | |
| Felice Tong 2018[40] | 78% | Australia | Adults | 185 patients, 1 hospital | Orthopaedic | Elective | Routine | | p=0.70 thromboembolic events within 30-days p=0.21 superficial wound infection within 30-days Deep wound infections p=0.70 | |

| Citation | QA | Location | Population | Number of patients/ hospitals | Patient type | Type of admission | Level of care | Data that favour single room | Data showing no difference | Data that favour shared room |
|---|---|---|---|---|---|---|---|---|---|---|
| | | | | | | | | | medical complications within 30-days | |
| Knight 2016[66] | 59% | United Kingdom | Elderly | 100 patients, 2 hospitals | Geriatric, Dementia | Mixed | Routine | | *p*>0.95, patients with hip fracture as result of inpatient fall | |
| Lehtonen 2020[71] | 74% | Canada, Australia, New Zealand, Finland, Israel, Japan, Spain, Sweden, Switzerland, Italy | Neonates | 4662 patients, 331 units | Preterm neonates | Emergency | ICU | OR 0.76, 0.64-0.89, death or any major morbidity | OR 0.95, 0.84-1.08, composite of mortality or any morbidity<br>OR 0.84, 0.71-1.00, sepsis<br>OR 1.10, 0.95-1.27, Broncho-pulmonary dysplasia<br>OR 1.14, 0.95-1.37, Intraventricular haemorrhage / Periventricular leukomalacia<br>OR 0.81, 0.66-0.99, Retinopathy of prematurity treatment | |
| Vohr 2017[140] | 67% | United States | Neonates | 651 patients, 1 hospital relocation | Neonates | Emergency | NICU | Bayley composites:<br>p=0.02 Cognitive<br>p=0.04 Language<br>p=0.006 Expressive communication<br>p=0.08 Motor<br>p=0.04 Fine motor<br><br>Bayley III composite scores: | Bayley composites:<br>p=0.14 receptive communication<br>p=0.67 gross motor<br>p=0.11 normal neurologic examination | Suspicious neurological examination<br>Abnormal neurological examination |

| Citation | QA | Location | Population | Number of patients/ hospitals | Patient type | Type of admission | Level of care | Data that favour single room | Data showing no difference | Data that favour shared room |
|---|---|---|---|---|---|---|---|---|---|---|
| | | | | | | | | p=0.05, cognitive<br>p=0.02, language<br>p=0.07, motor | | |
| Lester 2014[72] | 63% | United States | Neonates | 403 patients, 1 hospital | Neonates | Emergency | ICU | p=0.05, sepsis | | |
| Zaal 2013[145] | 67% | Netherlands | Older Adults | 156 patients<br>1 hospital | Older Adults with dementia | Mixed | ICU | | p=0.53, crude risk of delirium | |
| **Evidence synthesis** | | | | | | | | | | |
| OECD WHO 2019[92] | Report 14% | Europe | NR | NR | Mixed | Mixed | Mixed | p<0.05<br>Reduced medical errors | | |
| Scottish Intercollegiate Guidelines Network 2019[115] | Report 73% | United Kingdom | Adults | NR | At risk for delirium | NR | Routine | Managing patients with delirium | | |
| Taylor 2018[129] | SLR 91% | International | NR | NR | Mixed | Mixed | Mixed | ICU delirium | | |

### Table 6. Summary of studies reporting data on prevention of infection

| Citation | QA | Location | Population | Number of patients/ hospitals | Patient type | Type of admission | Level of care | Data that favour single room | Data showing no difference | Data that favour shared room |
|---|---|---|---|---|---|---|---|---|---|---|
| **Before and after a hospital relocation plus Contemporaneous comparison** | | | | | | | | | | |
| Maben 2016[77] | 67% | United Kingdom | Unclear | 32 patients, 21 HCP, 1 hospital relocation, 2 control hospitals | Mixed | Unclear | Mixed | *Clostridium difficile* in older people's ward (Control new-build hospital) | | *Clostridium difficile* in older people's ward (Study hospital) |
| **Before and after a hospital relocation** | | | | | | | | | | |
| Bonizzoli 2011[11] | 30% | Italy | Unclear | 818 patients, 1 unit | Trauma | Unclear | ICU | Isolates of MRSA, *Proteus mirabilis*, *Escherichia coli*, *Serratia marcescens*, and *Enterobacter* spp $p<0.01$, amoxicillin/ clavulanate use, ceftriaxone use $p<0.05$ oxacillin use, vancomycin use | | |
| Darley 2018[24] | 56% | United Kingdom | Unclear | 1 hospital relocation | Unclear | Unclear | Routine | $p=0.04$, *Escherichia coli* bacteraemia $p=0.01$, hospital-acquired *Clostridium difficile* infection | $p=0.22$, hospital acquired methicillin-sensitive *Staphylococcus aureus* bacteraemia | |
| Domanico 2011[29] | 63% | United States | Neonates | 162 patients (PEMRs 2/3=150, PEMRs 4=12), 1 hospital, 2 units | Paediatric | NR | NICU | Incidence of nosocomial sepsis (*Candida albicans*, CONS, *Enterococcus* | Incidence of nosocomial sepsis (*Escherichia coli*) | Incidence of nosocomial sepsis (*Enterobacter cloacae*, *Klebsiella pneumoniae*) |

| Citation | QA | Location | Population | Number of patients/ hospitals | Patient type | Type of admission | Level of care | Data that favour single room | Data showing no difference | Data that favour shared room |
|---|---|---|---|---|---|---|---|---|---|---|
| | | | | | | | | *faecalis*, MRSA, *Staphylococcus aureus*, total) | | |
| Davis 2019[25] | 67% | Australia | Adults | 1569 patients, 1 hospital relocation | Orthopaedic | Elective | Routine | | p=0.251, hospital acquired MRSA infections<br>p=0.865, MRSA present on admission | |
| Ferri 2015[41] | 100% | Canada | Adults | 39 HCPs, of which 13 nurses, 7 respiratory therapists, 5 HCPS (other), 6 physicians, 4 family members 4 support staff, 1 hospital | Unclear | Unclear | ICU | Patient perception (6 patients perceived better infection prevention) | | |
| Gregersen 2021[46] | 70% | Denmark | Elderly | 446 patients, 1 hospital relocation | Geriatric | Unclear | Routine | % hospital-acquired infections p=0.01, p=0.03[a] time from admission to first hospital-acquired infection<br>p=0.004 urinary tract infections | p=0.74, pneumonia<br>p=0.50, gastritis<br>p=0.09, sepsis<br>p=0.22, other (wound infection, nephritis, and erysipelas) | |
| Halaby 2017[48] | 48% | Netherlands | Unclear | 16 beds, 1 hospital | Unclear | Unclear | ICU | p=0.001, transmission of any Multidrug resistant bacteria<br>p=0.0015, | p=0.37 transmission of *Morganella* spp | |

| Citation | QA | Location | Population | Number of patients/ hospitals | Patient type | Type of admission | Level of care | Data that favour single room | Data showing no difference | Data that favour shared room |
|---|---|---|---|---|---|---|---|---|---|---|
| | | | | | | | | transmission of *Citrobacter* spp<br>p=0.0005 transmission of *Enterobacter* spp | p=0.99, transmission of *Proteus* spp<br>p=0.25, transmission of *Serratia* spp<br>p=0.39, transmission of *Pseudomonas* spp | |
| Hourigan 2018[53] | 63% | United States | Neonates | 32 patients,<br>1 hospital | Premature neonates | Emergency | NICU | p=0.0001, fewer positive skin swabs<br>p=0.0003, fewer positive environmental swab samples<br>Presence of antibiotic resistance genes (including resistome and virulome) | p=NS<br>comparison of the entire bacterial community at the genus level<br>Potential human pathogenic viruses in 2-week stool, discharge stool and skin samples<br>Species alpha diversity | |
| Jansen 2021[55] | 63% | Netherlands | Neonates | 712 patients<br>1 hospital,<br>2 units | Premature neonates | Maternity care | NICU | | p=0.62, incidence density per 1000 patient-days<br>p=0.59, cumulative incidence per 100 infants<br>p=0.66, skin and/or soft tissue infection<br>p=0.15, conjunctivitis | |
| Jung 2022[62] | 67% | South Korea | Adults | 901 patients,<br>1 hospital | Mixed | Unclear | ICU | p<0.001[a], CRAB acquisition | | |

| Citation | QA | Location | Population | Number of patients/ hospitals | Patient type | Type of admission | Level of care | Data that favour single room | Data showing no difference | Data that favour shared room |
|---|---|---|---|---|---|---|---|---|---|---|
| Lazar 2015[70] | 48% | Israel | Children | 4162 patients, 1 hospital | Children | Mixed | PICU | p=0.01, incidence of BSI<br>p=0.03, nosocomial BSI | p=0.26, community-acquired BSI | |
| McDonald 2019[80] | 48% | Canada | Unclear | 1 hospital relocation | Mixed | Mixed | Mixed | Enterococcus, MRSA, and *Clostridium difficile* infections per 10,000 patient-, days | p=NS, decline in rates of *Clostridium difficile* and MRSA infection | |
| Puumala 2020[102] | 67% | United States | Neonates | 9995 patients, 1 hospital | Premature neonates | Emergency | NICU | p=0.02, sepsis in preterm infants (<28 weeks preterm) | p=0.43, sepsis in preterm infants (28 – 32 weeks preterm)<br>p=0.42, sepsis in preterm infants (32 – 37 weeks preterm) | p=0.001 sepsis in term/post-term infants (>37 weeks) |
| Song 2018[119] | 63% | United States | Neonates | 171 patients, 1 hospital | Premature neonates | Emergency | NICU | hospital-acquired ESBL-E incidence | | |
| Teltsch 2011[131] | 67% | Canada | Adults | 19343 patients, 2 hospitals | Unclear | Unclear | ICU | positive cultures per 10,000 patient-days for yeast, coagulase-negative *Staphylococcus* spp, *Enterococcus* spp, *Staphylococcus aureus*, *Escherichia* spp, *Pseudomonas* spp, *Klebsiella* spp, *Clostridium difficile*, *Corynebacterium* spp, | | Positive cultures per 10,000 patient-days for *Enterobacter* spp, *Haemophilus* spp, MRSA, *Streptococcus viridans*, *Acinetobacter* spp, *Streptococcus pneumoniae*, Group B *Streptococcus* spp, *Neisseria* spp |

| Citation | QA | Location | Population | Number of patients/ hospitals | Patient type | Type of admission | Level of care | Data that favour single room | Data showing no difference | Data that favour shared room |
|---|---|---|---|---|---|---|---|---|---|---|
| | | | | | | | | *Stenotrophomonas maltophilia*, *Citrobacter* spp, *Proteus mirabilis*, *Serratia* spp, fungi, VRE, *Lactobacillus* spp, anaerobic cocci, *Morganella* spp, *Bacteroides* spp, *Moraxella* spp | | |
| Van der Hoeven 2022[135] | 63% | Netherlands | Neonates | 1293 patients, 1 hospital | Premature neonates | Unclear | NICU | Infection of multidrug-resistant organisms<br>Colonisation of third-generation cephalosporin resistant bacteria | Multidrug-resistant organisms: Bacteraemia<br>Colonisation of third-generation cephalosporin resistant bacteria<br>Third-generation cephalosporin resistant bacteria: Bacteraemia | Colonisation of multidrug-resistant organisms |
| Van Veenendaal 2020[137] | 70% | Netherlands | Neonates | 1152 patients, 1 hospital | Neonates | Emergency | NICU | % treated for early-onset sepsis<br>Overall late-onset sepsis<br>OR 0.55, 0.34-0.90<br>OR[a] 0.49, 0.30-0.81<br>Late-onset probable sepsis<br>OR 0.64, 0.38-1.08<br>OR[a] 0.56, 0.32-0.96 | Culture-proven late-onset sepsis<br>OR 0.83, 0.44-1.56<br>OR[a] 0.74, 0.39-1.41<br>Symptoms of late-onset sepsis<br>OR 0.22, 0.05-1.01<br>OR[a] 0.24, 0.05-1.08<br>Late-onset sepsis<br>OR 0.40, 0.16-1.03<br>OR[a] 0.34, 0.13-1.91 | |
| Vietri 2004[139] | 59% | United States | Adults | 261 patients, 1 hospital | Mixed | Unclear | Routine | | Positive MRSA culture | |

| Citation | QA | Location | Population | Number of patients/ hospitals | Patient type | Type of admission | Level of care | Data that favour single room | Data showing no difference | Data that favour shared room |
|---|---|---|---|---|---|---|---|---|---|---|
| Vohr 2017[140] | 67% | United States | Neonates | 651 patients, 1 hospital | Premature neonates | Emergency | NICU | *p*=0.09, sepsis or necrotizing enterocolitis ≥ Bell stage IIA | *p*=0.052, late-onset sepsis | |
| Walsh 2006[142] | 33% | United States | Neonates | 127 nurses, 1 hospital | Neonates | Emergency | NICU | p<0.05, catheter-related BSI | | |
| **Contemporaneous comparison** | | | | | | | | | | |
| Bevan 2016[5] | 59% | United Kingdom | Adults, Elderly | 50 patients, 2 hospitals | Acute medical illness | Emergency | Routine | Patient perception of hygiene and infection risk | | |
| Bocquet 2021[9] | 74% | France | Adults, Children | 233 patients, 1 hospital | Mixed, Influenza | Elective, Emergency | Routine | Nosocomial cases Community-acquired cases | | |
| Bracco 2007[13] | 74% | Canada | Adults | 2522 patients (of whom 207 known MRSA carriers at admission), 1 hospital, 1 ward | Mixed, Post surgery, Medical admission | Mixed | ICU | $p<0.001^{u,m}$, risk of BSI<br>$p<0.05^{u,m}$, risk of MRSA acquisition<br>$p=0.001^{u,m}$, risk of *Pseudomonas* spp acquisition<br>$p<0.001^{u}$<br>$p<0.03^{m}$, risk of *Candida* spp acquisition | | |
| Caruso 2014[19] | 74% | Brazil | Adults | 1253 patients, 1 hospital | Adults | Mixed | ICU | | p=0.19 acquired infections | |
| Cobo 2001[20] | 74% | Spain | Adults | 50 patients, 1 hospital, 2 wards | Respiratory, HIV | Unclear | Routine | | p=0.052, likelihood of multi-drug resistant tuberculosis due to | |

| Citation | QA | Location | Population | Number of patients/ hospitals | Patient type | Type of admission | Level of care | Data that favour single room | Data showing no difference | Data that favour shared room |
|---|---|---|---|---|---|---|---|---|---|---|
| | | | | | | | | | *Mycobacterium bovis* | |
| Everts 1996[39] | 52% | New Zealand | Elderly | 27 patients, 1 hospital | Unclear | Rehabilitation | Routine | Cases of clinical influenza | | |
| Ford-Jones 1990[44] | 52% | Canada | Children | 1530 patients | Cardiological, General admission, Neurosurgical | Unclear | Routine | Cases of nosocomial diarrhoea (GA and neurosurgical unit) | Cases of nosocomial diarrhoea (cardiological unit) | |
| Fraenkel 2018[45] | 67% | Sweden | Adults, Children, Elderly | 251 patients, 8 hospitals | Mixed (all hospitalised patients who acquired norovirus during admission) | Unclear | Routine | p<0.01, norovirus | | |
| Harris 2006[50] | 63% | United States | Neonates | 21 parents, 75 HCPs, 11 hospitals | Neonates | Emergency | NICU | | Nosocomial BSI | Nosocomial pneumonia |
| Julian 2015[61] | 78% | United States | Neonates | 1823 patients 1 hospital, 1 unit | Neonates | Mixed | NICU | p=0.039, MRSA colonization rate for each additional one patient | p=0.10, incidence of MRSA colonization p=0.89, *Clostridium difficile* infection rate | |
| Jou 2015[60] | 74% | United States | Adults | 225 patients, 1 hospital | Mixed | Elective | Mixed | | | p=0.001, nosocomial *Clostridium difficile* infection p<0.001, malignancy |

Health Economics Unit

| Citation | QA | Location | Population | Number of patients/ hospitals | Patient type | Type of admission | Level of care | Data that favour single room | Data showing no difference | Data that favour shared room |
|---|---|---|---|---|---|---|---|---|---|---|
| Kinnula 2008[64] | 63% | Finland | Children | 1927 patients, 1 hospital | Children, infectious disease | Mixed | Routine | p=0.03, risk for hospital acquired infection | | |
| Kinnula 2012[65] | 67% | Finland, Switzerland | Children | 5119 patients, 3 hospitals, 4 wards | Children, mixed | Mixed | Routine | p<0.001, risk for hospital acquired infection during hospitalization (1 hospital) | p=0.56, risk for hospital acquired infection during hospitalization (1 hospital)<br>p=NS, risk of hospital acquired infection after discharge (3 hospitals) | |
| Liu 2019[74] | 100% | Canada | Adults | 1 hospital<br>15 parents of hospitalised infants | Neonates | Emergency | ICU | Parents' perception (reduced spread of infection) | | |
| Lorenz 2011[75] | 78% | United States | Adults, Elderly | 166 patients, 1 hospital | Medical, Surgical, Oncologic | Unclear | Routine | | p=NS, hospital-acquired infections | |
| Mattner 2007[79] | 74% | Germany | Adults | 336 patients, 1 hospital | Cardiovascular, Thoracic surgery | Mixed | ICU | | Enterococci<br>OR 1.06, 0.36-3.12<br>p=0.91 | |
| Monson 2018[86] | 78% | United States | Neonates | 90 preterm infants, 15 term-born control infants, 1 hospital | Preterm neonates | Emergency | NICU | | p=0.38, infection | |
| Morgan 2010[87] | 44% | United Kingdom, United States | Adolescents, Adults, Children | 146 patients, 114 HCP, 2 hospitals | Unclear | Mixed | Routine | HCP preference for isolation and infection control | | |

| Citation | QA | Location | Population | Number of patients/ hospitals | Patient type | Type of admission | Level of care | Data that favour single room | Data showing no difference | Data that favour shared room |
|---|---|---|---|---|---|---|---|---|---|---|
| Munier-Marion 2016[88] | 74% | France | Adults | 93 patients, 1 hospital | Geriatric, Mixed, Surgical | Unclear | Routine | p=0.028, p=0.039[a], risk of hospital-acquired influenza | p=0.16, influenza vaccination coverage | |
| O'Neill 2018[94] | 74% | United States | Mixed | >1 million patients, 218 hospitals with >50% private rooms, 117 with >50% bay rooms | Mixed | Mixed | Mixed | p<0.001, p=0.005[a], central-line-associated BSIs p<0.001, central-line-associated BSIs related mortality | | |
| Park 2020[95] | 63% | United States | Mixed | 2,670,855 discharges, 340 hospitals | Mixed | Mixed | Mixed | p<0.001, p<0.001[a], hospital-acquired MRSA infections | | |
| Pilmis 2020[99] | 63% | France | Adults | 107 patients, 1 hospital | Mixed | Unclear | Routine | p=0.13[u], p=0.0005[m], contamination | | |
| Quach 2018[104] | 59% | Canada, United States | Children | 83,334 patient-days, 2 hospitals | Mixed | Mixed | Mixed | p<0.0001, hospital-acquired respiratory viral infections | | |
| Sadatsafavi 2016[111] | 100% | Canada | Unclear | 8811 patient-days, 1 hospital (simulation) | Medical, Surgical | Unclear | ICU | Annual cases of MRSA acquisition, Pseudomonas species acquisition, and Candida species colonization | | |
| Stiller 2017[124] | 59% | Germany | Unclear | 534 units | Unclear | Unclear | ICU | Polymicrobial BSI OR 0.66, 0.51-0.86 | | |
| Tandberg 2019[128] | 67% | Norway | Neonates | 77 patients, 2 hospitals | Premature neonates | Emergency | NICU | | p=0.36, septicaemia | |

| Citation | QA | Location | Population | Number of patients/ hospitals | Patient type | Type of admission | Level of care | Data that favour single room | Data showing no difference | Data that favour shared room |
|---|---|---|---|---|---|---|---|---|---|---|
| Vaisman 2018[133] | 67% | United States | Adults | 189 patients, 512/515 controls, 1 hospital | Unclear | Unclear | Routine | | P=NS, hospital-onset *Clostridium difficile* | |
| Washam 2018[143] | 78% | United States | Neonates | 1751 patients, 1 hospital | Neonates | Emergency | NICU | p=0.03[u], p=0.03[m], MRSA | | |
| **Evidence synthesis** | | | | | | | | | | |
| OECD WHO 2019[92] | Report 14% | Europe | NR | NR | Mixed | Mixed | Mixed | p<0.05, hospital-acquired infections | | |
| Taylor 2018[129] | SLR 91% | International | NR | NR | Mixed | Mixed | Mixed | 7 studies found advantages only | 3 studies found mixed results<br>4 studies found no difference | |
| Voigt 2018[141] | SLR 86% | International | NR | NR | NR | Unclear | Routine | 10 studies | 5 studies | 16 studies |

#### Table 7 Summary of studies reporting data on patient safety

| Citation | QA | Location | Population | Number of patients/ hospitals | Patient type | Type of admission | Level of care | Data that favour single room | Data showing no difference | Data that favour shared room |
|---|---|---|---|---|---|---|---|---|---|---|
| **Before and after a hospital relocation plus Contemporaneous comparison** | | | | | | | | | | |
| Maben 2016[77] | 67% | United Kingdom | Unclear | 32 patients, 21 HCP, 1 hospital relocation, 2 control hospitals | Mixed | Unclear | Mixed | | | Falls per 1,000 patient-days |
| **Before and after a hospital relocation** | | | | | | | | | | |
| Davis 2019[25] | 67% | Australia | Adults | 1569 patients, 1 hospital relocation | Orthopaedic | Elective | Routine | | p=0.599 Falls in hospital p=0.491 Unwitnessed fall p=0.082 Second fall | |
| Reid 2015[107] | 48% | United Kingdom | Adult, Elderly | 89 patients, 1 hospital relocation | Geriatric | Rehabilitation | Routine | | Falls per 1,000 occupied bed days | |
| Singh 2015[116] | 70% | United Kingdom | Adults, Elderly | 1749 patients, 1 hospital relocation | Internal medicine, Geriatric | Mixed | Routine | | | p<0.01, p<0.01[a], falls per 1,000 patient-bed days p<0.001, falls per in-patient faller |
| **Contemporaneous comparison** | | | | | | | | | | |
| Knight 2016[66] | 59% | United Kingdom | Elderly | 100 patients, 2 hospitals | Geriatric, Dementia | Mixed | Routine | | p=0.83, number of patients who sustained inpatient falls | p=0.035, falls per inpatient faller |
| Lorenz 2011[75] | 78% | United States | Adults, Elderly | 166 patients, 1 hospital | Medical, Surgical, Oncologic | Unclear | Routine | | p=0.37, likelihood of falls | |

Health Economics Unit

| Citation | QA | Location | Population | Number of patients/ hospitals | Patient type | Type of admission | Level of care | Data that favour single room | Data showing no difference | Data that favour shared room |
|---|---|---|---|---|---|---|---|---|---|---|
| Poncette 2021[101] | 55% | Germany | Unclear | 21 beds, 1 hospital | Unclear | Unclear | ICU | | | Alarms raised per bed |
| **Evidence synthesis** | | | | | | | | | | |
| OECD WHO 2019[92] | Report 14% | Europe | NR | NR | Mixed | Mixed | Mixed | $p<0.05$, patient falls | | |
| Taylor 2018[129] | SLR 91% | International | Adults | NR | Mixed | Mixed | Mixed | | No difference | 1 study found disadvantages only |
| Voigt 2018[141] | SLR 86% | International | NR | NR | NR | Unclear | Routine | | 5 studies found no difference | |

#### Table 8. Summary of studies reporting data on readmissions and reinterventions

| Citation | QA | Location | Population | Number of patients/ hospitals | Patient type | Type of admission | Level of care | Data that favour single room | Data showing no difference | Data that favour shared room |
|---|---|---|---|---|---|---|---|---|---|---|
| **Contemporaneous comparison** | | | | | | | | | | |
| Erdeve 2008[36] | 74% | Turkey | Infants | 60 infants,<br>1 hospital | Preterm neonates | Emergency | ICU | p<0.05, hospitalisation | | |
| Felice Tong 2018[40] | 78% | Australia | Adults | 185 patients,<br>1 hospital | Orthopaedic | Elective | Routine | | | p=0.03, return to theatre within 6 weeks |

Health Economics Unit

#### Table 9. Summary of studies reporting views on privacy

| Citation | QA | Location | Population | Number of patients/ hospitals | Patient type | Type of admission | Level of care | Data that favour single room | Data showing no difference | Data that favour shared room |
|---|---|---|---|---|---|---|---|---|---|---|
| **Before and after a hospital relocation plus Contemporaneous comparison** | | | | | | | | | | |
| Maben 2016[77] | 67% | United Kingdom | Unclear | 32 patients, 21 HCP, 1 hospital relocation, 2 control hospitals | Mixed | Unclear | Mixed | Qualitative (privacy, comfort, personal control, visitor flexibility) | | |
| **Before and after a hospital relocation** | | | | | | | | | | |
| Anåker 2019[3] | 90% | Sweden | Adults | 16 patients, 1 hospital | Stroke | Rehabilitation | Routine | Qualitative (privacy, personal control) | | |
| Carlson 2006[17] | 33% | United States | Neonates | 1 hospital, Patients unclear | Neonates | Emergency | ICU | Parent-reported privacy | | |
| Carter 2008[18] | 33% | United States | Adults | 1 hospital 53 parents | Neonates | Emergency | ICU | p<0.001, patients' perception of privacy | | |
| Curtis 2017[21] | 80% | United Kingdom | Children | 1 hospital, 4 wards 17 patients, 60 caregivers, 60 HCPs | Paediatric | Unclear | Routine | Qualitative (privacy) | | |
| Davis 2019[25] | 67% | Australia | Adults | 1569 patients, 1 hospital relocation | Orthopaedic | Elective | Routine | Perception of privacy | | |
| Domanico 2010[28] | 63% | United States | Parents | 1 hospital, 2 units 161 caregivers | Paediatric | NR | NICU | p<0.001, privacy for bonding (long-stay) Transitional parent perceptions: privacy for bonding and for breastfeeding | p=NS, privacy for bonding (short stay) p=0.111 (short stay), p=0.076 (long stay), privacy for breastfeeding | |
| Dowling 2012[32] | 63% | United States | Parents | 1 hospital 40 mothers | Neonates | Emergency | ICU | | p=NS, comfortable pumping breastmilk | |
| Ferri 2015[41] | 100% | Canada | Unclear | 1 hospital, 39 HCPs (13 nurses, | Unclear | Unclear | ICU | Qualitative (privacy) | | |

| Citation | QA | Location | Population | Number of patients/ hospitals | Patient type | Type of admission | Level of care | Data that favour single room | Data showing no difference | Data that favour shared room |
|---|---|---|---|---|---|---|---|---|---|---|
| | | | | 7 respiratory therapists), 5 HCPs (other), 6 physicians, 4 family members 4 support staff | | | | | | |
| Florey 2009[42] | 44% | United Kingdom | Adults | 2 hospitals, 1 before and after move, 80 patients | Medical and surgical, Adults | Unclear | Routine | p<0.001, discussing personal matters p<0.001, patient preference | | |
| Harris 2004[49] | 74% | Canada | Adults | 1 hospital, 976 patients | Pregnant women | Maternity | Routine | p=0.01, physicians' perception of privacy | | |
| Janssen 2000[56] | 56% | Canada | Adults | 1 hospital, 426 patients | Pregnant women | Maternity | Routine | p<0.001, respect shown by caregiver for privacy *p<0.001, greater number of different nurses, doctors, and staff who interacted with the patient* | | |
| Jones 2016[58] | 100% | Australia | Adults, Neonates | 1 hospital relocation 66 mothers, 51 nurses | Adults, Mothers of premature neonates, Nurses | Maternity | NICU | Qualitative (privacy) | | |
| Milford 2008[84] | 30% | United States | Neonates | 1 hospital, patients unclear | Neonates | Emergency | ICU | Staff perceptions of privacy | | |
| Real 2018[105] | 56% | United States | Unclear | 111 patients, 77 nurses, 1 hospital | Cardio-vascular | Unclear | Routine | Privacy | Communication Help from staff | |
| Reid 2015[107] | 48% | United Kingdom | Adult, Elderly | 89 patients, 1 hospital relocation | Geriatric | Rehabilitation | Routine | Qualitative (privacy) | | |

| Citation | QA | Location | Population | Number of patients/ hospitals | Patient type | Type of admission | Level of care | Data that favour single room | Data showing no difference | Data that favour shared room |
|---|---|---|---|---|---|---|---|---|---|---|
| Roos 2020[108] | 90% | Norway | Adults | 39 patients, 1 hospital relocation | Internal medicine, Surgical, Maternity | Maternity, Unclear | Routine | Qualitative (privacy) | | |
| Stevens 2011[121] | 52% | United States | Adults | 1 hospital, 147 patients | Neonates | Emergency | ICU | Patient-reported privacy | | |
| Swanson 2013[125] | 37% | United States | Adults | 1 hospital 55 parents | Neonates | Emergency | ICU | p<0.05, nurses', patients', and advanced practitioners' perceptions of privacy | | |
| **Contemporaneous comparison** | | | | | | | | | | |
| Apple 2014[4] | 52% | Sweden | Adults | 3 ICUs 81 HCP | Mixed | Unclear | ICU | Staff perceptions of privacy | | |
| Bevan 2016[5] | 59% | United Kingdom | Adults, elderly | 2 hospitals 50 patients | Aged 65+ years with acute illness | Emergency | Routine | Qualitative (privacy) | | |
| Bodack 2016[10] | 56% | Germany | Adults | 1 hospital 35 pairs of parents of 40 neonates | Neonates | Emergency | ICU | Patient reported privacy | | |
| Boztepe 2017[12] | 63% | Turkey | Children | 1 hospital, 1 ward 130 | Children | Mixed | Routine | | | Lack of privacy |
| Deitrick 2010[26] | 90% | United States | Adults | 24 patients, 29 HCP, 2 hospitals, 2 wards | Orthopaedic, Neurological, Surgical | Unclear | Routine | Patient preference for privacy | | |
| Douglas 2005[30] | 90% | United Kingdom | Adults | 1 hospital 785 patients (post discharge) | Adults | Unclear | Routine | | *Mixed results* | |
| Ehrlander 2009[35] | 78% | United States | Adults, elderly Adults | 1 hospital 117 patients | Adults | Unclear | Routine | p<0.01, adequate privacy | | |

| Citation | QA | Location | Population | Number of patients/ hospitals | Patient type | Type of admission | Level of care | Data that favour single room | Data showing no difference | Data that favour shared room |
|---|---|---|---|---|---|---|---|---|---|---|
| Harris 2006[50] | 63% | United States | Adults | 5 NICU units<br>SFR=2<br>Open-bay=3<br>HCPs=75<br>Parents=21 | Neonates | Maternity | Level 3, NICU | Parent preference for privacy | | |
| Hosseini 2017[52] | 63% | Iran | Adults | 2 hospitals<br>132 patients | Adults, Medical or surgical | Unclear | Routine | $p<0.001$, adequate privacy | | |
| Janssen 2006[57] | 59% | Canada | Adults | 1 hospital, 2 wards<br>415 patients | Pregnant women | Maternity | Routine | Patient satisfaction with for respect for privacy | | |
| Liu 2019[74] | 100% | Canada | Adults | 1 hospital<br>15 parents of hospitalised infants | Neonates | Emergency | ICU | Privacy enabled the learning and practice of caregiving skills | | |
| Malcolm 2005[78] | 80% | New Zealand | Adults | Hospitals unclear, 12 former patients | Mixed surgery, orthopaedic, medical, obstetric, ENT | Mixed | Routine | Qualitative (privacy) | | Qualitative (supportive) |
| Morgan 2010[87] | 44% | United Kingdom, United States | Children | 2 hospitals<br>146 patients, 114 HCP | Children | Mixed | Routine | Patient perception (privacy)<br>HCP perception (privacy, dignity, confidentiality) | | |
| Nahas 2016[89] | 56% | United Kingdom | Adults, Elderly | 60 patients,<br>2 hospitals | Orthopaedic | Elective | Routine | $p=0.004$, better privacy | | |
| Nash 2021[90] | 63% | Australia | Adults | 4 hospitals<br>602 patients | Indigenous Adults | Theoretical | Routine | Qualitative (privacy) | | |
| Nassery 2019[91] | 90% | Sweden | Adults | 1 hospital,<br>13 interviews (9 individual parents and 4 pairs of parents) | Children | Unclear | Mixed | Qualitative (privacy, comfort) | | |

Health Economics Unit

| Citation | QA | Location | Population | Number of patients/ hospitals | Patient type | Type of admission | Level of care | Data that favour single room | Data showing no difference | Data that favour shared room |
|---|---|---|---|---|---|---|---|---|---|---|
| Olson 1992[93] | 52% | United States | Adults | 1 hospital<br>351 patients,<br>28 HCP | Pregnant women | Maternity | Routine | Patient preference (privacy) | | |
| Persson 2012[97] | 90% | Sweden | Adults, Elderly | 16 patients,<br>10 nurses<br>1 hospital | Orthopaedic, Surgical | Unclear | Routine | Patients in shared rooms signalled their need for privacy | | |
| Persson 2015[98] | 90% | Sweden | Adults | 16 patients,<br>1 hospital | Surgical | Unclear | Routine | Feelings of homeliness | | |
| Rowlands 2008[110] | 90% | United Kingdom | Adults | 1 hospital<br>12 | Adults with advanced cancer | Unclear | Routine | Patient preference (privacy) | | |
| Schalkers 2015[114] | 100% | Netherlands | Children | 8 hospitals<br>63 patients | Children | Mixed | Routine | Qualitative (children's preferences for privacy) | | |
| Stelwagen 2021[120] | 100% | Netherlands | Adults | 1 hospital<br>36 parents | Neonates | Emergency | ICU | | | Privacy violations felt more in single rooms |
| Van de Glind 2008[134] | 74% | Netherlands | Adults | 1 hospital<br>52 encounters | Urology | Unclear | Routine | | Frequency or content of intimate communications | |
| **Evidence synthesis** | | | | | | | | | | |
| Bradbury-Jones 2013[14] | SLR<br>86% | International | Adults | NR | Mixed, Vulnerable, Learning difficulties | Unclear | Unclear | Side rooms ensure privacy | | |
| Dowdeswell 2004[31] | SLR<br>36% | International | Unclear | Unclear | Mixed | Mixed | Mixed | More privacy, which contributes to better outcomes | No quantifiable evidence of improved outcomes | |
| Mental Welfare Commission Scotland 1991[82] | Report<br>30% | United Kingdom | Unclear | 258 patients,<br>28 hospitals | Psychiatric | Unclear | Routine | Easier to meet with visitors | | |

| Citation | QA | Location | Population | Number of patients/ hospitals | Patient type | Type of admission | Level of care | Data that favour single room | Data showing no difference | Data that favour shared room |
|---|---|---|---|---|---|---|---|---|---|---|
| OECD WHO 2019[92] | Report 14% | Europe | NR | NR | Mixed | Mixed | Mixed | p<0.05, improved patient privacy | | |
| Søndergaard 2022[118] | SLR 91% | International | NR | NR | Acute, Surgical, Internal medicine | Unclear | Routine | Privacy, personal control and self-empowerment | | |
| Taylor 2018[129] | SLR 91% | International | NR | NR | Mixed | Mixed | Mixed | | All studies reported advantages and disadvantages | |

#### Table 10. Summary of studies reporting views on patients' loneliness/isolation and family contact

| Citation | QA | Location | Population | Number of patients/ hospitals | Patient type | Type of admission | Level of care | Data that favour single room | Data showing no difference | Data that favour shared room |
|---|---|---|---|---|---|---|---|---|---|---|
| **Before and after a hospital relocation plus Contemporaneous comparison** | | | | | | | | | | |
| Maben 2016[77] | 67% | United Kingdom | Unclear | 32 patients, 21 HCP, 1 hospital relocation, 2 control hospitals | Mixed (all patients in hospital) | Unclear | Mixed | | Mixed findings regarding communication | Not isolated<br><br>More interactions with other patients |
| **Before and after a hospital relocation** | | | | | | | | | | |
| Anåker 2017[2] | 59% | Sweden | Adults | 59 patients, 1 hospital | Stroke | Rehabilitation | Routine | | | Not isolated<br>Availability of interactions with physicians, nurses, nurse assistants, physiotherapists, occupational therapists, speech and language therapist, significant other, other team member, and interpreters |
| Anåker 2019[3] | 90% | Sweden | Adults | 16 patients, 1 hospital | Stroke | Rehabilitation | Routine | | | Less feeling of loneliness and emptiness<br>Have company to talk to |
| Bevan 2016[5] | 59% | United Kingdom | Adults, Elderly | 50 patients, 2 hospitals | Acute illness | Emergency | Routine | Private toilet and showing facilities | | Less feeling of loneliness and isolation<br>Greater companionship and goodwill |

| Citation | QA | Location | Population | Number of patients/ hospitals | Patient type | Type of admission | Level of care | Data that favour single room | Data showing no difference | Data that favour shared room |
|---|---|---|---|---|---|---|---|---|---|---|
| Campbell-Yeo 2021[15] | 74% | Canada | Neonates | 71 mothers, 2 wards | Neonates | Emergency | ICU | More parental presence and involvement, including time with skin-to-skin contact, singing/ talking/ reading to infant, bathing, diaper changes, and providing comfort during painful procedures.<br>More time partner spent holding infants clothed<br>Partner attended rounds at least once during stay | Mothers' attendance at rounds<br><br>Time mothers spent bathing infants | *More time mothers spent holding infants clothed* |
| Curtis 2017[21] | 80% | United Kingdom | Children | 17 patients, 60 caregivers, 60 HCPs, 1 hospital, 4 wards | Paediatric | Unclear | Routine | Enhanced family support | | Socialisation<br>Not isolated |
| Cusack 2019[22] | 56% | Australia | Adults, HCP | 43 nurses,<br>15 patients,<br>1 hospital | Unclear | Unclear | Routine | | | Not isolated |
| Domanico 2010[28] | 63% | United States | Neonates | 161 caregivers, 1 hospital, 2 units | Paediatric | NR | NICU | p=0.012, ability to relax with child (long stay) | p=0.065, perceptions of meeting other parents (short stay)<br>p=0.142 (short stay), p=0.542 (long stay), other | Socialisation<br>p=0.036, perceptions of meeting other parents (long stay) |

| Citation | QA | Location | Population | Number of patients/ hospitals | Patient type | Type of admission | Level of care | Data that favour single room | Data showing no difference | Data that favour shared room |
|---|---|---|---|---|---|---|---|---|---|---|
| | | | | | | | | | parents made stay easier<br>p=0.879, ability to relax with child (short stay) | |
| Ferri 2015[41] | 100% | Canada | Adults | 39 HCPs, of which 13 nurses,<br>7 respiratory therapists,<br>5 HCPS (other),<br>6 physicians,<br>4 family members<br>4 support staff, 1 hospital | Unclear | Unclear | ICU | Increased visitor presence<br>Increased visitor-provider interaction<br>Accommodates Routine and emergency care<br>Patient satisfaction<br>Confidentiality/ privacy | | Socialisation<br>Camaraderie |
| Florey 2009[42] | 44% | United Kingdom | Adults | 80 patients, 2 hospitals, 1 Before and after move | Medical and surgical, Adults | Unclear | Routine | p=0.002, better for visitors | | p<0.001, less loneliness |
| Janssen 2000[56] | 56% | Canada | Adults | 426 patients, 1 hospital | Pregnant women | Maternity | Routine | | *Patient satisfaction regardless of room design:*<br>*p=0.005, time spent with support person*<br>*p=0.007, time spent with baby*<br>*p=0.39, amount of rest* | |
| Jones 2016[58] | 100% | Australia | Neonates | 66 mothers, 51 nurses, 1 hospital relocation | Adults, Mothers of premature neonates, Nurses | Maternity | NICU | Qualitative (personal control, homeliness, accommodates overnight stay, facilitates mother- | | p<0.05, more support<br><br>Qualitative objections to single rooms |

| Citation | QA | Location | Population | Number of patients/ hospitals | Patient type | Type of admission | Level of care | Data that favour single room | Data showing no difference | Data that favour shared room |
|---|---|---|---|---|---|---|---|---|---|---|
| | | | | | | | | infant connection, confidence, parental skills, breastfeeding, and bonding) | | (inconsistent or lack of information, poor interpersonal skills, loneliness, isolation; shared rooms - shared information from other patients, and other patient-nurse interactions) |
| Kainiemi 2021[63] | 59% | Finland | Neonates | 61 families, 1 hospital, 1 unit (pre-post-restructuring) | Pre-term infants (<35 weeks) | Unclear | NICU | p<0.0001, parents', mother's, and father's presence | p=NS, skin-to-skin contact with either parent, mother, or father | |
| Real 2018[105] | 56% | United States | Unclear | 111 patients, 77 nurses, 1 hospital | Cardio-vascular | Unclear | Routine | Qualitative (visitor comfort, better family dynamic) | | |
| Reid 2015[107] | 48% | United Kingdom | Adult, Elderly | 89 patients, 1 hospital relocation | Geriatric | Rehabilitation | Routine | | % feeling lonely | |
| Roos 2020[108] | 90% | Norway | Adults | 39 patients, 1 hospital relocation | Internal medicine, Surgical, Maternity | Maternity, Unclear | Routine | Visiting hours | | Less boredom<br>Not isolated |
| Rosbergen 2020[109] | 74% | Australia | Adults, Elderly | 73 patients, 1 hospital relocation | Stroke, Neurological | Emergency, Rehabilitation | Routine | p=0.02, physical activity | P=NS, social activity<br>Cognitive activity | Less feeling of loneliness |
| Singh 2016[116] | 70% | United Kingdom | Adults, Elderly | 100 patients, 1 hospital relocation | Internal medicine, Geriatric | Mixed | Routine | | | p=0.03[a], less feeling of loneliness |
| Stevens 2011[121] | 52% | United States | Neonates | 147 patients, 1 hospital Before | Neonates | Emergency | ICU | Space for family<br>Accommodations for parents | | |

| Citation | QA | Location | Population | Number of patients/ hospitals | Patient type | Type of admission | Level of care | Data that favour single room | Data showing no difference | Data that favour shared room |
|---|---|---|---|---|---|---|---|---|---|---|
| | | | | and after relocation | | | | | | |
| Stevens 2012[122] | 44% | United States | Neonates | 73 patients, 1 hospital Before and after relocation | Neonates | Emergency | ICU | p=0.017, family-centred care | | |
| Toivonen 2017[132] | 63% | Finland | Neonates | 20 nurses, 1 hospital Before and after relocation | Neonates | Emergency | ICU | p<0.0001, total nurse–family interaction time p=0.001, total nurse–parent interaction time | p=NS, total nurse–infant interaction time | |
| Contemporaneous comparison | | | | | | | | | | |
| Apple 2014[4] | 52% | Sweden | Unclear | 81 HCP, 3 ICUs | Mixed | Unclear | ICU | Qualitative support for single rooms (family involvement, family presence during care) | | |
| Bodack 2016[10] | 56% | Germany | Neonates | 35 pairs of parents of 40 neonates, 1 hospital | Neonates | Emergency | ICU | More secure/ confident caring for baby | | Contact and exchange of knowledge with other parents |
| Darcy Mahoney 2020[23] | 59% | United States, International | Neonates | NR, 277 units | Paediatric, new-born | NR | NICU | p=0.018, parental presence following COVID-19 restrictions p=0.013, parental presence during rounds prior to COVID-19 restrictions | p=NS, parental presence prior to COVID-19 restrictions p=0.6, parental presence during rounds prior to COVID-19 restrictions | |

| Citation | QA | Location | Population | Number of patients/ hospitals | Patient type | Type of admission | Level of care | Data that favour single room | Data showing no difference | Data that favour shared room |
|---|---|---|---|---|---|---|---|---|---|---|
| De Matos 2020[27] | 63% | Brazil | Unclear | 176 family visitors, 1 hospital, 4 ICU units | Cancer | Unclear | ICU | | p=0.52, stress within 24 hrs p=0.15, stress within 7 days | |
| Ehrlander 2009[35] | 78% | United States | Adults | 117 patients, 1 hospital | Adults | Unclear | Routine | Accommodates visitors | p=0.913, loneliness | *>50% enjoy conversation with room mate and gave help to room mate* |
| Erdeve 2008[36] | 74% | Turkey | Infants | 60 infants 1 hospital | Preterm neonates | Emergency | ICU | | | *Time spent with infants during non-hospitalised time* |
| Harris 2006,[50] Harris 2006[51] | 63% | United States | Neonates | 75 HCP, 21 parents, 5 NICU units (SFR=2, open bay=3) | Neonates | Unclear | Level 3, NICU | | Contact with other parents | |
| Hosseini 2017[52] | 63% | Iran | Adults | 132 patients, 2 hospitals | Medical, Surgical | Unclear | Routine | p<0.001, visitor convenience | | p<0.001, less feeling of loneliness |
| Liu 2019[74] | 100% | Canada | Neonates | 15 parents, 1 hospital | Neonates | Emergency | ICU | Qualitative (engage in parenting activities beyond basic caregiving) | Qualitative (isolation) | |
| Malcolm 2005[78] | 80% | New Zealand | Adolescents, Adults | 12 former patients | Mixed surgery, orthopaedic, medical, obstetric, ENT | Mixed | Routine | | | Qualitative (camaraderie and support) |
| Milford 2008[84] | 30% | United States | Neonates | No. of patients unclear, 1 hospital | Neonates | Emergency | ICU | Staff perception of discussions with families | | |

| Citation | QA | Location | Population | Number of patients/ hospitals | Patient type | Type of admission | Level of care | Data that favour single room | Data showing no difference | Data that favour shared room |
|---|---|---|---|---|---|---|---|---|---|---|
| Morgan 2010[87] | 44% | UK, US | Children | 146 patients, 114 HCP, 2 hospitals | Children | Mixed | Routine | Patients' privacy<br>Visitor times<br>Undisturbed sleep<br>Personal control | | Patients:<br>Communication<br>Company<br>Entertainment<br><br>HCPs:<br>Interaction with other patients<br>Company |
| Nahas 2016[89] | 56% | United Kingdom | Adults, Elderly | 60 patients, 2 hospitals | Orthopaedic | Elective | Routine | | p=0.754, isolation<br>p=0.638, loneliness | |
| Nash 2021[90] | 63% | United Kingdom | Adults, Elderly | 100 patients, | Adults >65 years, recovering from acute illness | Emergency | Routine | Company of family, not strangers | | Qualitative (not isolated, social interactions) |
| Nassery 2019[91] | 90% | Sweden | Children | 13 parents, 1 hospital | Children | Unclear | Mixed | Qualitative (patient preference, privacy, stress, quieter) | | Qualitative (shared experience and advice) |
| Olson 1992[93] | 52% | United States | Adults | 351 patients, 28 HCP, 1 hospital | Pregnant women | Maternity | Routine | | *Mothers satisfied with visiting hours* | |
| Pease 2002[96] | 48% | United Kingdom | Unclear | 50 patients, 1 hospital | Oncologic, Terminal | Unclear | Routine | | | Qualitative (not isolated) |
| Persson 2012[97] | 90% | Sweden | Adults, Elderly | 16 patients, 10 nurses, 1 hospital | Orthopaedic, Surgical | Unclear | Routine | | | Qualitative (not isolated) |
| Persson 2015[98] | 90% | Sweden | Adults | 16 patients, 1 hospital | Surgical | Unclear | Routine | | | Qualitative (not isolated, company, social contact) |
| Pineda 2012[100] | 70% | United States | Neonates | 81 patients, 1 hospital | Premature neonates | Emergency | NICU | p=0.021[a], time parents spent | p=NS, time parents spent holding the infant | |

| Citation | QA | Location | Population | Number of patients/ hospitals | Patient type | Type of admission | Level of care | Data that favour single room | Data showing no difference | Data that favour shared room |
|---|---|---|---|---|---|---|---|---|---|---|
| | | | | | | | | visiting the infant (week 1-2) p=0.039, time spent cuddling, visiting, and with skin-to-skin contact (week 1-2) p=0.026, p=0.017[a], time parents spent visiting the infant during weeks 3-4 p=0.062, p=0.047[a], time parents spent visiting the infant by LOS | p=0.193, time parents spent visiting the infant (week 5–term) p=0.593, interval to first time parents hold infant p=0.810, days spent cuddling infant (week 1–2) p=0.548, days spent cuddling infant (week 3–4) p=0.592, days spent cuddling infant (week 5–term) p=0.361, days spent cuddling infant by LOS p=0.496, days with skin-to-skin (week 1–2) p=0.111, days with skin-to-skin contact (week 3–4) p=0.489, days with skin-to-skin contact (week 5–term) p=0.360, days with skin-to-skin contact by LOS | |
| Rowlands 2008[110] | 90% | United Kingdom | Adults | 12 patients, 1 hospital | Adults with advanced cancer | Unclear | Routine | Qualitative (privacy) | | Qualitative (social interactions) |

| Citation | QA | Location | Population | Number of patients/ hospitals | Patient type | Type of admission | Level of care | Data that favour single room | Data showing no difference | Data that favour shared room |
|---|---|---|---|---|---|---|---|---|---|---|
| Schalkers 2015[114] | 100% | Netherlands | Children | 8 hospitals<br>63 patients | Children | Mixed | Routine | | | Qualitative (company, patient preference if they have similarities with other patients) |
| Stelwagen 2021[120] | 100% | Netherlands | Neonates | 36 parents, 1 hospital Before and after relocation | Neonates | Emergency | ICU | Qualitative (family communication and closeness, personal control, privacy, tranquillity, comfort, practicing parenting skills) | | Qualitative (not isolated, ability to distance themselves from invasive procedures) |
| Swanson 2013[125] | 37% | United States | Neonates, Carers | 55 parents,<br>1 hospital | Neonates | Emergency | NICU | $p<0.05$ advanced practitioners' satisfaction with communication | | $p<0.05$, nurse satisfaction with communication<br>$p<0.05$, nurse satisfaction with team |
| Tandberg 2018[126] | 70% | Norway | Neonates | 64 patients, 115 parents, 2 hospitals | Neonates, Premature neonates | Emergency | ICU | $p<0.001$, time mother and father present during first 14 days<br>$p=0.02$, mother's skin-to-skin contact per 24 h<br>$p=0.05$, father's skin-to-skin contact per 24 h<br>$p=0.02$, guidance provided by the staff meets needs of mothers | $p=0.53$, guidance provided by the staff meets needs of fathers<br>$p=0.21$, fathers felt their opinions were considered | |

| Citation | QA | Location | Population | Number of patients/ hospitals | Patient type | Type of admission | Level of care | Data that favour single room | Data showing no difference | Data that favour shared room |
|---|---|---|---|---|---|---|---|---|---|---|
| | | | | | | | | p=0.04, mothers felt their opinions were considered p<0.001 (mothers), p=0.01 (fathers), participation in doctor visits, respectively p=0.05 (mothers), p<0.001 (fathers), emotion support received from staff | | |
| Tandberg 2019[127] | 67% | Norway | Infants | 77 infants, 132 parents, 2 hospitals | Infants | Emergency | ICU | p<0.0001, mother's and father's presence in week 1 p<0.0001, mother's and father's presence per day up to week 34 | | |
| Tandberg 2019[128] | 67% | Norway | Neonates | 77 patients, 2 hospitals | Neonates | Emergency | ICU | p<0.001, mother's presence in week 1 p value<0.001, father's presence in week 1 p<0.001, mother's presence overall and continuous p<0.001, father's presence overall and continuous p<0.001, skin-to-skin contact per day in week 1 | | |

| Citation | QA | Location | Population | Number of patients/ hospitals | Patient type | Type of admission | Level of care | Data that favour single room | Data showing no difference | Data that favour shared room |
|---|---|---|---|---|---|---|---|---|---|---|
| | | | | | | | | p<0.001, total skin-to-skin contact per day | | |
| Van Veenendaal 2022[138] | 70% | Netherlands | Neonates | 182 parents, 3 hospitals | Fathers of neonates | Emergency | ICU | p<0.001, p<0.001[a], total presence p<0.001, p<0.001[a], presence >8 h p<0.001, p=0.009[a], total participation p<0.001, p=0.005[a], participation in medical care p=0.23, p=0.04[a], information gathering p<0.001, p=0.005[a], advocacy and leadership p=0.006, p=0.005[a], time spent with neonate | p=0.04, p=0.13[a], participation in daily care p=0.69, p=0.57[a], time spent comforting neonate | |
| **Evidence synthesis** | | | | | | | | | | |
| Adamson 2003[1] | SLR 82% | United States, International | Mixed | Unclear | Mixed | Mixed | Mixed | Interaction with family members and flexibility for accommodating family members | | |
| Dowdeswell 2004[31] | SLR 36% | International | Unclear | Unclear | Mixed | Mixed | Mixed | Qualitative (frequency of visitors, privacy) | | |
| OECD WHO 2019 [92] | Report 14% | Europe | NR | NR | Mixed | Mixed | Mixed | p<0.05, social support Communication with family | | |

| Citation | QA | Location | Population | Number of patients/ hospitals | Patient type | Type of admission | Level of care | Data that favour single room | Data showing no difference | Data that favour shared room |
|---|---|---|---|---|---|---|---|---|---|---|
| Søndergaard 2022[118] | SLR 91% | International | NR | NR | Acute, Surgical, Internal medicine | Unclear | Routine | Quiet, private, better /easier communication | | Not isolated and not lonely |
| Taylor 2018[129] | SLR 91% | International | NR | NR | Mixed | Mixed | Mixed | | All studies reported advantages and disadvantages | |

#### Table 11. Summary of studies reporting patient's views on noise, disturbance and sleep

| Citation | QA | Location | Population | Number of patients/ hospitals | Patient type | Type of admission | Level of care | Data that favour single room | Data showing no difference | Data that favour shared room |
|---|---|---|---|---|---|---|---|---|---|---|
| **Before and after a hospital relocation plus Contemporaneous comparison** | | | | | | | | | | |
| Maben 2016[77] | 67% | United Kingdom | Unclear | 32 patients,<br>21 HCP,<br>1 hospital relocation, 2 control hospitals | Mixed | Unclear | Mixed | Patient perceptions (comfort, noise levels, privacy) | | |
| **Before and after a hospital relocation** | | | | | | | | | | |
| Carlson 2006[17] | 33% | United States | Neonates | Unclear, 1 hospital | Neonates | Emergency | ICU | Patient perceptions (noise levels) | | |
| Carter 2008[18] | 33% | United States | Neonates | 53 parents, 1 hospital | Neonates | Emergency | NICU | p<0.001, noise level<br>p<0.001, lighting | | |
| Davis 2019[25] | 67% | Australia | Adults | 1569 patients,<br>1 hospital relocation | Orthopaedic | Elective | Routine | *Adequate sleep reported but no comparison with shared room* | | |
| Domanico 2010[28] | 63% | United States | Neonates | 161 caregivers, 1 hospital, 2 units | Paediatric | NR | NICU | Actual noise levels | Patient perceptions (noise levels)<br>p=0.890, noise disturbance (short stay)<br>p=0.657, noise disturbance (long stay) | |
| Ferri 2015[41] | 100% | Canada | Adults | 39 HCPs (13 nurses, 7 respiratory therapists),<br>5 HCPS (other), | Unclear | Unclear | ICU | Qualitative (less disruption) | | |

| Citation | QA | Location | Population | Number of patients/ hospitals | Patient type | Type of admission | Level of care | Data that favour single room | Data showing no difference | Data that favour shared room |
|---|---|---|---|---|---|---|---|---|---|---|
| | | | | 6 physicians,<br>4 family members<br>4 support staff,<br>1 hospital | | | | | | |
| Florey 2009[42] | 44% | United Kingdom | Adults | 80 patients, 2 hospitals, 1 Before and after move | Medical and surgical, Adults | Unclear | Routine | p=0.019, noise disturbance | | |
| Harris 2004[49] | 74% | Canada | Adults | 976 patients, 1 hospital, Before and after new unit established | Pregnancy | Maternity | Routine | p<0.001, physicians' perceptions of noise | | |
| Janssen 2000[56] | 56% | Canada | Adults | 426 patients, 1 hospital, Before and after relocation | Pregnant women | Maternity | Routine | p<0.001, any noise disturbance<br>p<0.001, talking/ visiting by hospital neighbours<br>p=0.08, staff talking at the nursing station<br>p<0.001, crying babies | p=0.30, talking/visiting by hospital staff<br>p=0.28, women in labour | |
| Maben 2015[76] | 78% | United Kingdom | Unclear | 24 staff, 32 patients, 1 hospital (relocated), 2 control hospitals | All patients in hospital | Mixed | Mixed | Patient perceived benefit | | |
| Milford 2008[84] | 30% | United States | Neonates | Unclear, 1 hospital | Neonates | Emergency | ICU | Higher staff satisfaction | | |
| Pyrke 2017[103] | 59% | Canada | Adults | 47 patients,<br>1 hospital relocation | Psychiatric | Emergency | Routine | | p=0.399, sleep disturbed<br>p=0.065, time spent asleep | |

Health Economics Unit

| Citation | QA | Location | Population | Number of patients/ hospitals | Patient type | Type of admission | Level of care | Data that favour single room | Data showing no difference | Data that favour shared room |
|---|---|---|---|---|---|---|---|---|---|---|
| Real 2018[105] | 56% | United States | Unclear | 111 patients,<br>77 nurses,<br>1 hospital,<br>1 ward | Cardio-vascular | Unclear | ICU, Routine | Perceived noise level | | |
| Stevens 2011[121] | 52% | United States | Neonates | 147 patients, 1 hospital Before and after relocation | Neonates | Emergency | ICU | Restfulness | | |
| Stevens 2012[122] | 44% | United States | Neonates | 73 patients, 1 hospital Before and after relocation | Neonates | Emergency | ICU | $p<0.001$, actual noise level<br>$p<0.05$, lighting | Noise level adjacent to baby's ear | |
| Van Enk 2011[136] | 44% | United States | Neonates | 90 beds, 1 hospital | Neonates | Emergency | NICU | $p=0.04$, actual noise level (day time)<br>*$p=0.05$, less illumination (day time)*<br>*$p=0.01$, lower temperature (night time)*<br>*$p=0.001$, lower temperature (day and night combined)*<br>*$p<0.0001$, lower humidity (night time)* | $p=0.35$, actual noise level (night time)<br>$p=0.08$, actual noise level (day or night time)<br>$p=0.49$, illumination (night time)<br>$p=0.60$, temperature (day time) | *$p<0.0001$, lower humidity (day time)*<br>*$p<0.0001$, lower humidity (day and night combined)* |
| Walsh 2006[142] | 33% | Unclear | Neonates | 127 nurses, 1 hospital | Neonates | Emergency | NICU | Actual noise levels | | |
| **Contemporaneous comparison** | | | | | | | | | | |

| Citation | QA | Location | Population | Number of patients/ hospitals | Patient type | Type of admission | Level of care | Data that favour single room | Data showing no difference | Data that favour shared room |
|---|---|---|---|---|---|---|---|---|---|---|
| Apple 2014[4] | 52% | Sweden | Unclear | 81 HCP, 3 ICUs | Mixed | Unclear | ICU | Qualitative (privacy, fewer disturbances) | | |
| Bevan 2016[5] | 59% | United Kingdom | Adults, elderly | 50 patients, 2 hospitals | Acute illness | Emergency | Routine | Qualitative (less noise disturbance) | | |
| Bodack 2016[10] | 56% | Germany | Neonates | 35 pairs of parents of 40 neonates, 1 hospital | Neonates | Emergency | ICU | Qualitative (fewer disturbances) | | |
| Deitrick 2010[26] | 90% | United States | Adults | 24 patients, 29 HCP, 2 hospitals, 2 wards | Orthopaedic, Neurological, Surgical | Unclear | Routine | | | Qualitative (adequate rest and sleep due to the presence of a roommate) |
| Douglas 2005[30] | 90% | United Kingdom | Unclear | 785 patients (post discharge), 1 hospital | Surgical, Acute care, Maternity, Geriatric | Unclear | Routine | Fewer night-time disturbances | | |
| Eberhard-Gran 2000[33] | 59% | Norway | Adults | 160 patients, Unclear (one municipality) | Adults, Pregnant women | Maternity | Routine | More sleep/ rest Enough sleep/ rest (women ≥ 30 years old) OR 8.1, 1.7-39.3 amount of sleep and rest at Akershus | Enough sleep/rest OR 2.9, 0.3-30.3 amount of sleep and rest at Kongsvinger | |
| Edéll-Gustafsson 2015[34] | 90% | Sweden | Neonates | 12 parents, 1 unit | Neonates | Emergency | ICU | Qualitative (privacy, personal control) | | Qualitative (not confined) |
| Ehrlander 2009[35] | 78% | United States | Adults | 117 patients, 1 hospital | Adults | Unclear | Routine | Qualitative (peace and quiet) | | |

| Citation | QA | Location | Population | Number of patients/ hospitals | Patient type | Type of admission | Level of care | Data that favour single room | Data showing no difference | Data that favour shared room |
|---|---|---|---|---|---|---|---|---|---|---|
| Foo 2022[43] | 74% | Australia | Adults | 60 patients, 1 hospital | Cardio-respiratory, Obstetric, Sleep disorders, Other | Unclear | Routine | | $p>0.05$, number of interruptions in 24-h<br>$p>0.05$, number of disturbances at night<br>$p=0.11$, other measures of discomfort | |
| Harris 2006[51] | 52% | United States | Neonates | 21 parents, 75 HCPs | Neonates | Maternity | ICU | Parent satisfaction with physical environment | | |
| Hosseini 2017[52] | 63% | Iran | Adults | 132 patients, 2 hospitals | Medical, surgical | Unclear | Routine | | | $p<0.001$, better scores for sleep disorders |
| Meyer 1994[83] | 59% | United States | Unclear | Unclear, 1 hospital | Mixed | Mixed | Mixed | $p<0.05$, actual noise levels (day time)<br>$p<0.05$, actual noise levels (night time)<br>lower maximum illumination (day and night time) | Maximum period of uninterrupted sleep | |
| Morgan 2010[87] | 44% | UK, US | Children | 146 patients, 114 HCP, 2 hospitals | Children | Mixed | Routine | Qualitative (quiet sleep) | | |
| Nahas 2016[89] | 56% | United Kingdom | Adults, Elderly | 60 patients, 2 hospitals | Orthopaedic (elective hip/ knee arthroplasty) | Elective | Routine | $p=0.003$, good sleep at night | $p=0.127$, noise level | |
| Nassery 2019[91] | 90% | Sweden | Children | 13 interviews (9 individual parents, 4 pairs of | Children | Unclear | Mixed | Less stress sleeping alone | | |

| Citation | QA | Location | Population | Number of patients/ hospitals | Patient type | Type of admission | Level of care | Data that favour single room | Data showing no difference | Data that favour shared room |
|---|---|---|---|---|---|---|---|---|---|---|
| | | | | parents), 1 hospital | | | | | | |
| Olson 1992[93] | 52% | United States | Adults | 351 patients, 28 HCP, 1 hospital | Pregnant women | Maternity | Routine | *Mothers satisfied with room but no comparison with shared rooms* | | |
| Persson 2012[97] | 90% | Sweden | Adults, Elderly | 16 patients, 10 nurses 1 hospital, 2 wards | Orthopaedic, Surgical | Unclear | Routine | Less disturbance | | |
| Persson 2015[98] | 90% | Sweden | Adults | 16 patients, 1 hospital | Surgical | Unclear | Routine | Sleep undisturbed | | |
| Poncette 2021[101] | 56% | Germany | Unclear | 21 beds, 1 hospital | Unclear | Unclear | ICU | | | Less alarms raised |
| Rowlands 2008[110] | 90% | United Kingdom | Adults | 12 patients, 1 hospital | Adults with advanced cancer | Unclear | Routine | Qualitative (less stress related to disturbing others) | | |
| Sakr 2021[113] | 74% | Lebanon | Adults | 75 patients, 1 hospital | Internal medicine, Surgical | Mixed | Routine | p=0.011, fewer cases of new onset insomnia | p=0.272, patient perceived impact of room on new onset insomnia | |
| Stelwagen 2021[120] | 100% | Netherlands | Neonates | 36 parents, 1 hospital | Neonates | Emergency | ICU | Qualitative (privacy) | | Qualitative (less surprise when staff appear at bedside) |
| Tegnestedt 2013[130] | 70% | Sweden | Adults, Elderly | 15 patients 1 hospital | Adults | Emergency | ICU | | p=0.777 (7am to 3pm), p=0.885(3pm to 11pm), p=0.832 (11pm to 7am), actual noise | |

| Citation | QA | Location | Population | Number of patients/ hospitals | Patient type | Type of admission | Level of care | Data that favour single room | Data showing no difference | Data that favour shared room |
|---|---|---|---|---|---|---|---|---|---|---|
| Zaal 2013[145] | 67% | Netherlands | Older Adults | 156 patients<br>1 hospital | Older Adults with dementia | Mixed | ICU | | | p <0.001 lower light intensity |
| **Evidence synthesis** | | | | | | | | | | |
| Dowdeswell 2004[31] | SLR 36% | International | Unclear | Unclear | Mixed | Mixed | Mixed | Quieter (less sleep disturbance, better outcomes) | | |
| OECD WHO 2019[92] | Report 14% | Europe | NR | NR | Mixed | Mixed | Mixed | p<0.05, better sleep | | |
| Søndergaard 2022[118] | SLR 91% | International | NR | NR | Acute, Surgical, Internal medicine | Unclear | Routine | Quieter (less sleep disturbance) | | |
| Taylor 2018[129] | SLR 91% | International | NR | NR | Mixed | Mixed | Mixed | | Mixed findings on sleep outcomes | |

### Table 12. Summary of studies reporting patients' views on satisfaction with care

| Citation | QA | Location | Population | Number of patients/ hospitals | Patient type | Type of admission | Level of care | Data that favour single room | Data showing no difference | Data that favour shared room |
|---|---|---|---|---|---|---|---|---|---|---|
| **Before and after a hospital relocation plus Contemporaneous comparison** | | | | | | | | | | |
| Maben 2016[77] | 67% | United Kingdom | Unclear | 32 patients,<br>21 HCP,<br>1 hospital relocation, 2 control hospitals | Mixed | Unclear | Mixed | Patient preference (privacy, ensuites) | | Patient preference (social interaction) |
| **Before and after a hospital relocation** | | | | | | | | | | |
| Campbell-Yeo 2021[15] | 74% | Canada | Neonates | 71 mothers, 2 wards | Neonates | Emergency | ICU | Postpartum depression scores<br>Post-traumatic stress disorder scores | Parental stressor scores<br>EQ-5D-5L self-reported health | Perceived maternal self-efficacy<br>Intolerance of uncertainty |
| Carlson 2006[17] | 33% | United States | Neonates | 1 hospital,<br>Patients unclear | Neonates | Emergency | ICU | Patient perception (improved lighting control) | | |
| Carter 2008[18] | 33% | United States | Neonates | 53 parents,<br>1 hospital Before and after relocation | Neonates | Emergency | NICU | p<0.001<br>parent perceptions of security | | |
| Davis 2019[25] | 67% | Australia | Adults | 1569 patients,<br>1 hospital relocation | Orthopaedic | Elective | Routine | Patient satisfaction but no comparison with shared room | | |
| Florey 2009[42] | 44% | United Kingdom | Adults | 80 patients, 2 hospitals, 1 Before and after move | Medical and surgical, Adults | Unclear | Routine | | patient preference based on previous experience inconclusive | |
| Janssen 2000[56] | 56% | Canada | Adults | 426 patients, 1 hospital | Pregnant women | Maternity | Routine | p<0.001, patient opinions in care considered<br>p<0.001, information given to inform choices | | |

| Citation | QA | Location | Population | Number of patients/ hospitals | Patient type | Type of admission | Level of care | Data that favour single room | Data showing no difference | Data that favour shared room |
|---|---|---|---|---|---|---|---|---|---|---|
| | | | | | | | | p<0.001, patient choices supporter by caregivers<br>p<0.001, assistance given to support person<br>p<0.001, comfort measures for labour pain<br><br>p<0.001, comfort measures for pain after birth | | |
| Jongerden 2013[59] | 67% | Netherlands | Adults | 387 patients, 323 completed surveys, 1 hospital | Mixed, Adults | Mixed | ICU | p=0.02, overall family satisfaction<br>p=0.007, family satisfaction with care<br>p=0.02, overall patient satisfaction<br>p=0.01, patient satisfaction with care | p=0.12, family satisfaction with decision making<br>p=0.21, patient satisfaction with decision making | |
| Kainiemi 2021[63] | 59% | Finland | Neonates | 61 families, 1 hospital, 1 unit (pre-post-restructuring) | Pre-term infants (<35 weeks) | Unclear | NICU | | Patient perceptions: (mothers and fathers, respectively)<br>p=0.19, p=0.33, overall scores<br>p=0.11, p=0.94, extent staff listen to mothers/fathers<br>p=0.24, p=0.18, participation in baby's care | |

| Citation | QA | Location | Population | Number of patients/ hospitals | Patient type | Type of admission | Level of care | Data that favour single room | Data showing no difference | Data that favour shared room |
|---|---|---|---|---|---|---|---|---|---|---|
| | | | | | | | | | p=0.09, p=0.45, guidance provided by staff met needs p=0.71, p=0.16, opinion considered regarding care of baby p=0.51, p=0.16, mothers/fathers trust in staff caring for baby p=0.28, p=0.92, staff trust in mothers/fathers caring for baby p=0.12, p=0.89, participation in discussions during rounds p=0.51, p=0.41, information given by staff met needs p=0.70, p=0.87, staff offer emotional support | |
| Lawson 2000[69] | 41% | United Kingdom | Adults | 424 patients, 2 hospitals, 4 wards | Psychiatric and Orthopaedic | Unclear | Routine | Patient perceptions (spatially, visually) | | |
| Lester 2014[72] | 63% | United States | Neonates | 403 patients, 1 hospital | Neonates | Emergency | ICU | p<0.001, mother's overall satisfaction p<0.0001, mother's stress p<0.001 mother's satisfaction with family-centred care | | |

| Citation | QA | Location | Population | Number of patients/ hospitals | Patient type | Type of admission | Level of care | Data that favour single room | Data showing no difference | Data that favour shared room |
|---|---|---|---|---|---|---|---|---|---|---|
| | | | | | | | | p<0.0001 mother's involvement in infant care | | |
| Milford 2008[84] | 30% | United States | Neonates | No. of patients unclear, 1 hospital | Neonates | Emergency | ICU | Positive staff perceptions | | |
| Real 2018[105] | 56% | United States | Unclear | 111 patients, 77 nurses, 1 hospital, 1 ward | Cardio-vascular | Unclear | ICU, Routine | p<0.05, patients' satisfaction with design | | |
| Reid 2015[107] | 48% | United Kingdom | Adult, Elderly | 89 patients, 1 hospital relocation | Geriatric | Rehabilitation | Routine | 100% patients prefer private toilet<br>*84.8% of patients in single rooms would prefer single rooms*<br>*37.2% of patients in shared room would prefer single rooms* | | |
| Stevens 2011[121] | 52% | United States | Neonates | 147 patients, 1 hospital | Neonates | Emergency | ICU | p<0.001, parent satisfaction with environment<br>p=0.018, overall parent satisfaction<br>p=0.04, total parent satisfaction score | | |
| Swanson 2013[125] | 37% | United States | Neonates, Carers | 55 parents, 1 hospital | Neonates | Emergency | NICU | p<0.05, nurse perception of facilities<br>p<0.05, practitioners' perceptions of facilities | | |

| Citation | QA | Location | Population | Number of patients/ hospitals | Patient type | Type of admission | Level of care | Data that favour single room | Data showing no difference | Data that favour shared room |
|---|---|---|---|---|---|---|---|---|---|---|
| | | | | | | | | p<0.05, parents' perceptions of facilities | | |
| **Contemporaneous comparison** | | | | | | | | | | |
| Bevan 2016[5] | 59% | United Kingdom | Adults, Elderly | 50 patients, 2 hospitals | Acute illness | Emergency | Routine | Qualitative (privacy, personal control, private toilet)<br>p=0.038, patients perceived a high-level of care | | |
| Boztepe 2017[12] | 63% | Turkey | Children | 130 patients, 1 hospital, 1 ward | Children | Mixed | Routine | | Only 15.4% expected a large or single room | |
| Deitrick 2010[26] | 90% | United States | Adults | 24 patients, 29 HCP, 2 hospitals, 2 wards | Orthopaedic, Neurological, Surgical | Unclear | Routine | Patient preference (privacy) | | |
| de Matos 2020[27] | 63% | Brazil | Unclear | 176 family visitors, 1 hospital, 4 ICU units | Cancer | Unclear | ICU | p=0.02, patient satisfaction<br>Satisfaction of family members | | |
| Douglas 2005[30] | 90% | United Kingdom | Unclear | 785 patients (post discharge), 1 hospital | Surgical, Acute care, Maternity, Geriatric | Unclear | Routine | Patient satisfaction with needs met | | |
| Eberhard-Gran 2000[33] | 59% | Norway | Adults | 160 patients, Unclear (one municipality) | Adults, Pregnant women | Maternity | Routine | OR[a] 18, 2.2-149.1 more likely to be satisfied with care | Satisfaction with rooms<br>Satisfaction with sleep and rest<br>Satisfaction with LOS | |
| Ehrlander 2009[35] | 78% | United States | Adults | 117 patients, 1 hospital | Adults | Unclear | Routine | Patient preference | p=0.309, fear of dying | |

| Citation | QA | Location | Population | Number of patients/ hospitals | Patient type | Type of admission | Level of care | Data that favour single room | Data showing no difference | Data that favour shared room |
|---|---|---|---|---|---|---|---|---|---|---|
| Erdeve 2009[37] | 78% | Turkey | Adults, Neonates | 60 infants, 49 mothers, 2 hospitals | Preterm neonates | Emergency | NICU | | p=0.206, depression scores p=0.06, postpartum depression rate p=0.161, vulnerable child scores p=0.219, parenting stress scores | |
| Harris 2006[50] | 63% | United States | Neonates | 75 HCP, 21 parents, 5 NICU units (SFR=2, open bay=3) | Neonates | Unclear | Level 3, NICU | p<0.05, window view and proximity to infant during sleep Less stressful and less depressing | | |
| Harris 2006[51] | 52% | United States | Neonates | 21 parents, 75 HCPs | Neonates | Maternity | ICU | Less stressful and less depressing, better physical environment. | | |
| Hosseini 2017[52] | 63% | Iran | Adults | 132 patients, 2 hospitals | Medical, Surgical | Unclear | Routine | p<0.001, patients' overall satisfaction p<0.001, patients' total satisfaction | | |
| Janssen 2006[57] | 59% | Canada | Adults | 415 patients, 1 hospital, 2 wards | Pregnant women | Maternity | Routine | p<0.001, patients' overall satisfaction p<0.001, confidence in neonatal care p<0.001, provision of choice p<0.001, physical environment | | |
| Labarère 2004[68] | 70% | France | Adults | 4095 patients, 1 hospital | Mixed | Mixed | Mixed | p<0.01, overall patient experience | | |

| Citation | QA | Location | Population | Number of patients/ hospitals | Patient type | Type of admission | Level of care | Data that favour single room | Data showing no difference | Data that favour shared room |
|---|---|---|---|---|---|---|---|---|---|---|
| Miller 1998[85] | 59% | United States | Adolescents, Adults | 94 patients, 1 hospital | Inpatients, Outpatients | Unclear | Routine | % patients overall stating ideal rooming arrangements<br>% inpatients stating ideal rooming arrangements<br>% female inpatients stating ideal rooming arrangements<br>% inpatients aged 15 to 17 and 18 to 21 stating ideal rooming arrangements<br>% female outpatients stating ideal rooming arrangements | % outpatients aged 12 to 14 stating ideal rooming arrangements | % male inpatients and outpatients stating ideal rooming arrangements<br>% inpatients aged 12 to 14 stating ideal rooming arrangements<br>% outpatients stating ideal rooming arrangements<br>% outpatients aged 15 to 17 and 18 to 21 stating ideal rooming arrangements |
| Morgan 2010[87] | 44% | UK, US | Children | 146 patients, 114 HCP, 2 hospitals | Children | Mixed | Routine | | | % patient preference |
| Nahas 2016[89] | 56% | United Kingdom | Adults, Elderly | 60 patients, 2 hospitals | Orthopaedic (elective hip/ knee arthroplasty) | Elective | Routine | $p$=0.014, feeling of safety<br>Qualitative (privacy, security, pain management, cleanliness) | $p$=0.061, overall patient satisfaction | |
| Nash 2021[90] | 63% | Australia | Adults | 602 patients, 4 hospitals | Unclear | Unclear | Routine | | | Patient preference |
| Nassery 2019[91] | 90% | Sweden | Children | 13 interviews (9 individual parents, 4 pairs of | Children | Unclear | Mixed | Parents preference | | |

| Citation | QA | Location | Population | Number of patients/ hospitals | Patient type | Type of admission | Level of care | Data that favour single room | Data showing no difference | Data that favour shared room |
|---|---|---|---|---|---|---|---|---|---|---|
| | | | | parents), 1 hospital | | | | | | |
| Olson 1992[93] | 52% | United States | Adults | 351 patients, 28 HCP, 1 hospital | Pregnant women | Maternity | Routine | Nurse preference<br>*Mother satisfaction* | | |
| Pease 2002[96] | 48% | United Kingdom | Unclear | 50 patients, 1 hospital | Oncologic, Terminal | Unclear | Routine | Family preference | | Patient preference |
| Persson 2012[97] | 90% | Sweden | Adults, Elderly | 16 patients, 10 nurses, 1 hospital, 2 wards | Orthopaedic, Surgical | Unclear | Routine | | | Qualitative (security and safety) |
| Persson 2015[98] | 90% | Sweden | Adults | 16 patients, 1 hospital | Surgical | Unclear | Routine | | | Qualitative (security, company, not isolated) |
| Pineda 2012[100] | 70% | United States | Neonates | 81 patients, 1 hospital | Premature neonates | Emergency | NICU | | p=0.512, maternal depression<br>p=0.152, trait anxiety<br>p=0.830, state anxiety<br>p=0.071, life stress<br>p=0.603, avoidance coping<br>p=0.967, emotion-oriented coping<br>p=0.506, task-oriented coping<br>p=0.951, social support | p=0.040[a], stress levels |
| Roos 2020[108] | 90% | Norway | Adults | 39 patients, 1 hospital relocation | Internal medicine, Surgical, Maternity | Maternity, Unclear | Routine | | | Satisfaction for older/bedridden patients |

| Citation | QA | Location | Population | Number of patients/ hospitals | Patient type | Type of admission | Level of care | Data that favour single room | Data showing no difference | Data that favour shared room |
|---|---|---|---|---|---|---|---|---|---|---|
| Rowlands 2008[110] | 90% | United Kingdom | Adults | 12 patients, 1 hospital | Adults with advanced cancer | Unclear | Routine | | Qualitative (desire for choice of room) | |
| Stelwagen 2021[120] | 100% | Netherlands | Neonates | 36 parents, 1 hospital Before and after relocation | Neonates | Emergency | ICU | Qualitative (privacy, safety, homeliness, feelings of central engagement with child care) | | |
| Tandberg 2019[127] | 67% | Norway | Infants | 77 infants, 132 parents, 2 hospitals | Infants | Emergency | ICU | Mothers:<br>p=0.005, depression at day 14<br>p=0.04, anxiety at day 14<br>p=0.0001, role alteration at day 14<br>p=0.06, role alteration at discharge<br><br>Fathers:<br>p=0.06, environmental stress at day 14<br>p=0.003, role alteration at day 14<br>p=0.003, environmental stress at discharge<br>p=0.004, role alteration at discharge | Mothers:<br>p=0.12 Maternal distress at day 14<br>p=0.43 depression, and p=0.48, anxiety at discharge<br>p=0.13, distress at discharge<br>p=0.65, depression and p=0.54, anxiety at 4-month corrected age<br>p=0.60, distress at 4-month corrected age<br>p=0.62, dysfunctional interaction with child<br>p=0.23, perceived child to be difficult<br>p=0.42, stress<br>p=0.51, attachment | |

| Citation | QA | Location | Population | Number of patients/ hospitals | Patient type | Type of admission | Level of care | Data that favour single room | Data showing no difference | Data that favour shared room |
|---|---|---|---|---|---|---|---|---|---|---|
| | | | | | | | | | Fathers:<br>p=0.17, depression and p=0.25, anxiety at day 14<br>p=0.57, depression and p=0.73, anxiety at discharge<br>p=0.92, depression and p=0.11, anxiety at 4-month corrected age<br>p=0.16, dysfunctional interaction with child<br>p=0.77, perceived child to be difficult<br>p=0.68, stress<br>p=0.49, attachment | |
| Van Veenendaal 2022[138] | 70% | Netherlands | Neonates | 182 parents, 3 hospitals | Fathers of neonates | Emergency | ICU | p=0.001[a], stress overall<br>p=0.011[a], stress related to environment<br>p<0.001[a], stress related to role alteration | p=0.83[a], depression and anxiety<br>p=0.26[a], self-efficacy<br>p=0.27[a], impaired parent-newborn bonding<br>p=0.32, satisfaction with care | |
| Watson 2014[144] | 44% | Canada | Neonates | 85 families,<br>1 hospital | Neonates | Emergency | NICU | p=0.008, privacy<br>p=0.0001, comfort<br>p=0.009, interaction with other families | p=0.05, getting to know baby<br>p=0.05, feeling irritable, anxious, depressed or sad | |

| Citation | QA | Location | Population | Number of patients/ hospitals | Patient type | Type of admission | Level of care | Data that favour single room | Data showing no difference | Data that favour shared room |
|---|---|---|---|---|---|---|---|---|---|---|
| | | | | | | | | p=0.04. confidence feeding baby<br>p=0.04, easy to comfort baby<br>*p=0.003, family adjusted to having the baby home* | p=0.05, satisfied with care baby received | |
| **Economic analysis** | | | | | | | | | | |
| Boardman 2011[8] | 91% | Canada | Unclear | 537 beds,<br>1 hospital | Mixed | Mixed | Mixed | Patients and willingness to pay for single over shared rooms | | |
| **Evidence synthesis** | | | | | | | | | | |
| Bradbury-Jones 2013[14] | SLR 86% | International | Adults | NR | Mixed, Vulnerable, Learning difficulties | Unclear | Unclear | | Mixed views among patients with learning disabilities | |
| Dowdeswell 2004[31] | SLR 36% | International | Unclear | Unclear | Mixed | Mixed | Mixed | Quicker mobility recovery<br>Sense of self-reliance<br>Personal control leads to happier patients. | | |
| OECD WHO 2019[92] | Report 14% | Europe | NR | NR | Mixed | Mixed | Mixed | 12 studies showed single rooms positively affect patient satisfaction | 4 studies showed no difference | 1 study showed single rooms don't positively affect patient satisfaction |
| Søndergaard 2022[118] | SLR 91% | International | NR | NR | Acute, Surgical, Internal medicine | Unclear | Routine | | | Communication and interaction with kindred spirits was appreciated |

| Citation | QA | Location | Population | Number of patients/ hospitals | Patient type | Type of admission | Level of care | Data that favour single room | Data showing no difference | Data that favour shared room |
|---|---|---|---|---|---|---|---|---|---|---|
| | | | | | | | | | | Bedridden / older patients were less satisfied with single rooms. |
| Taylor 2018[129] | SLR 91% | International | NR | NR | Mixed | Mixed | Mixed | Patient perceptions of dignity | | |
| Voigt 2018[141] | SLR 86% | International | NR | NR | NR | Unclear | Routine | 1 study found advantages for feelings of safety<br>1 study found advantages for patient preference<br>1 study found advantage or no difference for patient preference | 1 study found mixed findings for feelings of safety<br>All studies found mixed findings regarding concern for others<br>1 study found mixed findings for patient preference | |

#### Table 13. Summary of studies reporting data on patient monitoring and safeguarding

| Citation | QA | Location | Population | Number of patients/ hospitals | Patient type | Type of admission | Level of care | Data that favour single room | Data showing no difference | Data that favour shared room |
|---|---|---|---|---|---|---|---|---|---|---|
| **Before and after a hospital relocation plus Contemporaneous comparison** | | | | | | | | | | |
| Maben 2016[77] | 67% | United Kingdom | Unclear | 32 patients, 21 HCP, 1 hospital relocation, 2 control hospitals | Mixed | Unclear | Mixed | | *Qualitative (regular visits by staff to single-rooms)* | |
| **Before and after a hospital relocation** | | | | | | | | | | |
| Real 2018[105] | 56% | United States | Unclear | 111 patients, 77 nurses, 1 hospital, 1 ward | Cardio-vascular | Unclear | ICU, Routine | | Staffing ratio | |
| Jansen 2021[55] | 63% | Netherlands | Neonates | 712 patients 1 hospital, 2 units | Premature neonates | Maternity care | NICU | | Nurse-to-patient ratio | |
| Jones 2016[58] | 100% | Australia | Neonates | 66 mothers, 51 nurses, 1 hospital relocation | Adults, Mothers of premature neonates, Nurses | Maternity | NICU | | | Nurse perception (parallel patient interactions, get caught in single rooms so can't attend to other families) |
| Jung 2022[62] | 67% | South Korea | Adults | 901 patients, 1 hospital | Mixed | Unclear | ICU | | Nurse-to-patient ratio | |
| **Contemporaneous comparison** | | | | | | | | | | |
| Bevan 2016[5] | 59% | United Kingdom | Adults, Elderly | 50 patients, 2 hospitals | Acute medical illness | Emergency | Routine | Patient perceptions (isolation) | | |
| Bodack 2016[10] | 55% | Germany | Neonates | 35 pairs of parents | Premature neonates | Maternity care | NICU | Somewhat less frequent | | |

| Citation | QA | Location | Population | Number of patients/ hospitals | Patient type | Type of admission | Level of care | Data that favour single room | Data showing no difference | Data that favour shared room |
|---|---|---|---|---|---|---|---|---|---|---|
| | | | | | | | | adequate monitoring | | |
| Bracco 2007[13] | 74% | Canada | Adults | 2522 patients (of whom 207 known MRS carriers at admission), 1 hospital, 1 ward | Mixed, Post surgery, Medical admission | Mixed | ICU | *Standard nurse-to-patient ratio 1:2* | | |
| Ehrlander 2009[35] | 78% | United States | Adults | 117 patients, 1 hospital | Mixed | Unclear | Routine | p=0.025, patient perception of nurse availability | | |
| Deitrick 2010[26] | 90% | United States | Adults | 24 patients, 29 HCP, 2 hospitals, 2 wards | Orthopaedic, Neurological, Surgical | Unclear | Routine | Better response to call lights. More visits to anticipate patient needs. | | |
| Hosseini 2017[52] | 63% | Iran | Adults | 132 patients, 2 hospitals | Medical, Surgical | Unclear | Routine | p=0.19, access to nurses | | |
| Julian 2015[61] | 78% | United States | Neonates | 1823 patients 1 hospital, 1 unit | Neonates | Mixed | NICU | | Nurse-to-patient ratio | |
| Nahas 2016[89] | 56% | United Kingdom | Adults, Elderly | 60 patients, 2 hospitals | Orthopaedic (elective hip/ knee arthroplasty) | Elective | Routine | | p=0.244, response to call bell | |
| **Early vs late response to new unit design** | | | | | | | | | | |

Health Economics Unit

| Citation | QA | Location | Population | Number of patients/ hospitals | Patient type | Type of admission | Level of care | Data that favour single room | Data showing no difference | Data that favour shared room |
|---|---|---|---|---|---|---|---|---|---|---|
| Ferri 2015[41] | 100% | Canada | Adults | 39 HCPs, of which 13 nurses, 7 respiratory therapists, 5 HCPS (other), 6 physicians, 4 family members 4 support staff 1 hospital, 1 unit | Unclear | Unclear | ICU | 75 negative comments on shared-room design | | Qualitative (less safety concerns related to distance between patient and care provider) |

#### Table 14. Summary of studies reporting views on patient confidentiality

| Citation | QA | Location | Population | Number of patients/ hospitals | Patient type | Type of admission | Level of care | Data that favour single room | Data showing no difference | Data that favour shared room |
|---|---|---|---|---|---|---|---|---|---|---|
| **Before and after a hospital relocation plus Contemporaneous comparison** | | | | | | | | | | |
| Maben 2016[77] | 67% | United Kingdom | Unclear | 32 patients, 21 HCP, 1 hospital relocation, 2 control hospitals | Mixed | Unclear | Mixed | Qualitative (confidentiality) | | |
| **Before and after a hospital relocation** | | | | | | | | | | |
| Ferri 2015[41] | 100% | Canada | Adults | 39 HCPs, of which 13 nurses, 7 respiratory therapists, 5 HCPS (other), 6 physicians, 4 family members 4 support staff, 1 hospital | Unclear | Unclear | ICU | Qualitative (patient perceptions of confidentiality) | | |
| Jones 2016[58] | 100% | Australia | Neonates | 66 mothers, 51 nurses, 1 hospital relocation | Adults, Mothers of premature neonates, Nurses | Maternity | NICU | Qualitative (nurse perceptions of confidentiality, facilitating care) | | |
| Florey 2009[42] | 44% | United Kingdom | Adults | 80 patients, 2 hospitals, 1 move | Medical and surgical, Adults | Unclear | Routine | $p<0.001$ ability to have confidential discussions | | |
| Real 2018[105] | 56% | United States | Unclear | 111 patients, 77 nurses, 1 hospital, 1 ward | Cardio-vascular | Unclear | ICU, Routine | | Patient satisfaction with confidentiality | |

| Citation | QA | Location | Population | Number of patients/ hospitals | Patient type | Type of admission | Level of care | Data that favour single room | Data showing no difference | Data that favour shared room |
|---|---|---|---|---|---|---|---|---|---|---|
| Roos 2020[108] | 90% | Norway | Adults | 39 patients, 1 hospital relocation | Internal medicine, Surgical, Maternity | Maternity, Unclear | Routine | Qualitative (patient perceptions of confidentiality) | | |
| **Contemporaneous comparison** | | | | | | | | | | |
| Bodack 2016[10] | 56% | Germany | Neonates | 35 pairs of parents of 40 neonates, 1 hospital | Neonates | Emergency | ICU | Qualitative (easier to guarantee confidentiality) | | |
| Bevan 2016[5] | 59% | United Kingdom | Adults, Elderly | 50 patients, 2 hospitals | Acute illness | Emergency | Routine | Qualitative (patient perceptions of confidentiality) | | |
| Hosseini 2017[52] | 63% | Iran | Adults | 2 hospitals 132 patients | Adults, Medical or surgical | Unclear | Routine | $p<0.001$ comfortable discussing personal problems | | |
| Malcolm 2005[78] | 80% | New Zealand | Adolescents, Adults | 12 former patients | Mixed surgery, orthopaedic, medical, obstetric, ENT | Mixed | Routine | Qualitative (patients in shared rooms felt a lack of privacy and confidentiality which affected relationships with other patients) | | |
| **Evidence synthesis** | | | | | | | | | | |
| OECD WHO 2019[92] | Report 14% | Europe | NR | NR | Mixed | Mixed | Mixed | $p<0.05$, improved patient confidentiality | | |

### Table 2. Summary of studies reporting data on availability of beds, space requirements, and capital costs

| Citation | QA | Location | Population | Number of patients/ hospitals | Patient type | Type of admission | Level of care | Data that favour single room | Data showing no difference | Data that favour shared room |
|---|---|---|---|---|---|---|---|---|---|---|
| **Before and after a hospital relocation plus contemporaneous comparison** | | | | | | | | | | |
| Maben 2016[77] | 67% | United Kingdom | Unclear | 32 patients,<br>21 HCP,<br>1 hospital relocation,<br>2 control hospitals | Mixed | Unclear | Mixed | | | Higher space requirement for single-bed wards<br>Building costs per bed |
| **Before and after a hospital relocation** | | | | | | | | | | |
| Darley 2018[24] | 56% | United Kingdom | Unclear | 1 hospital relocation | Unclear | Unclear | Routine | Ward closures per year<br>Bed days lost per 100,000 | | |
| Domanico 2011[29] | 63% | United States | Neonates | 162 patients (PEMRs 2/3=150, PEMRs 4=12), 1 hospital, 2 units | Paediatric | NR | NICU | Number of patients accommodated;<br>Total space | | |
| Jones 2016[58] | 100% | Australia | Neonates | 66 mothers, 51 nurses, 1 hospital relocation | Adults, Mothers of premature neonates, Nurses | Maternity | NICU | Capacity | | Room space |
| Jongerden 2013[59] | 67% | Netherlands | Adults | 387 patients, 323 completed surveys,<br>1 hospital Before and after relocation | Mixed, Adults | Mixed | ICU | Number of beds<br>Space per bed | | |
| Jung 2022[62] | 67% | Korea | Adults | 901 patients,<br>1 hospital Before and after renovation | Adult, mixed | Unclear | ICU | | Number of isolated rooms | Number of beds |
| Kosuge 2013[67] | 41% | Japan | Unclear | 555 beds,<br>1 hospital | Surgical, Internal medicine | Unclear | Routine | Number of beds (working, general, per nursing unit)<br>Wards in total | Number of beds (tuberculosis) | Number of beds (mental, cases of floor transfer) |

| Citation | QA | Location | Population | Number of patients/ hospitals | Patient type | Type of admission | Level of care | Data that favour single room | Data showing no difference | Data that favour shared room |
|---|---|---|---|---|---|---|---|---|---|---|
| | | | | | | | | | Total number of people / day and the wards | |
| Lawson 2000[69] | 41% | United Kingdom | Adults | 424 patients, 2 hospitals, 4 wards (pre-/post-relocation) | Orthopaedic | Unclear | Routine | Number of beds | | |
| Real 2018[105] | 56% | United States | Unclear | 111 patients, 77 nurses, 1 hospital, 1 ward | Cardio-vascular | Unclear | ICU, Routine | Qualitative (larger rooms promote more space for family) | | |
| Rosbergen 2020[109] | 74% | Australia | Adults, Elderly | 73 patients, 1 hospital relocation | Stroke, Neurological | Emergency, Rehabilitation | Routine | p=0.007, number of single bedrooms in acute stroke unit/ neurology<br>p<0.001, number of single bedrooms in inpatient rehab unit<br>Ward length<br>Total communal floor space | Number of any bedrooms, acute stroke unit/ neurology | Number of any bedrooms, inpatient rehabilitation unit |
| **Contemporaneous comparison** | | | | | | | | | | |
| Julian 2015[61] | 78% | United States | Neonates | 1823 patients, 1 hospital, 1 unit | Neonates | Mixed | NICU | | | Bed capacity |
| Kinnula 2008[64] | 63% | Finland | Children | 1927 patients, 1 hospital | Children, infectious disease | Mixed | Routine | Single rooms usage (approx. 90%) | Number of rooms | |
| Kinnula 2012[65] | 67% | Finland, Switzerland | Children | 5119 patients, 3 hospitals, 4 wards | Children, mixed | Mixed | Routine | | | Bed capacity |
| Pineda 2012[100] | 70% | United States | Neonates | 81 patients, 1 hospital | Premature neonates | Emergency | NICU | | | Number of beds; Room/ward area |

| Citation | QA | Location | Population | Number of patients/ hospitals | Patient type | Type of admission | Level of care | Data that favour single room | Data showing no difference | Data that favour shared room |
|---|---|---|---|---|---|---|---|---|---|---|
| Quach 2018[104] | 59% | Canada, United States | Children | 83,334 patient-days, 2 hospitals | Children | Mixed | Mixed | | | Bed capacity |
| Stelwagen 2021[120] | 100% | Netherlands | Neonates | 36 parents, 1 hospital | Neonates | Emergency | ICU | Capacity; Room/ward area | | |

Health Economics Unit

#### Table 16. Summary of studies reporting data on length of stay

| Citation | QA | Location | Population | Number of patients/ hospitals | Patient type | Type of admission | Level of care | Data that favour single room | Data showing no difference | Data that favour shared room |
|---|---|---|---|---|---|---|---|---|---|---|
| **Before and after a hospital relocation plus contemporaneous comparison** | | | | | | | | | | |
| Maben 2016[77] | 67% | United Kingdom | Unclear | 32 patients, 21 HCP, 1 hospital relocation, 2 control hospitals | Mixed | Unclear | Mixed | LOS (per 1,000 patient-days): new hospital older people's ward, control new-build hospital older people's ward, steady-state control hospital medical assessment unit | | LOS (per 1,000 patient-days): new hospital assessment unit, control new-build hospital medical assessment unit, steady-state control hospital older people's ward |
| **Before and after a hospital relocation** | | | | | | | | | | |
| Blandfort 2019[6] | 67% | Denmark | Elderly | 964 patients, 2 hospitals | Geriatric, Dementia | Elective | Routine | p=0.35, median LOS | | |
| Blandfort 2019[7] | 67% | Denmark | Elderly | 1014 patients, 2 hospitals | Geriatric, Dementia | Elective | Routine | Fewer cases with LOS ≥ 14 days | Minimum LOS | Maximum LOS |
| Cantoni 2009[16] | 67% | Switzerland | Adults | 227 patients, 1 hospital | Stem cell transplant | Elective | Routine | LOS<br>Duration of catheterisation<br>Number of patients catheterised | | |
| Carter 2008[18] | 33% | United States | Neonates | 53 parents, 1 hospital Before and after relocation | Neonates | Emergency | NICU | LOS | | |
| Davis 2019[25] | 67% | Australia | Adults | 1569 patients, 1 hospital relocation | Orthopaedic | Elective | Routine | | p=0.698, ward LOS<br>p=0.226, hospital LOS | |
| Domanico 2010[28] | 63% | United States | Neonates | 161 caregivers, 1 hospital, 2 units | Paediatric | NR | NICU | LOS | | |
| Domanico 2011[29] | 63% | United States | Neonates | 162 patients (PEMRs 2/3=150, | Paediatric | NR | NICU | | p=0.340, LOS for PEMR 2 and 3 patients | |

| Citation | QA | Location | Population | Number of patients/ hospitals | Patient type | Type of admission | Level of care | Data that favour single room | Data showing no difference | Data that favour shared room |
|---|---|---|---|---|---|---|---|---|---|---|
| | | | | PEMRs 4=12), 1 hospital, 2 units | | | | | p=0.890, LOS for PEMR 4 patients | |
| Erickson 2011[38] | 67% | United States | Neonates | 73 patients, 1 hospital Before and after relocation | Preterm neonates | Emergency | NICU | | *p*=0.73, LOS | |
| Gregersen 2021[46] | 70% | Denmark | Elderly | 446 patients, 1 hospital relocation | Geriatric | Unclear | Routine | | *p*=0.50, hospital LOS | |
| Harris 2004[49] | 74% | Canada | Adults | 976 patients, 1 hospital, Before and after new unit established | Pregnancy | Maternity | Routine | p<0.001, total LOS<br>p<0.001, postpartum LOS | | p=0.01, length of first stage labour<br>p=0.002, length of second stage labour<br>p=0.002, intrapartum LOS |
| Hourigan 2018[53] | 63% | United States | Neonates | 32 patients, 1 hospital Before and after relocation | Neonates | Emergency | ICU | | p=0.52, LOS | |
| Jansen 2021[55] | 63% | Netherlands | Neonates | 712 patients, 1 hospital, 2 units relocation | Premature neonates | Maternity | NICU | | p=0.36, hospital LOS | |
| Jongerden 2013[59] | 67% | Netherlands | Adults | 387 patients, 323 completed surveys, 1 hospital Before and after relocation | Mixed, Adults | Mixed | ICU | | p=0.25, ICU LOS: family<br>p=0.11, ICU LOS: patients<br>p=0.25, hospital LOS: family<br>p=0.60, hospital LOS: patients | |
| Jung 2022[62] | 67% | Korea | Adults | 901 patients, 1 hospital Before and after renovation | Adult, mixed | Unclear | ICU | | p=0.575, ICU LOS | |

| Citation | QA | Location | Population | Number of patients/ hospitals | Patient type | Type of admission | Level of care | Data that favour single room | Data showing no difference | Data that favour shared room |
|---|---|---|---|---|---|---|---|---|---|---|
| Kainiemi 2021[63] | 59% | Finland | Neonates | 61 families, 1 hospital, 1 unit (pre-post-restructuring) | Pre-term infants (<35 weeks) | Unclear | NICU | | p=0.1784, hospital LOS | |
| Kosuge 2013[67] | 41% | Japan | Unclear | 555 beds, 1 hospital | Surgical, Internal medicine | Unclear | Routine | Average hospital LOS (surgery, internal medicine) | | |
| Lawson 2000[69] | 41% | United Kingdom | Adults | 424 patients, 2 hospitals, 4 wards (pre-/post-relocation) | Psychiatric and Orthopaedic | Unclear | Routine | p<0.05, hospital LOS (orthopaedic patients not undergoing operation)<br>Hospital LOS overall (psychiatric patients)<br>ICU LOS (psychiatric patients) | Hospital LOS (orthopaedic patients undergoing operation) | |
| Milford 2008[84] | 30% | United States | Neonates | No. of patients unclear, 1 hospital Before and after relocation | Neonates | Emergency | ICU | Average LOS | | |
| Monson 2018[86] | 78% | United States | Neonates | 90 preterm infants, 15 term-born control infants, 1 hospital | Preterm neonates | Emergency | NICU | | p=0.81, LOS | |
| Puumala 2020[102] | 67% | United States | Neonates | 9995 patients, 1 hospital Before and after relocation | Neonates | Emergency | ICU | p=0.02, LOS for extremely preterm infants<br>p<0.0001, LOS for very preterm infants | p=0.71, LOS for moderately pre-term | p<0.0001, overall median hospital LOS<br>p<0.0001, LOS for term/post term infants |
| Pyrke 2017[103] | 59% | Canada | Adults | 47 patients, 1 hospital relocation | Psychiatric | Emergency | Routine | | *p*=0.832, LOS | |

| Citation | QA | Location | Population | Number of patients/ hospitals | Patient type | Type of admission | Level of care | Data that favour single room | Data showing no difference | Data that favour shared room |
|---|---|---|---|---|---|---|---|---|---|---|
| Sadatsafavi 2019[112] | 100% | United States | Neonates | NR, 1 hospital (theoretical) | Neonates | NR | ICU | Mean benefit–cost ratio 1.298 (95% CI: 1.282–1.315) when reduced LOS considered | | |
| Singh 2015[116] | 70% | United Kingdom | Adults, Elderly | 1749 patients, 1 hospital relocation | Internal medicine, Geriatric | Mixed | Routine | p<0.01, LOS | | |
| Stevens 2014[123] | 44% | United States | Neonates | 73 patients, 1 hospital | Neonates | Emergency | ICU | | | p=0.0052, hospital LOS |
| Teltsch 2011[131] | 67% | Canada | Adults | 19343 patients, 2 hospitals, Before and after relocation or control | Adults | Unclear | ICU | | | Average ICU LOS (year 2000, 2001, 2002, 2003, 2004, 2005, and total) |
| van der Hoeven 2022[135] | 63% | Netherlands | Infants | 1293 infants, 1 hospital Before and after relocation | Infants | Unclear | ICU | | p=0.49, hospital LOS | |
| van Veenendaal 2020[137] | 70% | Netherlands | Neonates | 1152 infants, 1 hospital Before and after relocation | Neonates | Emergency | ICU | p=0.016, LOS | | |
| Vietri 2004[139] | 59% | United States | Adults | 261 Adults, 1 hospital Before and after relocation | Mixed | Unclear | ICU | | p=NS, ICU LOS | |
| **Contemporaneous comparison** | | | | | | | | | | |
| Bodack 2016[10] | 56% | Germany | Neonates | 35 pairs of parents of 40 neonates, 1 hospital | Neonates | Emergency | ICU | LOS | | |
| Bracco 2007[13] | 74% | Canada | Adults | 2522 patients (of whom 207 known MRS carriers at | Mixed, Post surgery, Medical admission | Mixed | ICU | *LOS in the same bed* | LOS | |

| Citation | QA | Location | Population | Number of patients/ hospitals | Patient type | Type of admission | Level of care | Data that favour single room | Data showing no difference | Data that favour shared room |
|---|---|---|---|---|---|---|---|---|---|---|
| | | | | admission), 1 hospital, 1 ward | | | | | | |
| Caruso 2014[19] | 74% | Brazil | Adults | 1253 patients, 1 hospital | Adults | Mixed | ICU | | p=0.44, ICU LOS | |
| Deitrick 2010[26] | 90% | United States | Adults | 24 patients, 29 HCP, 2 hospitals, 2 wards | Orthopaedic, Neurological, Surgical | Unclear | Routine | LOS | | |
| Douglas 2005[30] | 90% | United Kingdom | Unclear | 785 patients (post discharge), 1 hospital | Surgical, Acute care, Maternity, Geriatric | Unclear | Routine | | LOS | |
| Erdeve 2008,[36] Erdeve 2009[37] | 74% | Turkey | Adults, Neonates | 60 infants, 49 mothers, 1 hospital | Preterm neonates | Emergency | NICU | | p=0.929, NICU LOS | |
| Felice Tong 2018[40] | 78% | Australia | Adults | 185 patients, 1 hospital | Orthopaedic | Elective | Routine | | p=0.36, overall LOS p=0.73, LOS for total hip arthroplasty p=0.55, LOS for knee arthroplasty | |
| Grundt 2021[47] | 67% | Norway | Neonates | 77 patients, 66 mothers, 2 hospitals, 2 units | Premature neonates | Maternity | NICU | | p=0.16, LOS | |
| Harris 2006[50] | 63% | United States | Neonates | 75 HCP, 21 parents, 5 NICU units (SFR=2, open bay=3) | Neonates | Unclear | Level 3, NICU | Patient transfers | | Average LOS Average discharges |
| Harris 2006[51] | 52% | United States | Neonates | 21 parents, 75 HCPs | Neonates | Maternity | ICU | | | Average LOS |
| Hyun 2021[54] | 78% | South Korea | Adults | 666 patients, 1 hospital | Respiratory, COVID-19 | Emergency | ICU | p=0.001, hospital LOS | | |
| Kinnula 2008[64] | 63% | Finland | Children | 1927 patients, 1 hospital | Children, infectious disease | Mixed | Routine | | hospital LOS | |

| Citation | QA | Location | Population | Number of patients/ hospitals | Patient type | Type of admission | Level of care | Data that favour single room | Data showing no difference | Data that favour shared room |
|---|---|---|---|---|---|---|---|---|---|---|
| Kinnula 2012[65] | 67% | Finland, Switzerland | Children | 5119 patients, 3 hospitals, 4 wards | Children, mixed | Mixed | Routine | | | Mean hospital LOS |
| Knight 2016[66] | 59% | United Kingdom | Elderly | 100 patients, 2 hospitals | Geriatric, Dementia | Mixed | Routine | | | p=0.001, overall LOS p=0.01, LOS (patients who experienced an inpatient fall) |
| Labarère 2004[68] | 70% | France | Adults | 4095 patients, 1 hospital | Mixed | Mixed | Mixed | | hospital LOS | |
| Lehtonen 2020[71] | 74% | International | Neonates | 4662 patients, 331 units | Preterm neonates | Emergency | ICU | Overall LOS OR[a] −3.4 (−4.7 to −3.1) | | |
| Mattner 2007[79] | 74% | Germany | Adults | 336 patients, 1 hospital | Cardiovascular Adults | Mixed | ICU | p=0.004, LOS | | |
| Vohr 2017[140] | 67% | United States | Neonates | 651 patients, 1 hospital Before and after relocation | Neonates | Emergency | NICU | | | p=0.07, hospital LOS |
| Tandberg 2019[128] | 67% | Norway | Neonates | 77 patients, 2 hospitals | Neonates | Emergency | ICU | | p=0.16, LOS | |
| Lester 2014[72] | 63% | United States | Neonates | 403 patients, 1 hospital | Neonates | Emergency | ICU | | p=0.382, LOS | |
| Lester 2016[73] | 59% | United States | Neonates | 216 patients, 1 hospital | Premature neonates | Maternity | ICU | | p=0.06, LOS | |
| Zaal 2013[145] | 67% | Netherlands | Older Adults | 156 patients 1 hospital | Older Adults with dementia | Mixed | ICU | | p=0.56, LOS | |
| **Evidence synthesis** | | | | | | | | | | |
| OECD WHO 2019[92] | 14% | Europe | NR | NR | Mixed | Mixed | Mixed | | p=NS. LOS | |
| Voigt 2018[141] | 86% | International | NR | NR | NR | Unclear | Routine | | LOS | |

#### Table 17. Summary of studies reporting data on costs of care

| Citation | QA | Location | Population | Number of patients/ hospitals | Patient type | Type of admission | Level of care | Data that favour single room | Data showing no difference | Data that favour shared room |
|---|---|---|---|---|---|---|---|---|---|---|
| **Before and after a hospital relocation plus contemporaneous comparison** | | | | | | | | | | |
| Maben 2015[76] | 78% | United Kingdom | Unclear | 24 staff, 32 patients, 1 hospital (relocated), 2 control hospitals | All patients in hospital | Mixed | Mixed | | Cost impact (changes in falls, LOS, medication errors, hospital-acquired infections) | |
| Maben 2016[77] | 67% | United Kingdom | Unclear | 32 patients, 21 HCP, 1 hospital relocation, 2 control hospitals | Mixed | Unclear | Mixed | | | Cleaning costs per bed<br>Nursing staff full-time equivalent<br>Nursing staff costs |
| **Before and after a hospital relocation** | | | | | | | | | | |
| Davis [25]2019 | 67% | Australia | Adults | 1569 patients, 1 hospital relocation | Orthopaedic | Elective | Routine | | p=0.311, discharge to home<br>p=0.406, transfer to other facility | |
| Harris 2004[49] | 74% | Canada | Adults | 976 patients, 1 hospital, Before and after new unit established | Pregnancy | Maternity | Routine | Reduction in overall staffing costs after opening single-room maternity care | | |
| Milford 2008[84] | 30% | United States | Neonates | No. of patients unclear, 1 hospital | Neonates | Emergency | ICU | Cost savings due to reduced LOS | | |
| Reed 1986[106] | 10% | United States | Adults | No. of patients unclear, 1 hospital | Pregnant women | Maternity care | Routine | Number of staff required | | |
| Sadatsafavi 2019[112] | 100% | United States | Neonates | 1 hospital (theoretical) | Neonates | NR | ICU | Investment justifiable when direct costs considered, mean benefit–cost ratio 1.794 (1.783–1.804)<br>Investment justifiable when LOS considered | | Investment not justifiable when nosocomial infections considered, mean benefit–cost ratio 0.730 (0.724-0.735) |

| | | | | | | | | | | |
|---|---|---|---|---|---|---|---|---|---|---|
| Singh 2015[116] | 70% | United Kingdom | Adults, Elderly | 1749 patients, 1 hospital relocation | Internal medicine, Geriatric | Mixed | Routine | | p=0.74, discharge to home<br>p =0.21, discharge to new care home | |
| Stevens 2012[122] | 44% | United States | Neonates | 73 patients, 1 hospital Before and after relocation | Neonates | Emergency | ICU | Direct cost (infants with equal comorbidities, duration of hospitalisation) | Costs per square foot | $p^a$=statistically significant, need for nursing and all unit staff |
| Stevens 2014[123] | 44% | United States | Neonates | 73 patients, 1 hospital | Neonates | Emergency | ICU | p<0.0001, lower costs for supplies<br>p<0.0001, lower depreciation in costs<br>Full adjustment of the model shows a cost advantage for SFR | p=0.2316, total direct costs<br>p=0.1551, other costs<br><br>General linear model:<br>p=0.2854, admission<br>p=0.2485, severity<br>p=0.2806, duration of respiratory support | p=0.0373, direct costs for NICU labour<br>p=0.0002, direct costs for other labour costs (therapies, radiology, pharmacy) |
| **Contemporaneous comparison** | | | | | | | | | | |
| Apple 2014[4] | 52% | Sweden | Unclear | 81 HCP, 3 ICUs | Mixed | Unclear | ICU | | | Number of staff equired |
| Boardman 2011[8] | 91% | Canada | Unclear | 537 beds, 1 hospital | Mixed | Mixed | Mixed | Reduced transfers and waiting time<br>Net social benefits taking into account upfront and ongoing costs and annual benefits | | Cost of a bed per day<br>Up-front land and construction costs<br>On-going annual maintenance, housekeeping, operating, additional nursing and phsycian costs |
| Felice Tong 2018 [40] | 78% | Australia | Adults | 185 patients, 1 hospital | Orthopaedic | Elective | Routine | $p=0.002^u$, $p=0.002^m$ discharge to rehabilitation | | |
| Harris 2006[50] | 63% | United States | Neonates | 75 HCP, 21 parents, 5 NICU units (SFR=2, open bay=3) | Neonates | Unclear | Level 3, NICU | | | Construction costs per square foot |
| Harris 2006[51] | 63% | Canada | Adults | 976 patients, 1 hospital, Before and | Pregnancy | Maternity | Routine | | | Average costs per square foot[a] |

| | | | | | | | | | | |
|---|---|---|---|---|---|---|---|---|---|---|
| | | | | after new unit established | | | | | | |
| Knight 2016[66] | 59% | United Kingdom | Elderly | 100 patients, 2 hospitals | Geriatric, Dementia | Mixed | Routine | | p=0.17, discharged to home p=0.19, discharged to new care home | |
| Sadatsafavi 2016[111] | 100% | Canada | Unclear | 8811 patient-days, 1 hospital | Medical and surgical | Unclear | ICU | Costs due to hospital acquired infection | | Construction and operating costs |
| **Evidence synthesis** | | | | | | | | | | |
| Adamson 2003[1] | 82% | United States, International | Mixed | Unclear | Mixed | Mixed | Mixed | | | Costs per patient by floor plan type |
| Voigt 2018[141] | 86% | International | NR | NR | NR | Unclear | Routine | Operational efficiencies | | |

Health Economics Unit

30. Douglas CH, Douglas MR. Patient-centred improvements in health-care built environments: perspectives and design indicators. Health Expect. 2005 Sep;8(3):264–76.
31. Dowdeswell B, Erskine J, Heasman M. A Report for NHS Estates, England by the EU Health Property Network. :40.
32. Dowling DA, Blatz MA, Graham G. Mothers' Experiences Expressing Breast Milk for Their Preterm Infants: Does NICU Design Make a Difference? Advances in Neonatal Care. 2012 Dec;12(6):377–84.
33. Eberhard-Gran M, Eskild A, Opjordsmoen S, Schei B. Maternity care - sleep, rest and satisfaction. :10.
34. Edéll-Gustafsson U, Angelhoff C, Johnsson E, Karlsson J, Mörelius E. Hindering and buffering factors for parental sleep in neonatal care. A phenomenographic study. J Clin Nurs. 2015 Mar;24(5–6):717–27.
35. Ehrlander W, Ali F, Chretien KC. Multioccupancy hospital rooms: Veterans' experiences and preferences. Journal of Hospital Medicine. 2009;4(8):E22–7.
36. Erdeve O, Arsan S, Yigit S, Armangil D, Atasay B, Korkmaz A. The impact of individual room on rehospitalization and health service utilization in preterms after discharge. Acta paediatrica (Oslo, Norway : 1992). 2008 Oct;97(10):1351–7.
37. Erdeve O, Arsan S, Canpolat FE, Ertem IO, Karagol BS, Atasay B, et al. Does individual room implemented family- centered care contribute to mother-infant interaction in preterm deliveries necessitating neonatal intensive care unit hospitalization? American Journal of Perinatology. 2009 Feb 19;26(2):159–64.
38. Erickson C, Kattelmann K, Remington J, Cuirong Ren, Carol Helseth, Stevens D. Traditional open-bay versus single-family room neonatal intensive care unit: a comparison of selected nutrition outcomes. Research and Reports in Neonatology. 2011 Mar;15.
39. Everts RJ, Hanger HC, Jennings LC, Hawkins A, Sainsbury R. Outbreaks of influenza A among elderly hospital inpatients. The New Zealand Medical Journal. 1996 Jul 1;109(1026):272–4.
40. Felice Tong YY, Karunaratne S, Youlden D, Gupta S. The Impact of Room-Sharing on Length of Stay After Total Hip or Knee Arthroplasty: A Retrospective Study. Arthroplasty Today. 2021 Apr;8:289-294.e2.
41. Ferri M, Zygun DA, Harrison A, Stelfox HT. Evidence-based design in an intensive care unit: End-user perceptions. BMC Anesthesiology. 2015 Apr 25;15(1):1–9.
42. Florey L, Flynn R, Isles C. Patient Preferences for Single Rooms or Shared Accommodation in a District General Hospital. Scott Med J. 2009 May;54(2):5–8.
43. Foo CT, O'Driscoll DM, Ogeil RP, Lubman D, Young AC. Barriers to sleep in acute hospital settings. Sleep Breath. 2022 Jun;26(2):855–63.
44. Ford-Jones EL, Mindorff CM, Gold R, Petric M. THE INCIDENCE OF VIRAL-ASSOCIATED DIARRHEA AFTER ADMISSION TO A PEDIATRIC HOSPITAL. American Journal of Epidemiology. 1990 Apr;131(4):711–8.
45. Fraenkel Cj, Inghammar M, Söderlund-Strand A, Johansson PJH, Böttiger B. Risk factors for hospital norovirus outbreaks: impact of vomiting, genotype, and multi-occupancy rooms. Journal of Hospital Infection. 2018 Apr;98(4):398–403.

Health Economics Unit

103. Pyrke RJL, McKinnon MC, McNeely HE, Ahern C, Langstaff KL, Bieling PJ. Evidence-Based Design Features Improve Sleep Quality Among Psychiatric Inpatients. HERD. 2017 Oct;10(5):52–63.
104. Quach C, Shah R, Rubin LG. Burden of Healthcare-Associated Viral Respiratory Infections in Children's Hospitals. Journal of the Pediatric Infectious Diseases Society. 2018 Feb 19;7(1):18–24.
105. Real K, Fay L. Using Systems Theory to Examine Patient and Nurse Structures, Processes, and Outcomes in Centralized and Decentralized Units. :16.
106. Reed G, Schmid M. Nursing Implementation of Single-Room Maternity Care. Journal of Obstetric, Gynecologic & Neonatal Nursing. 1986 Sep;15(5):386–9.
107. Reid J, Wilson K, Anderson KE, Maguire CPJ. Older inpatients' room preference: single versus shared accommodation. Age and Ageing. 2015 Mar;44(2):331–3.
108. Roos AKØ, Skaug EA, Grøndahl VA, Helgesen AK. Trading company for privacy: A study of patients' experiences. Nurs Ethics. 2020 Jun;27(4):1089–102.
109. Rosbergen ICM, Tonello I, Clark RA, Grimley RS. Does hospital design impact on patient activity levels and time spent alone? Disability and Rehabilitation. 2022 Jun 19;44(13):3173–80.
110. Rowlands J, Noble S. How does the environment impact on the quality of life of advanced cancer patients? A qualitative study with implications for ward design. Palliat Med. 2008 Sep;22(6):768–74.
111. Sadatsafavi H, Niknejad B, Zadeh R, Sadatsafavi M. Do cost savings from reductions in nosocomial infections justify additional costs of single-bed rooms in intensive care units? A simulation case study. J Crit Care. 2016 Feb;31(1):194–200.
112. Sadatsafavi H, Niknejad B, Shepley M, Sadatsafavi M. Probabilistic Return-on-Investment Analysis of Single-Family Versus Open-Bay Rooms in Neonatal Intensive Care Units—Synthesis and Evaluation of Early Evidence on Nosocomial Infections, Length of Stay, and Direct Cost of Care. Journal of Intensive Care Medicine. 2019 Feb 1;34(2):115–25.
113. Sakr N, Hallit S, Mattar H. Incidence of and Factors Associated with New-Onset Insomnia Among Lebanese Hospitalised Patients: A single-centre study. Sultan Qaboos Univ Med J. 2021 Jun 21;21(2):e210-220.
114. Schalkers I, Dedding CWM, Bunders JFG. '[I would like] a place to be alone, other than the toilet' - Children's perspectives on paediatric hospital care in the Netherlands. Health Expect. 2015 Dec;18(6):2066–78.
115. Scottish Intercollegiate Guidelines Network. Sign 50: a guideline developer's handbook. Healthcare Improvement Scotland; 2014.
116. Singh I, Okeke J, Edwards C. Outcome of in-patient falls in hospitals with 100% single rooms and multi-bedded wards. Age and Ageing. 2015;44(6):1032–5.
117. Singh I. (PDF) Loneliness among Older People in Hospitals: A Comparative Study between Single Rooms and Multi- Bedded Wards to Evaluate Current Health Service within the Same Organisation [Internet]. 2016 [cited 2022 May 4]. Available from:

https://www.researchgate.net/publication/304495269_Loneliness_among_Older_People_in_Hospitals_A_Comparative_Study_between_Single_Rooms_and_Multi-_Bedded_Wards_to_Evaluate_Current_Health_Service_within_the_Same_Organisation
